# Opioid Analgesic Initiation During Pregnancy and the Risk of Adverse Maternal and Neonatal Outcomes: A Target Trial Emulation

**DOI:** 10.64898/2026.10.19.26365967

**Authors:** Bianca Varney, Malcolm B Gillies, Krista F. Huybrechts, Sonia Hernandez-Diaz, Sallie-Anne Pearson, Antonia Shand, Duong Tran, Jonathan Brett, Ximena Camacho, Claudia Bruno, Helga Zoega, Alys Havard

**Author notes:** Correspondence: Bianca Varney, National Drug and Alcohol Research Centre, Faculty of Medicine, UNSW Sydney, NSW 2052 Australia. Contributed equally as senior author (Helga Zoega, Alys Havard).

## Abstract

**Background:** We used sequential target trial emulation (TTE) to examine associations between opioid analgesic initiation and adverse maternal and neonatal outcomes while accounting for gestational timing, time at risk, and potential confounding.

**Methods:** Using the Early Life Course platform, comprising linked population-based data for births in New South Wales, Australia (2013–2019), we emulated a new trial at each gestational week, comparing women who initiated a prescribed opioid analgesic with eligible non-initiators. Baseline covariates were balanced using inverse probability weighting. Weighted logistic regression estimated odds ratios (ORs) for neonatal outcomes, postpartum haemorrhage, and severe maternal morbidity. Pooled logistic regression estimated hazard ratios (HRs) for other maternal outcomes, combined across gestational weeks using fixed-effects meta-analysis. Sensitivity analyses assessed unmeasured confounding, exposure misclassification, and potential reverse causation.

**Findings:** Among 511,357 pregnancies, over 80% of opioid initiators received only one dispensing. Opioid initiation was not associated with increased risks of placental abruption (HR 0.94, 95% CI 0.75–1.16), severe maternal morbidity (OR 1.05, 0.97–1.11), postpartum haemorrhage (OR 0.98, 0.95–1.02), low Apgar score (OR 0.99, 0.87–1.08), small for gestational age (OR 0.90, 0.85–0.95), or neonatal abstinence syndrome (OR 0.98, 0.72–1.30). Risks were elevated for PPROM (HR 1.10, 0.98–1.23), preterm birth (HR 1.26, 1.19–1.33), severe neonatal morbidity (OR 1.08, 1.02–1.17), and perinatal death (OR 1.29, 1.10–1.52). Perinatal death estimates attenuated in analyses addressing potential reverse causation.

**Interpretation:** Using a TTE approach that accounted for gestational timing and time at risk, we found no appreciable increase in most maternal or neonatal outcomes following predominantly short-term opioid initiation. Elevated risks of preterm birth, perinatal death, and third-trimester PPROM may reflect causal effects or confounding by pregnancy-related indications. Studies using time-appropriate designs and detailed information on prescribing indications and their timing are needed to clarify these associations.

**Research in Context:** *Evidence before this study:* We searched the electronic databases MEDLINE (PubMed) for published articles, without language restrictions, from 1 January 2000 until 9 September 2026. The search strategy included terms for exposure (e.g. analgesic opioids), population (e.g. pregnancy and neonatal) and maternal and neonatal outcomes (e.g. preterm birth). Our search identified 10 observational studies which examined maternal and neonatal outcomes following opioid analgesic exposure during pregnancy. In these studies, prenatal opioid exposure above certain levels was consistently associated with neonatal abstinence syndrome (NAS). Beyond NAS, findings were inconsistent: some studies reported increased risks of peripartum haemorrhage, stillbirth, small for gestational age, low Apgar scores, preterm birth, and placental abruption, whereas others found no increased risk. It remains unclear whether observed associations are causal or reflect residual confounding. Methodological limitations may also have obscured associations, including immortal time bias when treatment initiation and follow-up were not aligned, and analyses that did not adequately account for gestational timing and therefore may have missed periods of greater susceptibility. As opioid analgesics are among the few options for managing pain during pregnancy, high-quality evidence on their safety is essential for informed clinical decision-making.

*Added value of this study:* This study applied a sequential target trial emulation framework to estimate the effects of opioid analgesic initiation during pregnancy across a broad range of maternal and neonatal outcomes, aligning eligibility, treatment initiation, and follow-up while accounting for gestational timing. Opioid initiation was not associated with appreciably increased risks of most outcomes, including placental abruption, postpartum haemorrhage, severe maternal morbidity, and low Apgar scores. A small increase was observed for severe neonatal morbidity. Relative risks increased across trimesters for preterm birth, while PPROM and perinatal death were most elevated following third-trimester initiation. These findings may reflect opioid effects, although evolving pregnancy complications that both prompt opioid treatment and independently increase outcome risk remain a plausible explanation. NAS risk was not increased overall, although estimates were higher for oxycodone and higher oral morphine equivalent (OME) exposure.

*Implications of all available evidence:* Taken together with broader evidence showing no substantial increase in the risk of major congenital malformations or adverse neurodevelopmental outcomes, these findings are generally reassuring for short-term opioid analgesic use in pregnancy, particularly codeine. The observed signals for preterm birth, PPROM and perinatal death, should not, in isolation, deter clinically indicated use, but support existing guidance to use opioids at the lowest effective dose for the shortest necessary duration and highlight the need for further studies that can distinguish direct opioid effects from shared clinical pathways.

## Introduction

Pain is common in pregnancy and can substantially impair quality of life,^1^ yet pharmaceutical options for its management are limited.^2^ Paracetamol remains the only first-line therapy considered safe; non-steroidal anti-inflammatory drugs (NSAIDs) are not recommended due to potential risks of miscarriage early in pregnancy, and established risks of oligohydramnios and neonatal renal impairment after the first trimester.^3^ Opioid analgesics may be used for moderate to severe pain;^4^ however, their effects on maternal and neonatal outcomes remain uncertain.^5–10^ Despite this uncertainty, 0.4% to 19.1% of pregnant women in high-income countries are dispensed opioid analgesics during pregnancy.^11^ With few if any alternative pain management options during pregnancy, robust safety evidence for opioids is needed urgently.

Because randomised trials of opioid analgesics in pregnancy are unlikely to be performed, evidence must be drawn from observational data. A key methodological challenge in this population is time-related bias. Pregnancy duration determines the window of opportunity for exposure, meaning that women experiencing an adverse outcome early have less opportunity to be exposed to opioids than those whose pregnancies continue to term.^12^ This issue is particularly relevant for outcomes defined by or strongly related to gestational duration, such as preterm birth. Without accounting for gestational time at risk, studies may underestimate risks or even produce protective associations. For example, a Swedish registry study^7^ suggested opioid exposure was protective against very preterm birth, and a Canadian study^10^ reported decreasing risk with advancing trimester of exposure; neither of these studies adjusted for time at risk. By contrast, a US study^5^ employing time-varying exposure analysis found consistently elevated risks of preterm birth across both early and late pregnancy opioid exposure. However, this study evaluated only four outcomes, including placental abruption, preterm delivery, small for gestational age, and preeclampsia, leaving uncertainty regarding a broader range of maternal and neonatal outcomes associated with opioid initiation.

Target trial emulation (TTE) provides a structured causal framework to address common biases in observational pharmacoepidemiology, including immortal time bias, time-varying confounding, and selection bias.^13^ By specifying eligibility criteria, treatment strategies, treatment assignment, time zero, and follow-up, TTE aligns observational analyses with the protocol of a hypothetical randomised trial and minimises bias arising from misalignment of eligibility criteria, treatment initiation and follow-up. In this study, we implemented a TTE to investigate the risk of a broad range of adverse maternal and neonatal outcomes following initiation of prescription opioid analgesics in pregnancy using a large, whole-of-population cohort of pregnancies resulting in birth.

## Methods

We emulated a sequence of hypothetical target trials (appendix 1), classifying eligible individuals as treated or untreated in each gestational week of pregnancy. We estimated the observational analogue of the intention-to-treat effect of receiving opioid analgesics versus not receiving them, using linked, routinely collected healthcare data from the Early Life Course (ELC) data platform.^14^ Key specifications of the target trials and their observational emulations are summarised in appendix table 1, and reported according to the TARGET guidelines (appendix 2).^15^

### Data source

The Early Life Course Platform^14^ is a whole-of-population database, capturing pregnancies resulting in a live-or stillbirth (gestational age ≥20 weeks or birthweight ≥400g) in New South Wales (NSW), Australia’s most populous state, between July 2001 and December 2019. Women and their neonates were identified from the NSW Perinatal Data Collection (PDC) (*perinatal data*), a legislated and mandatory surveillance system capturing all births in NSW public and private hospitals, as well as homebirths. We used linked perinatal data, hospital admissions, opioid agonist therapy (OAT) for opioid use disorder (OUD), social security, mortality, Medicare Benefits Schedule (MBS) and Pharmaceutical Benefits Scheme (PBS) records. The PBS data collection includes records of medicines subsidised by the Australian Government and dispensed through community pharmacies, private hospitals, and outpatient services; private prescriptions and over-the-counter (OTC) medicines are not captured. PBS records also do not include the clinical indication for which a medicine was prescribed.^16^ Further details are provided in appendix 3.

### Study population

The study included women who conceived from July 1, 2013, to March 9, 2019. The earliest cohort entry date was chosen because the PBS database only captured complete dispensing records from 1 July 2012 onward, and at least 12 months of dispensing data before conception were required in order to ascertain new use. Conception date was calculated as the date of delivery – gestational age at delivery (days) + 14 days.^5^ The latest date for cohort entry was chosen to allow a full 40 weeks of follow up for all pregnancies. We excluded women with unknown maternal age, women aged <12 or >60 years, non-NSW residents, those with prior opioid dispensing within 90 days pre-conception, a cancer diagnosis or treatment in the prior year, treated or untreated OUD in the prior two years, those who gave birth at a single hospital that was not available for linkage, and mothers of neonates with missing or implausible gestational age (appendix table 2; appendix figure 1).

### Exposure

We emulated the target trial by creating a series of weekly trials. For each gestational week from week 3 (conception defined as the last day of week 2) through week 42, the last week in which an exposure could occur, we identified all eligible women and assigned them as opioid-initiators if they were dispensed their first opioid in that week, and as non-initiators otherwise (Figure 1). Gestational weeks were defined as consecutive 7-day intervals counted from the estimated date of conception (appendix 4). After opioid initiation, women were ineligible for subsequent trials. Women who did not initiate an opioid remained eligible to enter subsequent weekly trials, for a maximum of 40 trials. Opioid dispensings included all PBS-listed prescription opioid analgesics during the study period: buprenorphine, codeine, fentanyl, hydromorphone, methadone, morphine, oxycodone, tapentadol and tramadol (appendix table 3). In Australia, buprenorphine and methadone used to treat OUD require specialised authority approval and are supplied and recorded through a separate treatment records. Buprenorphine and methadone prescriptions captured in these dispensing data were therefore considered likely to represent use for pain.

**Figure 1:**
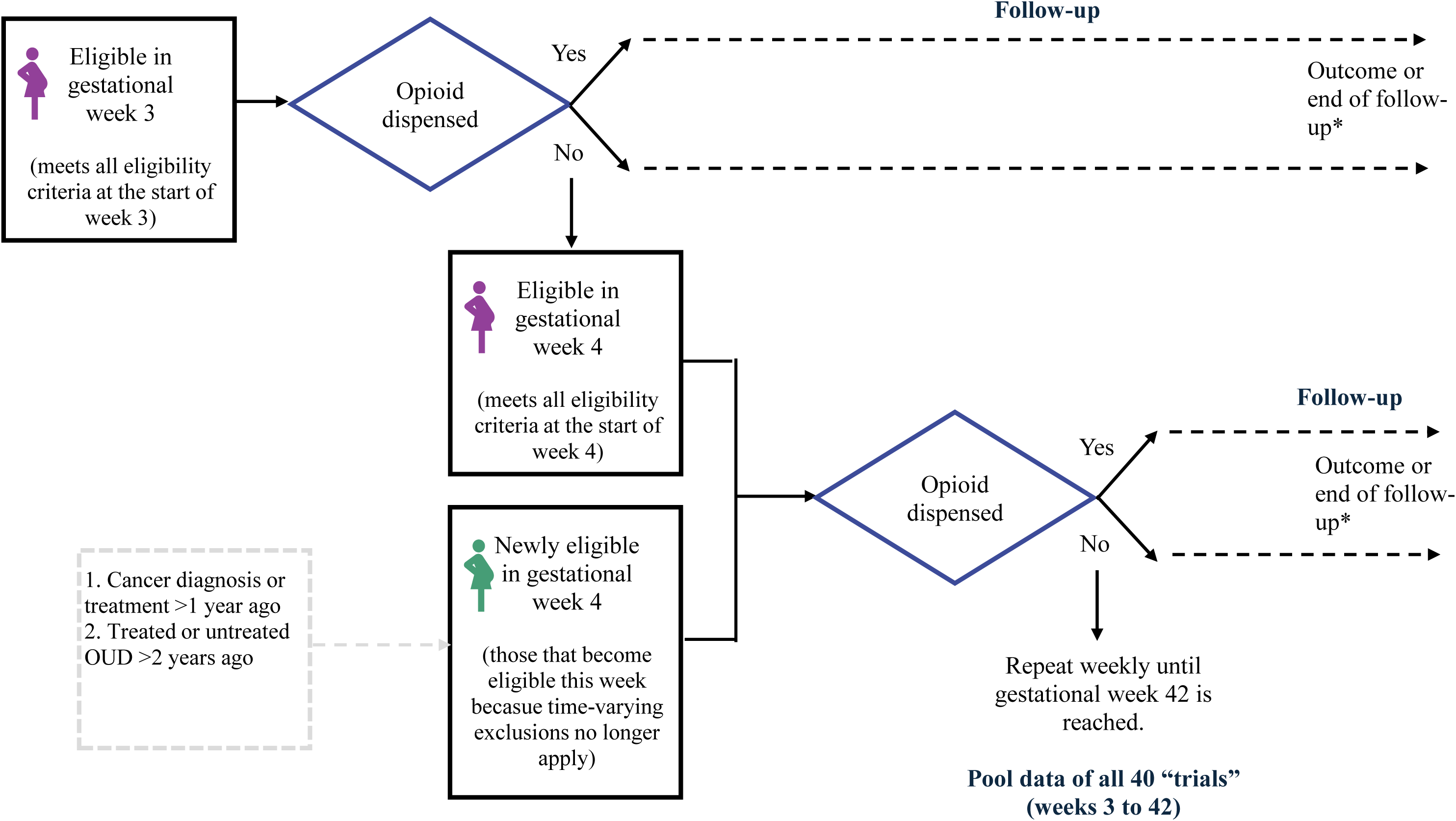
Schematic of the sequential trial design using gestational week as a new trial. Newly eligible women were those who met the eligibility criteria for the first time, for example because a previous cancer diagnosis or treatment had occurred more than one year earlier. *For each eligible woman, follow-up began at randomisation and ended at the occurrence of either the outcome of interest, discharge from the maternity ward (for mothers), 28 days after birth (for infants), death, loss to follow up, or December 31, 2019, whichever occurred first.

To characterise opioid analgesic use in this population, we calculated the proportion of women initiating opioid analgesics during each gestational week. We also determined the proportion dispensed 1, 2, or ≥3 opioid analgesics at any time during pregnancy, stratified by the trimester and type of opioid initiated. Finally, we calculated the dose of the initiated opioid analgesic in oral morphine equivalents (OME) milligrams (categorised as ≤100 OME vs >100 OME).

### Outcomes

Maternal pregnancy outcomes assessed included placental abruption, preterm premature rupture of membranes (PPROM, <37 weeks), and preterm birth (<37 weeks). Postpartum outcomes included haemorrhage and severe maternal morbidity complications, a composite indicator used to measure major maternal morbidities.^17^ Neonatal outcomes included low Apgar score (<7 at 5 minutes), perinatal death (stillbirth or neonatal death), small for gestational age (SGA) (<10th centile), neonatal abstinence syndrome (NAS), and a composite indicator of severe neonatal morbidity complications.^18^ Outcomes were identified using validated codes or algorithms in linked perinatal, hospital, and mortality records (appendix table 4).

### Covariates

We assumed that opioid initiation was random within levels of measured baseline covariates (appendix table 5). Covariates included maternal demographics and conditions, obstetric history, conditions associated with pain, other prescription medicine exposures and healthcare utilisation metrics as a proxy for overall morbidity burden. Covariates were ascertained during the 365 days prior to treatment assignment or for certain chronic conditions, up to the time of childbirth, reflecting prior evidence^19–21^ that increased contact with the hospital system during pregnancy and childbirth increases the identification of pre-existing conditions. Missing values for partner status, socio-economic disadvantage, remoteness, parity, and previous caesarean were imputed using the median or mode; missingness was ≤2.5% (appendix table 5).

### Statistical analysis

We used stabilised inverse probability of treatment weighting (IPTW) based on propensity scores to balance patient characteristics between opioid initiators and non-initiators within each weekly trial (i.e., within each gestational week of pregnancy). Weighting effectiveness was assessed by comparing baseline characteristics before and after weighting, with absolute standardised differences ≤0.1 indicating adequate balance. We incorporated stabilised weights into models to estimate the average treatment effect on the treated. For non-time-to-event outcomes, including neonatal outcomes and maternal postpartum outcomes (postpartum haemorrhage and severe maternal morbidity), we used weighted logistic regression fitted to the pooled results from all weekly trials to estimate odds ratios (OR). For time-to-event maternal pregnancy outcomes (placental abruption, PPROM, and preterm birth), we used pooled logistic regression to estimate hazard ratios (HR) according to the gestational week of treatment initiation, then combined the gestational-week-specific estimates into an overall effect using fixed-effects meta-analysis. For these outcomes, we also estimated crude cumulative incidence non-parametrically using the Kaplan-Meier method. For all analyses, 95% confidence intervals (CI) were estimated using nonparametric percentile bootstrapping with 200 samples to account for clustering from the repeated inclusion of individuals (women in multiple trials, and women with multiple pregnancies) (refer to appendix 5 for a more detailed description of the statistical analyses).

### Subgroup analyses

In sub-group analyses, we examined effects by trimester of treatment initiation (1, 2, or 3), dose in OME (≤100 OME and >100 OME) and opioid type, restricted to the three most commonly dispensed opioids: codeine, oxycodone, and tramadol.

### Sensitivity analysis

We conducted several sensitivity analyses. Firstly, to test the robustness to confounding by indication we (i) restricted the cohort to women with a history of chronic conditions associated with pain (appendix table 5), and (ii) performed an active comparator analysis using initiation of non-codeine opioids as the treatment arm and initiation of codeine as the comparator. Secondly, given that low-dose codeine products were available over the counter in Australia until they were up-scheduled to prescription-only medicines on 1 February 2018, we assessed the robustness of our results to potential exposure misclassification arising from unrecorded low-dose codeine use by limiting the analysis to pregnancies conceived from 1 February 2018, onwards. Finally, we analysed stillbirth and neonatal death separately, to assess the contribution of each to the composite outcome of perinatal death.

### Post-hoc analyses

Given that stillbirth is recorded on the delivery date although fetal death may have occurred several days earlier, we conducted a post-hoc analysis of perinatal death in which the recorded childbirth date was shifted seven days earlier to reduce the potential for opioid dispensings occurring after fetal death or diagnosis of a life-limiting fetal condition to be classified as exposure. This approach may, however, exclude opioid exposures occurring within seven days of the outcome that could plausibly have an immediate causal effect.

In a further post-hoc analysis, we examined the extent to which maternal and neonatal outcomes co-occurred within the same pregnancy. For each outcome, we calculated the proportion of affected pregnancies in which each other maternal or neonatal outcome also occurred, separately among opioid initiators and non-initiators. This analysis was used to assess whether observed associations across multiple outcomes arose largely within the same pregnancies, which may provide insight into whether findings reflect correlated outcomes or shared underlying pregnancy complications rather than independent effects.

All analysis were performed using SAS (SAS Institute, Cary, NC, USA) and R v.4.2.2^22^ using the following packages: *gee*,^23^ *speedglm*^24^ and *boot*.^25, 26^

## Results

After exclusions, 511,357 pregnancies (398,944 women) met our eligibility criteria. Of these, 26,984 (5.3%) initiated an opioid at some point during pregnancy and 484,373 (94.7%) did not. In the weekly trial emulation, pregnancies could contribute to multiple eligible weeks until opioid initiation or the end of eligibility; this corresponded to 18,360,776 person-trials: 26,984 initiator person-trials and 18,333,792 non-initiator person-trials (appendix figure 2). Median gestational age at trial entry was 21 weeks (IQR 12–30) among initiator person-trials and 23 weeks (IQR 13-33) among non-initiator person-trials. Corresponding median follow-up from trial entry to birth was 16.9 weeks (IQR 5.9–26.9) and 18.9 weeks (IQR 9.9–27.9), respectively. The difference in follow-up partly reflects differences in the distribution of gestational week at entry across the pooled sequential trials.

Among opioid initiators, 86.7% initiated codeine, including codeine with paracetamol, 11.5% initiated oxycodone, and 1.5% initiated tramadol (appendix table 6). Most initiators (81.2%) had only one opioid dispensing during pregnancy. This was more common among those assigned to treatment in later pregnancy (trimester 1: 74.2%; trimester 2: 78.9%; trimester 3: 87.6%), consistent with less remaining gestational time for a subsequent dispensing and varied by opioid type (codeine: 82.5%; oxycodone: 73.2%; tramadol: 69.4%) (appendix figure 3).

The proportion of opioid initiators was relatively consistent across gestational weeks but peaked between 39 and 41 weeks (appendix figure 4; appendix table 7). Based on the first dispensing, median OME was highest for tramadol (200 mg; IQR 200–200), followed by oxycodone (150 mg; IQR 150–150) and codeine (60 mg; IQR 60–60). Most initiators (86.8%) started on a low-dose opioid analgesic (≤100 OME), while 13.2% (n=3,454) initiated at doses >100 OME.

Compared with non-initiators, a higher proportion of opioid initiators were younger (aged <25 years), had had previous children, did not have a partner, were social security beneficiaries, and had had a previous adverse pregnancy outcome; a higher proportion also smoked during pregnancy, had chronic illnesses, used concomitant medicines, and had greater healthcare utilisation (Table 1). After weighting, baseline covariates were well balanced (appendix figure 5). Characteristics and covariate balance for sensitivity analyses are presented in the appendix (tables 8-10; figures 5-8).

**Table 1:**
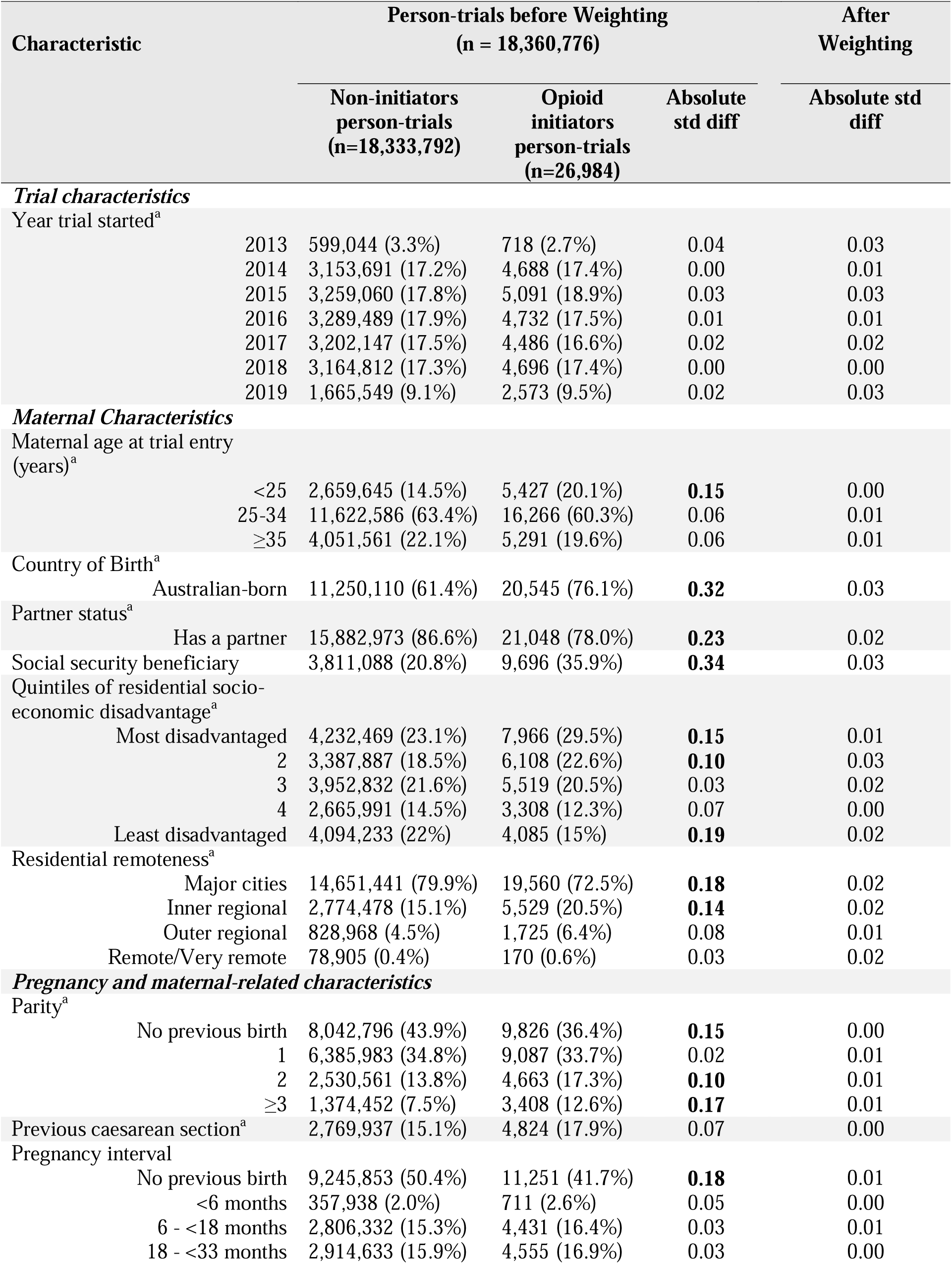
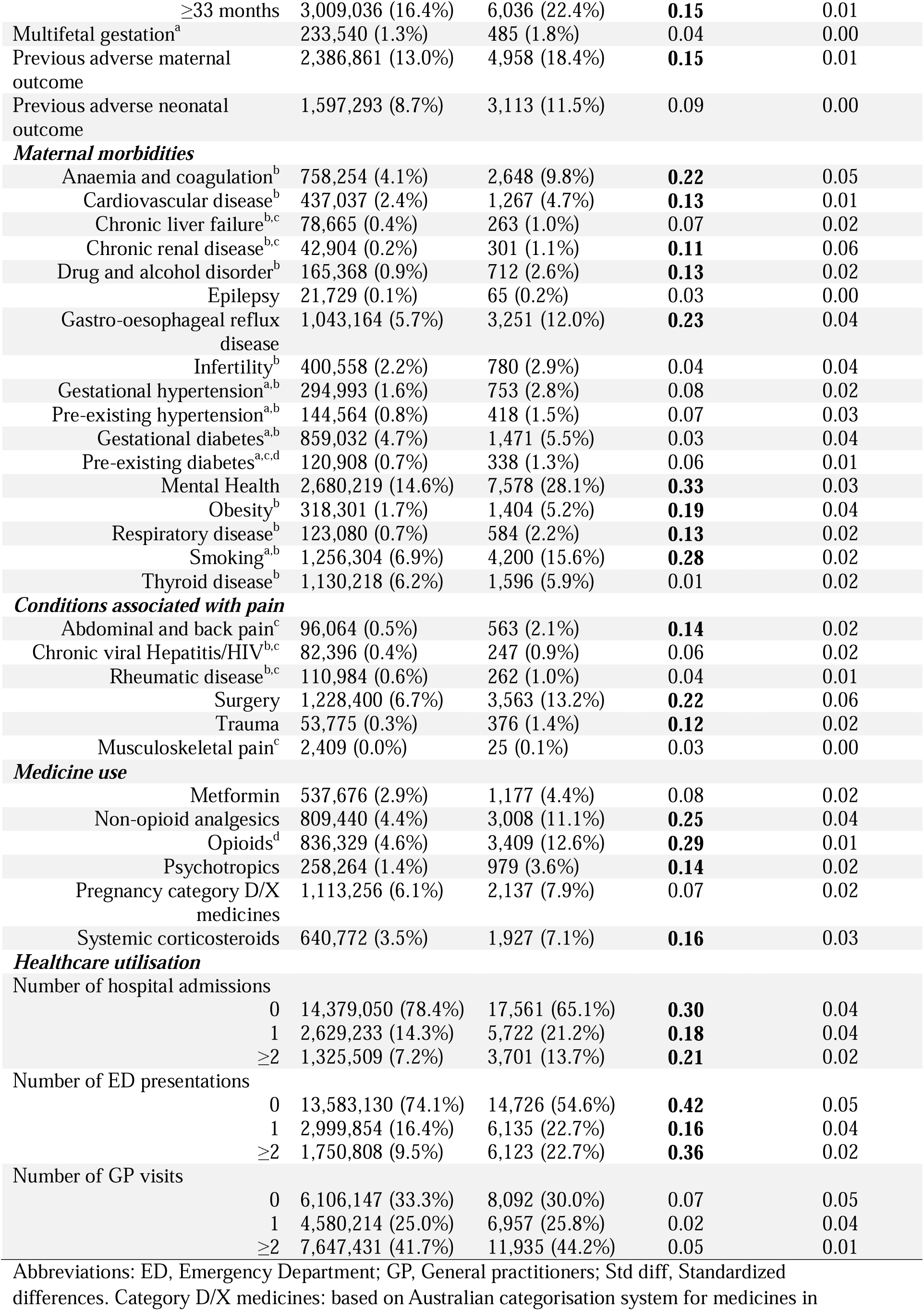
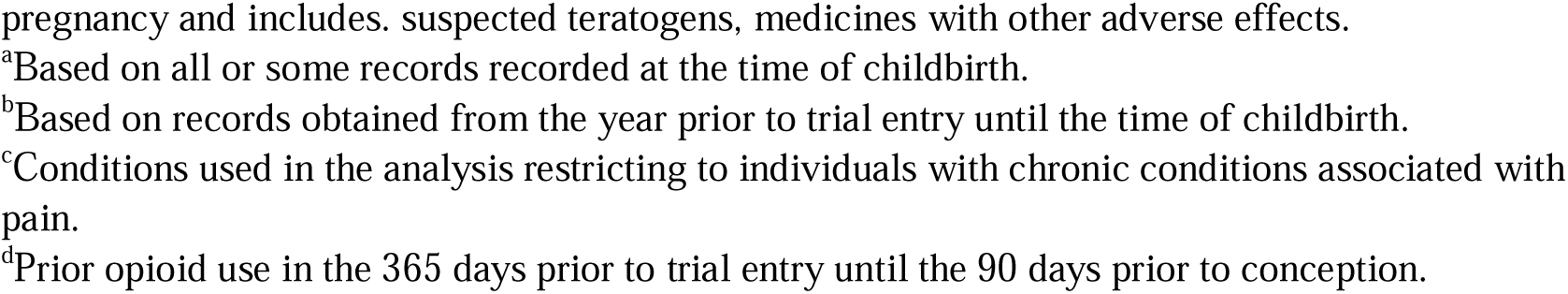
Characteristics and absolute standardised mean differences of opioid analgesic initiators and non-initiators before and after weighting.

| Characteristic | Person-trials before Weighting<br>(n = 18,360,776) |  |  | After Weighting |
| --- | --- | --- | --- | --- |
|  | Non-initiators<br>person-trials<br>(n=18,333,792) | Opioid<br>initiators<br>person-trials<br>(n=26,984) | Absolute<br>std diff | Absolute std<br>diff |
| ***Trial characteristics*** |  |  |  |  |
| Year trial started<sup>a</sup> |  |  |  |  |
| 2013 | 599,044 (3.3%) | 718 (2.7%) | 0.04 | 0.03 |
| 2014 | 3,153,691 (17.2%) | 4,688 (17.4%) | 0.00 | 0.01 |
| 2015 | 3,259,060 (17.8%) | 5,091 (18.9%) | 0.03 | 0.03 |
| 2016 | 3,289,489 (17.9%) | 4,732 (17.5%) | 0.01 | 0.01 |
| 2017 | 3,202,147 (17.5%) | 4,486 (16.6%) | 0.02 | 0.02 |
| 2018 | 3,164,812 (17.3%) | 4,696 (17.4%) | 0.00 | 0.00 |
| 2019 | 1,665,549 (9.1%) | 2,573 (9.5%) | 0.02 | 0.03 |
| ***Maternal Characteristics*** |  |  |  |  |
| Maternal age at trial entry<br>(years)<sup>a</sup> |  |  |  |  |
| <25 | 2,659,645 (14.5%) | 5,427 (20.1%) | **0.15** | 0.00 |
| 25-34 | 11,622,586 (63.4%) | 16,266 (60.3%) | 0.06 | 0.01 |
| ≥35 | 4,051,561 (22.1%) | 5,291 (19.6%) | 0.06 | 0.01 |
| Country of Birth<sup>a</sup> |  |  |  |  |
| Australian-born | 11,250,110 (61.4%) | 20,545 (76.1%) | **0.32** | 0.03 |
| Partner status<sup>a</sup> |  |  |  |  |
| Has a partner | 15,882,973 (86.6%) | 21,048 (78.0%) | **0.23** | 0.02 |
| Social security beneficiary | 3,811,088 (20.8%) | 9,696 (35.9%) | **0.34** | 0.03 |
| Quintiles of residential socio-economic disadvantage<sup>a</sup> |  |  |  |  |
| Most disadvantaged | 4,232,469 (23.1%) | 7,966 (29.5%) | **0.15** | 0.01 |
| 2 | 3,387,887 (18.5%) | 6,108 (22.6%) | **0.10** | 0.03 |
| 3 | 3,952,832 (21.6%) | 5,519 (20.5%) | 0.03 | 0.02 |
| 4 | 2,665,991 (14.5%) | 3,308 (12.3%) | 0.07 | 0.00 |
| Least disadvantaged | 4,094,233 (22%) | 4,085 (15%) | **0.19** | 0.02 |
| Residential remoteness<sup>a</sup> |  |  |  |  |
| Major cities | 14,651,441 (79.9%) | 19,560 (72.5%) | **0.18** | 0.02 |
| Inner regional | 2,774,478 (15.1%) | 5,529 (20.5%) | **0.14** | 0.02 |
| Outer regional | 828,968 (4.5%) | 1,725 (6.4%) | 0.08 | 0.01 |
| Remote/Very remote | 78,905 (0.4%) | 170 (0.6%) | 0.03 | 0.02 |
| ***Pregnancy and maternal-related characteristics*** |  |  |  |  |
| Parity<sup>a</sup> |  |  |  |  |
| No previous birth | 8,042,796 (43.9%) | 9,826 (36.4%) | **0.15** | 0.00 |
| 1 | 6,385,983 (34.8%) | 9,087 (33.7%) | 0.02 | 0.01 |
| 2 | 2,530,561 (13.8%) | 4,663 (17.3%) | **0.10** | 0.01 |
| ≥3 | 1,374,452 (7.5%) | 3,408 (12.6%) | **0.17** | 0.01 |
| Previous caesarean section<sup>a</sup> | 2,769,937 (15.1%) | 4,824 (17.9%) | 0.07 | 0.00 |
| Pregnancy interval |  |  |  |  |
| No previous birth | 9,245,853 (50.4%) | 11,251 (41.7%) | **0.18** | 0.01 |
| <6 months | 357,938 (2.0%) | 711 (2.6%) | 0.05 | 0.00 |
| 6 - <18 months | 2,806,332 (15.3%) | 4,431 (16.4%) | 0.03 | 0.01 |
| 18 - <33 months | 2,914,633 (15.9%) | 4,555 (16.9%) | 0.03 | 0.00 |
| ≥33 months | 3,009,036 (16.4%) | 6,036 (22.4%) | **0.15** | 0.01 |
| Multifetal gestation<sup>a</sup> | 233,540 (1.3%) | 485 (1.8%) | 0.04 | 0.00 |
| Previous adverse maternal outcome | 2,386,861 (13.0%) | 4,958 (18.4%) | **0.15** | 0.01 |
| Previous adverse neonatal outcome | 1,597,293 (8.7%) | 3,113 (11.5%) | 0.09 | 0.00 |
| **Maternal morbidities** |  |  |  |  |
| Anaemia and coagulation<sup>b</sup> | 758,254 (4.1%) | 2,648 (9.8%) | **0.22** | 0.05 |
| Cardiovascular disease<sup>b</sup> | 437,037 (2.4%) | 1,267 (4.7%) | **0.13** | 0.01 |
| Chronic liver failure<sup>b,c</sup> | 78,665 (0.4%) | 263 (1.0%) | 0.07 | 0.02 |
| Chronic renal disease<sup>b,c</sup> | 42,904 (0.2%) | 301 (1.1%) | **0.11** | 0.06 |
| Drug and alcohol disorder<sup>b</sup> | 165,368 (0.9%) | 712 (2.6%) | **0.13** | 0.02 |
| Epilepsy | 21,729 (0.1%) | 65 (0.2%) | 0.03 | 0.00 |
| Gastro-oesophageal reflux disease | 1,043,164 (5.7%) | 3,251 (12.0%) | **0.23** | 0.04 |
| Infertility<sup>b</sup> | 400,558 (2.2%) | 780 (2.9%) | 0.04 | 0.04 |
| Gestational hypertension<sup>a,b</sup> | 294,993 (1.6%) | 753 (2.8%) | 0.08 | 0.02 |
| Pre-existing hypertension<sup>a,b</sup> | 144,564 (0.8%) | 418 (1.5%) | 0.07 | 0.03 |
| Gestational diabetes<sup>a,b</sup> | 859,032 (4.7%) | 1,471 (5.5%) | 0.03 | 0.04 |
| Pre-existing diabetes<sup>a,c,d</sup> | 120,908 (0.7%) | 338 (1.3%) | 0.06 | 0.01 |
| Mental Health | 2,680,219 (14.6%) | 7,578 (28.1%) | **0.33** | 0.03 |
| Obesity<sup>b</sup> | 318,301 (1.7%) | 1,404 (5.2%) | **0.19** | 0.04 |
| Respiratory disease<sup>b</sup> | 123,080 (0.7%) | 584 (2.2%) | **0.13** | 0.02 |
| Smoking<sup>a,b</sup> | 1,256,304 (6.9%) | 4,200 (15.6%) | **0.28** | 0.02 |
| Thyroid disease<sup>b</sup> | 1,130,218 (6.2%) | 1,596 (5.9%) | 0.01 | 0.02 |
| **Conditions associated with pain** |  |  |  |  |
| Abdominal and back pain<sup>c</sup> | 96,064 (0.5%) | 563 (2.1%) | **0.14** | 0.02 |
| Chronic viral Hepatitis/HIV<sup>b,c</sup> | 82,396 (0.4%) | 247 (0.9%) | 0.06 | 0.02 |
| Rheumatic disease<sup>b,c</sup> | 110,984 (0.6%) | 262 (1.0%) | 0.04 | 0.01 |
| Surgery | 1,228,400 (6.7%) | 3,563 (13.2%) | **0.22** | 0.06 |
| Trauma | 53,775 (0.3%) | 376 (1.4%) | **0.12** | 0.02 |
| Musculoskeletal pain<sup>c</sup> | 2,409 (0.0%) | 25 (0.1%) | 0.03 | 0.00 |
| **Medicine use** |  |  |  |  |
| Metformin | 537,676 (2.9%) | 1,177 (4.4%) | 0.08 | 0.02 |
| Non-opioid analgesics | 809,440 (4.4%) | 3,008 (11.1%) | **0.25** | 0.04 |
| Opioids<sup>d</sup> | 836,329 (4.6%) | 3,409 (12.6%) | **0.29** | 0.01 |
| Psychotropics | 258,264 (1.4%) | 979 (3.6%) | **0.14** | 0.02 |
| Pregnancy category D/X medicines | 1,113,256 (6.1%) | 2,137 (7.9%) | 0.07 | 0.02 |
| Systemic corticosteroids | 640,772 (3.5%) | 1,927 (7.1%) | **0.16** | 0.03 |
| **Healthcare utilisation** |  |  |  |  |
| Number of hospital admissions |  |  |  |  |
| 0 | 14,379,050 (78.4%) | 17,561 (65.1%) | **0.30** | 0.04 |
| 1 | 2,629,233 (14.3%) | 5,722 (21.2%) | **0.18** | 0.04 |
| ≥2 | 1,325,509 (7.2%) | 3,701 (13.7%) | **0.21** | 0.02 |
| Number of ED presentations |  |  |  |  |
| 0 | 13,583,130 (74.1%) | 14,726 (54.6%) | **0.42** | 0.05 |
| 1 | 2,999,854 (16.4%) | 6,135 (22.7%) | **0.16** | 0.04 |
| ≥2 | 1,750,808 (9.5%) | 6,123 (22.7%) | **0.36** | 0.02 |
| Number of GP visits |  |  |  |  |
| 0 | 6,106,147 (33.3%) | 8,092 (30.0%) | 0.07 | 0.05 |
| 1 | 4,580,214 (25.0%) | 6,957 (25.8%) | 0.02 | 0.04 |
| ≥2 | 7,647,431 (41.7%) | 11,935 (44.2%) | 0.05 | 0.01 |

Abbreviations: ED, Emergency Department; GP, General practitioners; Std diff, Standardized differences. Category D/X medicines: based on Australian categorisation system for medicines in
pregnancy and includes. suspected teratogens, medicines with other adverse effects.
<sup>a</sup>Based on all or some records recorded at the time of childbirth.
<sup>b</sup>Based on records obtained from the year prior to trial entry until the time of childbirth.
<sup>c</sup>Conditions used in the analysis restricting to individuals with chronic conditions associated with pain.
<sup>d</sup>Prior opioid use in the 365 days prior to trial entry until the 90 days prior to conception.

### Risk of maternal and neonatal outcomes

Compared with non-initiators, weighted relative risk estimates for maternal outcomes among opioid analgesic initiators during pregnancy were 0.94 (95% CI 0.75-1.16) for placental abruption, 1.10 (95% CI 0.98-1.23) for PPROM, 1.26 (95% CI 1.19-1.33) for preterm birth, 1.05 (95% CI 0.97-1.11) for severe maternal morbidity, and 0.98 (95% CI 0.95-1.02) for postpartum haemorrhage (Figure 2). Cumulative incidence curves for initiators and non-initiators overlapped for both placental abruption and PPROM, except for third-trimester PPROM, where the cumulative risk for initiators was higher. In contrast, the curves for preterm birth were consistently higher for initiators (appendix figures 9-11, respectively). For neonatal outcomes, risk estimates for those born to opioid initiators compared with non-initiators were 1.08 (95% CI 1.02-1.17) for severe neonatal morbidity, 1.29 (95% CI 1.10-1.52) for perinatal death, 0.99 (95% CI 0.87-1.08) for low Apgar score, 0.90 (95% CI 0.85-0.95) for SGA, and 0.98 (95% CI 0.72-1.30) for neonatal abstinence syndrome (NAS) (Figure 2).

**Figure 2:**
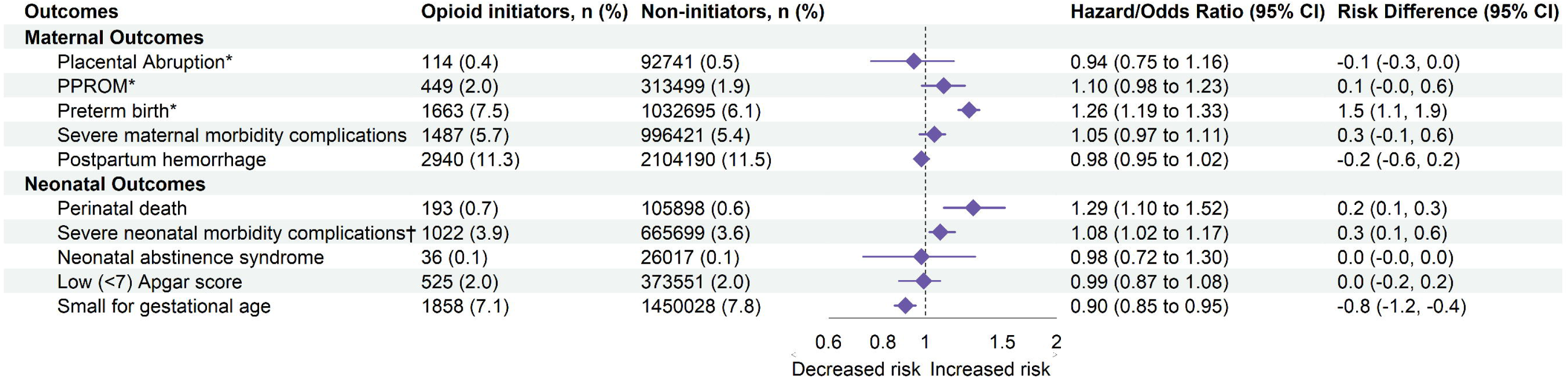
Adjusted risks (odds ratios and hazard ratios) and risk differences (expressed as percentages) for adverse maternal and neonatal outcomes following opioid initiation versus non-initiation during pregnancy. The number and proportion of each outcome within each treatment group are shown as n (%). * Risk estimates represent hazard ratios; remaining risk estimates represent odds ratios. † For severe neonatal morbidity, some individual outcomes reported separately above, including very preterm birth (gestational age <32 weeks), stillbirth and neonatal death, are also included within the composite indicator. None of the individual outcomes were components of the severe maternal morbidity composite.

### Subgroup analysis

Effect estimates from subgroup analyses, by trimester of opioid initiation, opioid dose, and opioid type, were generally consistent with those observed in the primary analysis considering confidence intervals for low Apgar score, SGA, postpartum haemorrhage, severe maternal morbidity complications (Figure 3; appendix figure 12). For perinatal death, odds ratios were close to unity for initiation in the first trimester (OR 1.02, 95% CI 0.68-1.33), but were higher with later initiation, particularly in the second (OR 1.56, 95% CI 1.25-1.88) and third trimesters (OR 1.71, 95% CI 0.98-2.46), with no clear evidence of a dose-response relationship (low OME: OR 1.32, 95% CI 1.11-1.56; high OME: OR 1.01, 95% CI 0.95-2.46). Estimates for NAS were higher among women dispensed higher OME doses (OR 1.73, 95% CI 0.83-3.45) and those initiating oxycodone (OR 1.85, 95% CI 0.58-3.82), although confidence intervals were wide. For preterm birth, hazard ratios increased progressively with later trimester of initiation (first trimester: 1.13, 95% CI 1.01-1.26; second: 1.27, 95% CI 1.16-1.38; third: 1.37, 95% CI 1.24-1.51). For PPROM, estimates were near null for first (1.09, 95% CI 0.88-1.37) and second trimester initiation (1.07, 95% CI 0.90-1.28), but higher for third trimester initiation (1.30, 95% CI 1.02-1.65), although confidence intervals were largely overlapping.

**Figure 3:**
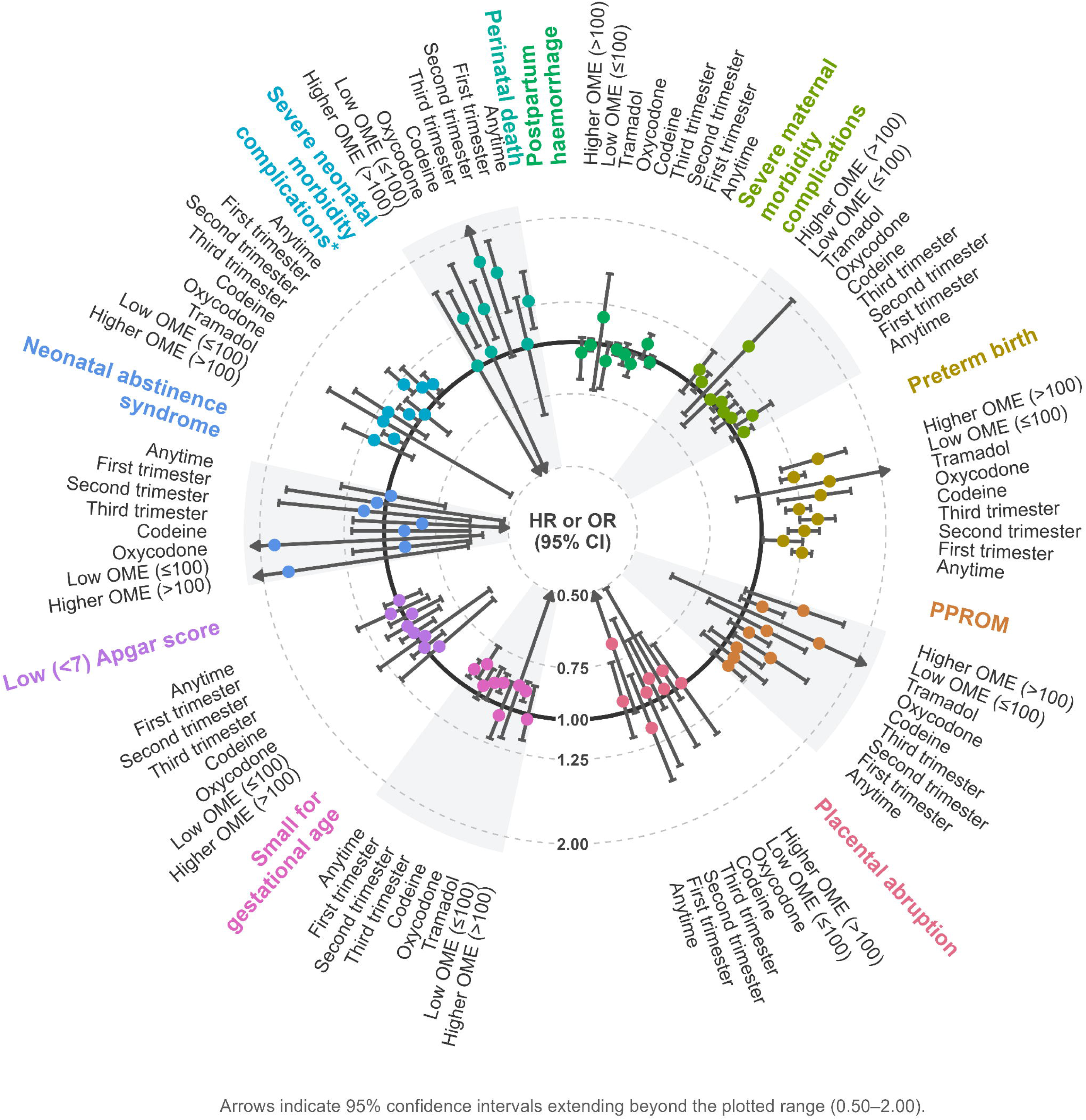
Adjusted risks of adverse maternal and neonatal non–time-to-event outcomes following opioid initiation versus non-initiation during pregnancy. Greater detail of these analyses is provided in appendix figure 12.

### Sensitivity analyses

Effect estimates were elevated for both stillbirth (OR 1.28, 95% CI 1.08-1.58) and neonatal death (OR 1.29, 95% CI 0.88-1.60) (appendix figure 13). Sensitivity analyses addressing residual confounding (restricting to women with conditions associated with chronic pain and using codeine as an active comparator) yielded estimates generally consistent with those from the main analysis when accounting for the width of the confidence intervals (appendix figure 14 and 15), except for perinatal death in the restricted cohort (OR 1.13, 95% CI 0.30-1.74) and with the active comparator (OR 0.61, 95% CI 0.28-0.87). Restricting the analysis to February 2018 onwards, when low-dose codeine was up-scheduled, produced results generally consistent with the main findings (appendix figure 16), except for perinatal death. For this outcome, estimates were attenuated (OR 1.01, 95% CI 0.50-1.84), with wide confidence intervals.

### Post-hoc analyses

In the post hoc analysis addressing potential reverse causation, estimates for perinatal death were attenuated but remained slightly elevated for opioid initiation at any time during pregnancy (OR 1.10, 95% CI 0.91–1.36), in the second trimester (OR 1.20, 95% CI 0.90– 1.62), and in the third trimester (OR 1.23, 95% CI 0.60–2.03); confidence intervals were wide (appendix figure 17). Preterm birth was the most common co-occurring maternal outcome, affecting approximately 80% of pregnancies with perinatal death. Co-occurrence patterns were otherwise similar between opioid initiators and non-initiators (appendix figure 18).

## Discussion

Using real-world, whole-of-population data, we performed a sequential target trial emulation to evaluate the risk of adverse maternal and neonatal outcomes following prescription opioid analgesic initiation in pregnancy. We found no or small increased risk (<20%) for most maternal and neonatal adverse outcomes overall and across different trimesters and opioid type (codeine, oxycodone and tramadol), including placental abruption, postpartum haemorrhage, severe maternal and neonatal morbidity, low Apgar score, SGA, and NAS. Given the observational design of this study, the potential for residual or unmeasured confounding, and the small absolute differences in risk between groups, these small elevations are unlikely to reflect clinically meaningful effects. More than 80% of women received only a single opioid dispensing, therefore, findings primarily reflect short-term use and should not be extrapolated to persistent or chronic use, which may carry different risks.

We observed increased risks of preterm birth, with relative risks increasing across trimesters, as well as higher risks of PPROM and perinatal death following third-trimester initiation. The preterm birth finding is consistent with the only prior study addressing time-related bias;^5^ to our knowledge, PPROM has not previously been examined. These findings could reflect effects of opioid exposure, potentially with greater susceptibility later in pregnancy. However, given that these outcomes are clinically interrelated, they may also reflect confounding by pregnancy conditions or symptoms that both prompted opioid treatment and independently increased the risk of these outcomes. For example, women may be prescribed opioids for abdominal, pelvic, or other pain associated with evolving pregnancy complications that subsequently lead to PPROM, preterm birth, or stillbirth. In this setting, opioid initiation may therefore occur in response to early manifestations of the outcome process rather than being its cause.

Our sensitivity analyses addressing confounding by indication produced slightly attenuated but imprecise estimates, and restriction to women with chronic pain may not eliminate residual confounding by pain severity nor confounding from painful pregnancy complications. A Swedish study also found little evidence of an association with preterm birth when opioid initiators were compared with discontinuers (adjusted OR 1.05, 95% CI 0.96–1.14), suggesting that differences in the underlying indication for treatment may contribute to the association observed in conventional comparisons.^9^ The interrelationship among the three outcomes was also evident in our data, with approximately 80% of perinatal deaths occurring in pregnancies complicated by preterm birth and 15% in those with PPROM. Rather, this co-occurrence may partly reflect pregnancies with underlying complications that increased the risk of both medically indicated preterm delivery and perinatal death, rather than a single pathway from opioid exposure to PPROM or spontaneous preterm birth and subsequently to perinatal death.

The perinatal death association warrants additional caution because it was sensitive to analytic decisions: estimates varied across sensitivity analyses and were attenuated in post hoc analyses addressing reverse causation. This pattern suggests that confounding from pregnancy-specific indications and reverse causation may explain at least part of the association rather than it being a direct effect of opioids. Similar uncertainty applies to previous research. A large Ontario cohort reported increased risks of preterm birth (RR 1.3, 95% CI 1.2–1.3) and stillbirth (RR 1.6, 95% CI 1.4–1.8) following prenatal opioid exposure, but these associations could reflect causal effects, residual confounding by pregnancy-related indications, reverse causation, or a combination of these mechanisms. Larger studies with detailed information on treatment indication, and pregnancy complications and their timing are needed to distinguish direct effects from these alternative explanations.

Although opioid exposure is an established risk factor for NAS, particularly exposure close to birth, prolonged use, or stronger opioids such as oxycodone,^28, 29^ we found no overall increase in risk. Risks were higher with oxycodone and higher OME doses, consistent with prior literature, but confidence intervals were wide, and no elevation was seen with third-trimester exposure. These findings, alongside analyses showing that most women received a single low-dose codeine dispensing (81.2%) and that third-trimester initiation occurred late in pregnancy (weeks 39-41), suggest that opioid exposure in this population was unlikely to be sufficient in duration or intensity to result in neonatal withdrawal.

Overall, the evidence provides reassurance regarding the safety of short-term opioid analgesic use when clinically indicated, while reinforcing guidance to use the lowest effective dose for the shortest necessary duration, as safety has not been established for prolonged or higher-dose exposure or for all individual opioids, and the risks of NAS and dependence increase with greater exposure.

While our study has advantages including the use of a whole-of-population cohort and explicit emulation of a hypothetical target trial, it has some important limitations. Firstly, residual confounding is possible, impacting our ability to successfully emulate the randomisation component of the target trial. Although we adjusted for baseline covariates measured at the time of opioid initiation, we were unable to account for the indication for opioid analgesic use which is not recorded in the PBS data. While pelvic girdle pain, a common indication for opioid use during pregnancy, is unlikely to meaningfully confound the observed elevated associations, less common indications, e.g. surgery or rheumatoid arthritis, and pregnancy complications, such as threatened preterm labour, may have greater potential for confounding. We addressed differences in chronic conditions in sensitivity analyses but could not fully account for differences in the types and severity of underlying conditions or pregnancy complications, which may have contributed to residual confounding and overestimation of the observed associations between opioid analgesic initiation and these outcomes. Second, our findings may be affected by exposure misclassification, as medicine dispensing records confirm that a medicine was supplied but not that it was consumed; this may lead to false-positive exposures. This may be particularly relevant for opioid analgesics, which are often used as needed and may be avoided during pregnancy because of safety concerns.^30^ Such non-differential misclassification would generally bias estimates towards the null, suggesting that observed associations may underestimate true effects. Further, some false-negative exposure is likely given the PBS data do not capture opioids supplied to public hospital inpatients, on private prescriptions, or as OTC medicines, estimated to account for 13.8% of all prescribed opioids in Australia in 2015.^31^ Analyses restricted to pregnancies conceived after the up-scheduling of codeine in 2018 suggest OTC misclassification had limited impact.^32, 33^

## Conclusion

This sequential TTE, which overcomes the time-related biases present in existing literature, found no appreciable increase in the risk of most adverse maternal or neonatal outcomes following predominantly short-term opioid initiation in pregnancy. Elevated risks of preterm birth, perinatal death, and third-trimester PPROM may reflect direct effects, shared pathways, or reverse causation and require investigation using detailed prescription indication and emerging symptom data. Overall, findings are reassuring for short-term opioid use in pregnancy but should not be extrapolated to prolonged or higher-dose exposure.

## Data Availability

The datasets we used were constructed with the permission of each of the source data custodians and with specific ethics approvals. We do not have permission to share patient-level data because of the highly confidential nature of the data. Permission to access the data is restricted to researchers named and approved by relevant human research ethics committees.

## Acknowledgements

This research was completed using the Early Life Course (ELC) data platform. The authors would like to thank data providers: the Australian Government Department of Health and Aged Care, NSW Ministry of Health and the Australian Government Department of Social Services. The authors acknowledge that Cause of Death Unit Record File (COD URF) is provided by the Australian Coordinating Registry for the COD URF on behalf of the NSW Registry of Births, Deaths and Marriages, NSW Coroner and the National Coronial Information System. The COD URF is held by the NSW Ministry of Health Secure Analytics for Population Health Research and Intelligence. The authors gratefully acknowledge the assistance of the Centre for Health Record Linkage (CHeReL) and the Australian Institute of Health and Welfare (AIHW) Data Integration Unit in linking the numerous data collections. They also wish to thank the investigators named on the UNSW Research Infrastructure Scheme grant for their assistance with securing funds to build the ELC platform, and investigators at the Faculty of Medicine and Health, UNSW, for their ongoing management and governance of the ELC platform. The linkage was funded by a UNSW Research Infrastructure Scheme grant and a UNSW Scientia Program Award. UNSW’s National Perinatal Epidemiology and Statistics Unit contributed research support funds for project management. Initial data checking and cleaning was resourced by a National Health and Medical Research Council Ideas grant (#2010778).

## Ethics

The study received approval from the Australian Institute of Health and Welfare Human Research Ethics Committee (EO2020-2-1130) and the NSW Population and Health Services Research Ethics Committee (2019/ETH11830).

