## Supplementary for "Opioid Analgesic Initiation During Pregnancy and the Risk of Adverse Maternal and Neonatal Outcomes: A Target Trial Emulation"

**Appendix:**

**Supplementary Tables:**

**Supplementary Figures:**

Appendix 1: Specification of the hypothetical target trial

The components of the hypothetical target trial are as follows:

*Eligibility criteria*

Eligibility criteria included pregnant women aged 12-60 years who were residents of NSW, had not been dispensed an opioid within the 90 days prior to conception, had not received a cancer diagnosis or undergone cancer treatment within the year prior to conception, and did not have active opioid use disorder (OUD), defined as treated or untreated OUD within the previous 2 years (from conception).

*Treatment strategies*

The treatment strategies in this target trial included initiating opioid analgesic treatment, with individuals in this group referred to as opioid initiators. This group encompassed those initiating any type of opioid analgesic, including combinations with paracetamol or oxycodone with naloxone. The other strategy involved not initiating opioid analgesics, with individuals in this group referred to as non-initiators.

*Treatment assignment*

Eligible women would be randomly assigned to a treatment strategy and would be aware of their assignment.

*Outcomes*

Maternal outcomes that could occur at any time during pregnancy included placental abruption, preterm premature rupture of membranes (PPROM) (membrane rupture <37 weeks gestation), and preterm birth (birth <37 weeks gestation). Maternal outcomes assessed after childbirth included postpartum haemorrhage (>500mL blood loss associated with vaginal delivery; >1000mL blood loss associated with caesarean delivery) and severe maternal morbidity complications, a composite indicator used to measure major maternal morbidities.^1^ Neonatal outcomes included low (<7) Apgar score at 5 minutes after birth, perinatal death (stillbirth or neonatal death [within 28 days of birth]), small for gestational age (SGA) (<10th weight centile based on gestational age), neonatal abstinence syndrome (NAS), and a composite indicator of severe neonatal morbidity complications. Some of the individual outcomes described above, or components thereof, were also part of the composite indicator for severe neonatal morbidity complications, including very preterm birth (gestational age <32 weeks) and preterm birth (Table S2). None of the individual outcomes were included in the severe maternal morbidity complications composite.

*Follow-up period*

For each eligible woman, follow-up began at randomisation and ended at the occurrence of either the outcome of interest, discharge from the maternity ward (for mothers), 28 days after birth (for infants), death, loss to follow up, or December 31, 2019, whichever occurred first.

*Causal contrast of interest*

The causal contrast of interest was the intention-to-treat effect.

*Statistical analysis*

Risk ratios and risk differences for the intention-to-treat effect were estimated by comparing the risk of adverse maternal or neonatal outcomes among women who initiated opioids with those who did not, with adjustment for loss to follow-up.

Appendix 2: TARGET Checklist

| Item no. | | | | Checklist item | | | | | Location reported | |
| --- | --- | --- | --- | --- | --- | --- | --- | --- | --- | --- |
| Abstract | | | | | | | | | | |
| 1 | a | | | | Identify that the study attempts to emulate a target trial using observational data. State the study objectives and briefly summarize the specified target trial. | | | | Reported in the abstract on page 4. | |
|  | b | | | | Report the data sources used for emulation. | | | | Reported in the abstract on page 4. | |
|  | c | | | | Summarize key assumptions, statistical methods, findings and conclusions. | | | | Reported in the abstract on page 4. | |
| Introduction | | | | | | | | | | |
| 2 | Background | | | | Describe the scientific background of the study and the gap in knowledge. | | | | Background and knowledge gap presented on page 7. | |
| 3 | Causal question | | | | Summarize the causal question. | | | | Causal question described on page 7. | |
| 4 | Rationale | | | | Describe the rationale for emulating a target trial with the available data. Cite randomized trials informing the design of the target trial if applicable. | | | | Rationale for emulating the target trial is described on page 7. There were no existing randomised trials to inform the design of this trial. | |
| **Methods** | | | | | | | | | | |
| 5 | Data sources | | Cite the data sources contributing to the analyses and for each one describe the following: original purpose, type, the geographic locations, setting and time-period. If relevant, describe how the data were linked or pooled. | | | | | | The data sources are described on page 7, with additional information described in Appendix 3 of the supplementary. | |
| 6 | Target trial specification  Specify the components of the target trial protocol that would answer the causal question. | | | | | 7 | **Target trial emulation**  Describe how the components of the target trial protocol were emulated with the observational data, including how all variables were measured or ascertained. | | **Location item 6 (specification) reported** | **Location item 7 (emulation) reported** |
|  | Eligibility criteria | | | | |  | Eligibility criteria | | Described in the supplementary in Appendix 1 and appendix Table 1. | Described under the sub-heading ‘Study Population’ on page 8, and in appendix Table 1. |
|  | a | Describe the eligibility criteria. | | | |  | a | Describe how the eligibility criteria were operationalized with the data. |  |  |
|  | **Treatment strategies** | | | | |  | **Treatment strategies** | | Described in the supplementary in Appendix 1 and appendix Table 1. | Described under the sub-heading ‘Exposure’ on page 8-9, and in appendix Table 1. |
|  | b | Describe the treatment strategies that would be compared. | | | |  | b | Describe how the treatment strategies were operationalized with the data. |  |  |
|  | **Assignment procedures** | | | | |  | **Assignment procedures** | | Described in the supplementary in Appendix 1 and appendix table 1. | Described under the sub-heading ‘Exposure’ on page 8-9, Appendix 4, and in appendix table 1. |
|  | c | Report that eligible individuals would be randomly assigned to treatment strategies and may be aware of their treatment allocation. | | | |  | c | Describe how assignment to treatment strategies was operationalized with the data. |  |  |
|  | **Follow-up** | | | | |  | **Follow-up** | | Described in the supplementary in Appendix 1 and appendix table 1. | Described in appendix table 1 |
|  | d | Clarify that follow-up would start at time of assignment to the treatment strategies. Specify when follow-up would end. | | | |  | d | Clarify that follow-up starts at the time individuals were assigned to the treatment strategies. Describe how the end of follow-up was operationalized with the data. |  |  |
|  | **Outcomes** | | | | |  | **Outcomes** | | Described in the supplementary in Appendix 1 and appendix table 1. | Described under sub-heading ‘Outcomes’ on page 1, in appendix table 1 and in appendix table 4. |
|  | e | Describe the outcomes. | | | |  | e | Describe how the outcomes were operationalized with the data. |  |  |
|  | Causal contrasts | | | | |  | Causal contrasts | | Described in the supplementary in Appendix 1 and appendix table 1. | Described under Methods on page 7 and appendix table 1. |
|  | f | Describe the causal contrasts of interest, including effect measures. | | | |  | f | Describe how the causal contrasts were operationalized with the data, including effect measures. |  |  |
|  | **Identifying assumptions** | | | | |  | Identifying assumptions | | Described in the supplementary in Appendix 1 and appendix table 1. | Described under sub-heading ‘Covariates’ and ‘Statistical analysis’ on page 9-10 and appendix table 1. |
|  | g | Describe assumptions that would be made to identify each causal estimand. Describe the variables, if any, related to these assumptions. | | | |  | g.i | For each causal estimand, describe assumptions made to identify it, including assumptions regarding baseline confounding due to lack of randomization. |  |  |
|  |  |  | | | |  | g.ii | Describe how the variables related to these assumptions were operationalized with the data |  | Described under sub-heading ‘Statistical analysis’ on page 9-10, Appendix 5, and appendix table 1. |
|  | **Data analysis plan** | | | | |  | **Data analysis plan** | | Described in the supplementary in Appendix 1 and appendix table 1. | Described under sub-heading ‘Covariates’ and ‘Statistical analysis’ on page 9-10 and appendix table 1. Missing covariate data quantified in appendix table 5. |
|  | h | For each causal estimand, describe the data analysis procedures and any associated statistical modelling assumptions, including approaches for handling missing data. | | | |  | h.i | For each causal estimand, describe the data analysis procedures and any associated statistical modelling assumptions, including approaches for handling missing data. |  |  |
|  |  |  | | | |  | h.ii | For each causal estimand, describe any additional analyses conducted to assess the sensitivity of the results to the choice of operationalizations, assumptions and analysis. |  | Described under sub-heading ‘Subgroup analysis’, ‘Sensitivity analysis’ and ‘Post-hoc analysis’ on page 9-11, appendix table 1, and Appendix 5. |
| **Results** | | | | | | | | | | |
| 8 | Participant selection | | | | Report numbers of individuals assessed for eligibility, eligible, and assigned to each treatment strategy. A flow diagram is strongly recommended. | | | | Described in Results on page 11 and reported in a flow diagram in appendix figure 2. | |
| 9 | Baseline data | | | | Describe the distribution of characteristics of individuals at baseline, by treatment strategy. | | | | Described in Results on page 11, reported in Table 1, and appendix figure 3. | |
| 10 | Follow-up | | | | Summarize length of follow-up and describe reasons for end of follow-up for each treatment strategy and causal contrast. | | | | Described in Results on page 11. | |
| 11 | Missing data | | | | Describe the frequency of missing data in all variables, by treatment strategy when applicable. | | | | Missing covariate data quantified in appendix table 5. | |
| 12 | Outcomes | | | | Describe the frequency or distribution of each outcome, by treatment strategy. | | | | Figures 2-3; appendix figures 9-17. | |
| 13 | Effect estimates | | | | Report the effect estimates for each causal contrast with corresponding measures of precision, including both absolute and relative measures of effect, when applicable. | | | | Described in Results on page 11-13; Figures 2-3; appendix figures 9-17. | |
| 14 | Additional analyses | | | | Report results of all analyses to assess the sensitivity of the estimates to choices in operationalizations, assumptions and analysis. | | | | Described in Results on page 12-13; appendix figures 9-17. | |
| **Discussion** | | | | | | | | | | |
| 15 | Interpretation | | Provide an interpretation of the key findings. | | | | | | Interpretation of key findings are provided in the Discussion on pages 13-14. | |
| 16 | Limitations | | Discuss the limitations of the study considering differences between the target trial and its emulation and the plausibility of assumptions, including assumptions regarding baseline confounding due to lack of randomization. | | | | | | Limitations and confounding due to lack of randomisation described on page 14. | |
| **Other information** | | | | | | | | | | |
| **17** | Ethics | | | | Provide the institutional research board or ethics committee that approved the study and approval numbers, if relevant. | | | | Ethics information provided. | |
| **18** | Registration | | | | State whether, when and where the study protocol was registered. | | | | NA - Protocol was not registered. | |
| **19** | Sharing of study materials | | | | Provide information on whether data, analytic code and/or other materials are accessible, and where and how they can be accessed. | | | | Information on data sharing are reported. | |
| **20** | Funding sources | | | | Provide the sources of funding and detail the role of the funders in the design, conduct and reporting of the study. | | | | Sources and roles of the funders are declared. | |
| **21** | Conflicts of interest | | | | State any conflicts of interest and financial disclosures for all authors. | | | | Conflicts of interest are declared. | |

Appendix 3: Additional data source information

The Early Life Course (ELC) data were created through probabilistic linkage by both the NSW Centre for Health Record Linkage and the Australian Institute of Health and Welfare, using names, date of birth, sex and address to link information for individuals across data collections.^2^ The data separation principle was applied, such that no one had access to both identifying information and content data. The record linkage was completed in mid-2022 and unit record data were supplied to the researchers between November 2022 and May 2023.

The perinatal data are based on the NSW Perinatal Data Collection (PDC), which contains information on mothers and babies collected at the time of birth including medical and obstetric information, delivery information, condition of the infant, and gestational age at birth measured in completed weeks. For each birth, a record is generated in the perinatal data (referred to as ***perinatal record***), one record is generated in the hospital admission data for the mother (referred to as the ***woman’s childbirth admission***), and one record is generated in the hospital admission records for the baby (referred to as the ***baby’s birth admission****).* We defined a delivery episode as all hospital records from admission to the first discharge home for either the mother or baby, including any transfer admissions. Delivery episodes were used to identify several of the maternal and neonatal outcomes (Table S3). Subsidised PBS medicines are available to Australian citizens, permanent residents, and eligible foreign visitors. In Australia, most opioids are prescription-only and listed on the PBS. During the study period, PBS-listed opioids included buprenorphine, codeine, fentanyl, hydromorphone, methadone, morphine, oxycodone, tapentadol (sustained-release formulation only), and tramadol. Three prescription opioids were not PBS-listed: alfentanil and pethidine (available as injection formulations only) and immediate-release tapentadol. Both formulations of tapentadol were approved for marketing in Australia in 2011, and the sustained-release formulation was added to the PBS on 1 June 2014. Low-dose codeine products (≤15 mg) are also not PBS-listed; they were the only opioids available over the counter until their rescheduling to prescription-only status on 1 February 2018, after which they were available as private prescriptions. The hospital admissions database is a statutory data collection containing records of all hospital discharges from NSW public and private hospitals and day procedure centres. Diagnoses are coded according to the International Classification of Diseases, Version 10, Australian Modification (ICD-10-AM)^3^ and procedures coded according to the Australian Classification of Health Interventions (ACHI).^4^ Mortality data included information on deaths obtained from the NSW Register of Births, Deaths and Marriages.

Appendix 4: Creating sequential trials using observational data

To derive the sequential target trials, we used information from the perinatal records to estimate the conception date according to the following equation, conception=date of delivery – gestational age at delivery (days) + 14 days.^5^ To estimate the starting date of each trial, we used 7-day intervals from the conception date (conception = last day of gestational week 2). For example, trial 1 included days 1 - 7 post-conception (gestational week 3), trial 2 included 8 – 14 days post-conception (gestational week 4), and so forth until the date of childbirth.

Appendix 5: Additional statistical analysis information

*Statistical analysis*

We excluded participants who experienced placental abruption or preterm prelabour rupture of membranes (PPROM) before trial entry (>0.1% of participants for each outcome). For severe neonatal morbidity, preterm birth, and PPROM, we included trials from gestational week 3 through 36 completed weeks of gestation, because pregnancies were no longer at risk of these outcomes from 37 completed weeks onwards. For the remaining maternal outcomes, we included all possible trials from gestational week 3 through week 42.

Within each sequential trial, we estimated a trial-specific propensity score using logistic regression, with opioid initiation at trial entry as the dependent variable and measured baseline covariates as independent variables. Baseline covariates included those listed in Supplementary Table 5 and were re-assessed at the start of each trial. These trial-specific propensity scores were then used to generate stabilised inverse probability of treatment weights (IPTWs) separately for each weekly trial, such that treatment groups were balanced on measured baseline characteristics within each trial.

For the target trial emulation, we could not adjust for informative censoring due to loss to follow-up because the available data included only pregnancies with a recorded childbirth. We therefore assumed that opioid initiation did not affect the probability of early pregnancy loss or other forms of loss to follow-up, and/or that there were no common causes of pregnancy loss and the outcomes of interest that were not adequately accounted for. Because pregnancy loss precludes the occurrence or observation of subsequent neonatal outcomes, neonatal outcomes, including neonatal abstinence syndrome (NAS), small for gestational age (SGA), neonatal death, and low Apgar score (<7), were estimated as risks conditional on live birth.

For time-to-event outcomes, we subsequently pooled trial-specific effect estimates using fixed-effect meta-analysis.

*Subgroup analysis*

We investigated the impact of starting opioid treatment based on the trimester during which trial entry occurred (trimester 1; gestational weeks 3-12, trimester 2; weeks 13-27, trimester 3; week 28 onwards), and the type of opioid (codeine, oxycodone, tramadol). For the opioid-type analysis, each woman's first dispensing assigned her to a single opioid type, so she could not initiate, for example, both codeine and oxycodone. We further performed analyses categorised by the total quantity of opioid dispensed at treatment assignment, expressed in oral morphine equivalents (OME), categorised as ≤100 OME, >100 OME and calculated as follows:^6^

OME (mg) total= Product strength (mg) * quantity of opioids dispensed * OME conversion factor of opioid dispensed (according to published metrics)^6^

*Sensitivity analysis*

To assess how robust any identified risks are to unmeasured confounding we performed two additional analyses. Firstly, we restricted the cohort to women who have a history of chronic pain diagnosis (see Table S5) within the year prior to trial entry. Secondly, we re-specified the hypothetical target trial and its observational analogue to focus on women who were clinically indicated to receive opioids by randomly assigning women to receive either codeine or a non-codeine opioid treatment. Codeine is a valid active comparator for opioids given the overlap in most indications (indicated for mild/moderate pain vs moderate/severe pain, respectively) and that it is generally considered safer to take during pregnancy.^7^ For the sensitivity analysis, we repeated the IPTW analyses but excluded covariates with very small or zero cell counts from the propensity score model in each trial to avoid violations of the positivity assumption and model instability. In addition, some variables were collapsed into broader categories to increase cell counts. Covariate balance was assessed for all variables using standardized mean differences (SMDs) before and after weighting. If any covariates remained imbalanced after weighting (SMD >0.1), they were additionally included in the outcome model to provide a doubly robust adjustment.

| **Table 1: Protocol for the hypothetical target trial and its emulation.** | | |
| --- | --- | --- |
| **Components of the target trial and its emulation** | **Target trial** | **Emulations** |
| *Eligibility criteria* | - Enrolment period: July 1, 2013, to March 9, 2019. - Pregnant women aged 12 to 60 years at enrolment. - Resident of NSW. - No cancer diagnosis or treatment within the previous year. - No dispensing of an opioid in the 90 days prior to the start of pregnancy (conception). - No prior opioid use in the current pregnancy. - No active OUD: no history of treated or untreated opioid use disorder within the previous 2 years. - No documented? clinical indication is required. | Same as target trial and additionally:   - No missing or implausible gestational age. - No missing maternal age. - No births that occurred in the one hospital not available for linkage.   Departures from the target trial:   - The perinatal database includes complete pregnancies only, i.e., those that result in live or still birth to those that last at least 20 weeks gestation or 400g birth weight. - NSW residence was ascertained at the time of birth. |
| *Treatment strategies* | Eligible women are then randomised to initiate one of two treatment strategies at the time of trial enrolment:   (1) Opioid analgesic initiators: any type of opioids (including opioids received in combination with for example, acetaminophen, or naloxone) (any dose). (2) Non-initiators: no opioid treatment. | Same but eligible women at the start of each trial (each gestational week) were classified to treatment arms based on the observed data.  This was ascertained through dispensing records from the Pharmaceutical Benefits Scheme data. |
| *Treatment assignment* | Patients are randomised to either treatment strategy at enrolment and are *aware* of the strategy they are assigned to. | Same but randomisation was emulated using stabilised inverse probability of treatment weighting based on the propensity score, estimated from baseline and post-baseline (chronic morbidities) covariates. |
| *Outcomes* | Maternal outcomes include:   - Preterm birth - PPROM - Placental abruption - Postpartum haemorrhage - Composite endpoint of severe maternal complications   Neonatal outcomes include:   - Low (<7) Apgar score at 5 minutes - Perinatal death (stillbirth or neonatal death) - Composite endpoints of severe neonatal morbidity complications - NAS - SGA | Same.  Diagnoses were identified with code or algorithms in perinatal, hospital, or mortality databases. |
| *Follow-up* | Follow-up begins when an individual meets all eligibility criteria:   - When an individual is randomly assigned to one of the treatment strategies.   Maternal outcomes:  Follow-up ends for women at the earliest of:   - Outcome of interest - Death - Loss to follow-up (moved state/country or pregnancy ends < 20 weeks) - Administrative end of follow-up (until discharge after birth or 31/12/2019)   Neonatal outcomes:  Follow-up ends for infants at the earliest of:   - Outcome of interest - Death - Loss to follow-up (moved state/country) - Administrative end of follow-up (until 28 days after birth or 31/12/2019) | Same.  Follow-up begun at the time of dispensing for those assigned to opioid treatment and midweek for those assigned to no opioid treatment and ends at the administrative end of follow-up.    We could not ascertain loss to follow-up because women appear in the perinatal database at the end of pregnancy (i.e., during delivery) which forces a “complete case” approach. |
| *Causal contrast of interest* | We estimate the intention-to-treat effect of receiving opioid analgesics versus *no treatment*. | Observational analogue of intention-to-treat. |
| *Analysis plan* | The causal contrast of interest is the intention-to-treat effect of comparing risks for each outcome of interest among the exposed group in comparison to the reference group, with adjustment for loss to follow-up.  Neonatal outcomes including NAS, SGA, neonatal death, and low Apgar score (<7) at 5 minutes will be reported as conditional probabilities along with the risk of livebirth.  The remaining neonatal outcomes will be examined in both live and stillbirths. | Same – the target trial was sequentially emulated during each gestational week of pregnancy in the database.    However, we did not adjust for informative censoring (loss to follow up) given that we have a complete case analysis. We assumed that treatment does not affect the risk of pregnancy loss or other censoring, and/or there is no common cause of pregnancy loss and the outcomes of interest. |
| Abbreviations: NAS, neonatal abstinence syndrome; OUD, opioid use disorder; PPROM, Preterm premature rupture of membranes; SGA, small for gestational age. | | |

Table 2: Eligibility Criteria: Exclusion definitions

| **Exclusion criteria** | **Timeframe** | **Data collection** | **Relevant Codes** |
| --- | --- | --- | --- |
| Missing maternal age or age <12 or >60 | Ascertained at the time of birth | Perinatal records, supplemented from the NSW Registry of Births, Deaths and Marriages, and hospital records. |  |
| Non-NSW residence | Ascertained at the time of birth | Perinatal records |  |
| Prescription opioids | 90 days prior to conception | Medicine dispensing records | **ATC codes**:  N02A, R05DA04, N02BE51, N02BA51 |
|  | Prior use during current pregnancy |  |  |
| Cancer | 1 year prior to trial entry | Hospital records | **ICD-10-AM codes:**  C00-C97, Z51.1 |
|  |  | Medicine dispensing records | **ATC codes**  L01AA01-L01AX04, L01BA01,^a^  L01BA03- L01XX53, L02BA01,^b^  L02BG03, L02BG04, L02BG06,  L02BB01-L02BB04, L02BX01-L02BX03,  L04AX02, L04AX04, L04AX06,  L02AE02,^c^ L02AE03^d^ |
| Treated or untreated OUD | 2 years prior to trial entry | Hospital | **ICD-10-AM codes:**  F11.1, F11.2, T40.0, T40.1, T40.2, T40.2 |
|  |  | OAT records | Presence of authority for prescription of methadone or buprenorphine for OUD treatment |
| One hospital not available for linkage | Ascertained at the time of birth | Perinatal records | Hospital code: B259, B359 |
| Missing or implausible gestational age |  |  | Gestational age < 20 weeks |
| Abbreviations: NSW, New South Wales; OAT, opioid agonist therapy; OUD, opioid use disorder. | | | |

^a^ Excludes PBS item codes (01622J, 01623K, 02272N)
^b^ Excludes PBS item code (10911G)
^c^ Excludes PBS item codes (10255R, 10256T, 11944P, 11960L)
^d^ Excludes PBS item code (01454M)

Table 3: Opioids included in the study and their corresponding Anatomical Therapeutic Classification (ATC) codes.

| **Medicine Name** | **ATC Code** |
| --- | --- |
| Codeine | R05DA04, N02AA |
| Codeine combination preparations | N02AA59, N02AJ06, N02AJ07, N02BA51, N02BE51 |
| Buprenorphine* | N02AE01 |
| Fentanyl | N02AB03 |
| Hydromorphone | N02AA03 |
| Methadone* | N02AC |
| Morphine | N02AA01 |
| Oxycodone | N02AA05 |
| Oxycodone with naloxone | N02AA55 |
| Tapentadol | N02AX06 |
| Tramadol | N02AX02 |

*****During the study period, PBS-listed buprenorphine and methadone were available only for pain management, and prescriptions of buprenorphine or methadone for the treatment of opioid use disorder were not captured in these data.

| Table 4: Outcome ascertainment. | | | | |
| --- | --- | --- | --- | --- |
| Outcome | Data collection & definition | ICD-10-AM Codes | ACHI procedure Codes | Citation or validation results |
| Maternal outcomes |  |  |  |  |
| Placental abruption | Hospital records when gestational age was ≥20 weeks. | O45 |  | Validated during a women’s delivery admission: PPV: 100% (95% CI: 27.6% - 100%).^8^ |
| Preterm premature rupture of membranes | Hospital records when gestational age at birth was <37 weeks. | O42 |  |  |
| Preterm birth | Preterm birth is characterized by childbirth occurring before 37 completed weeks, either as a result of spontaneous onset of labour or medical intervention and is determined based on the gestational age at delivery and the onset of labour (recorded in the perinatal record). |  |  |  |
| Postpartum haemorrhage (>500 mL for vaginal, >1000 mL for caesarean delivery) | Identified from a recorded postpartum haemorrhage in the perinatal records or a relevant ICD-10 diagnosis code (during the delivery hospital admission. | O72.0, O72.1 |  | Validated during a women’s delivery admission: PPV: 83.9% (95% CI: 73.7% - 91.4%).^8^ |
| Severe maternal morbidity complications | Mother’s hospital admission for childbirth | Acute abdomen (K35, K37, K65.0, K65.9, N73.3, N73.5); Acute renal failure (O90.4, N17, N19, N99.0); Acute psychosis (F53.1, F23); Cardiac arrest, failure, or infarction (O89.1, O74.2, O90.3, I21, I42, I43, I46, I50, J81); Cerebral oedema or coma (G93.6, R40.2); Disseminated intravascular coagulopathy (D65); Cerebro-vascular accident (I60–I64); Major complications of anaesthesia (O74.0, O74.2, O74.3, O89.0–O89.2, O29.0–O29.2); Obstetric embolism (O88); Shock (R57, O75.1, T80.5, T88.6); Sickle cell anaemia with crisis (D57.0); Status asthmaticus (J46); Status epilepticus (G41); Uterine rupture (O71.0, O71.1). | Assisted ventilation including tracheostomy (13857-00, 13879-00, 13882-00, 13882-01, 13882-02, 92038-00, 92039-00, 92040-00, 92041-00, 41880-00, 41883-00, 41883-01, 90179-06, 92046-00, 92047-00); Curettage in combination with a general anaesthetic (16564-00–01, 35640-00–03, 92514-10–99, 92502-00–03); Dialysis (13100, 13112-00, 90351-00, 13109-00, 13109-01, 13110-00); Evacuation of haematoma (90484-00, 90484-01, 90484-02, 30394-00); Hysterectomy (35653-00 to 03); Procedures to reduce blood flow to uterus (34103-12, 33833-03, 35321-00, 33845-00, 30385-00, 35759-00); Reclosure of disrupted CS wound (30403-03); Repair of bladder or cystostomy (90480-00, 37004-03, 37004-02, 37011-00, 37008-01); Repair of intestine (30566-00, 30375-19, 30565-00, 32069-00, 30375-24, 32003-00, 32000-00, 32003-01, 32000-01, 32005-01, 32004-01, 32006-00, 32006-01, 32005-00, 32004-00, 32012-00, 32009-00, 30375-25, 43816-02); Repair ruptured or inverted uterus (90478-00, 16570-01); Transfusion of blood or coagulation factors (13706-01, 13706-02, 13706-03, 92061-00, 92062-00, 92206-00) | Validated.^9^ |
| **Neonatal outcomes** |  |  |  |  |
| Severe neonatal morbidity complications | Obtained from perinatal records, baby’s birth admission or in any hospital transfer admission that occurred prior to the first discharge home, and mortality records. | Gestational age<32 weeks, birthweight<1500g, perinatal death, respiratory distress syndrome (P22.0), Seizure (P90, R56); Intraventricular haemorrhage (grades 2, 3 and 4) (P52.1, P52.2); Cerebral infarction (I63); Periventricular leukomalacia (P91.2); Birth trauma (intracranial haemorrhage, paralysis due to brachial plexus injury, skull, or long bone fracture) (P10.0 – P10.3, P13.0, P13.2, P13.3, P14.0, P14.1); Hypoxic ischemic encephalopathy (P91.5, P91.81, P91.6); Necrotising enterocolitis (P77); Broncho-pulmonary dysplasia (P27.1); Sepsis/septicaemia (streptococcus, staphylococcus, E. coli, unspecified Gram-negative) (P36, A40, A41.5, A41.9, B95.1, B96.2); Pneumonia (P23, J12 to J18); Other respiratory: primary atelectasis, respiratory failure (P28.0, P28.5). | Resuscitation (92052, 92053, 92042–00, 90225); Ventilatory support (mechanical ventilation and/or CPAP) (13882, 13857–00, 13879–00, 22007, 90179, 92038, 92039); Central venous or arterial catheter (38206, 13303–00, 34524–00, 34530–01, 13300–00, 13300–02, 13319–00, 13815); Transfusion of blood or blood products (13706–01 to 04, 92206–00, 13306–00); Pneumothorax requiring an intercostal catheter (38409–00); Any body cavity surgical procedure (30373, 30375, 30378–00, 30562, 30564 to 30566, 30571, 30601, 30615–00, 30617–00, 32123–00, 36516, 36537, 36564, 36579, 38403–00, 38600–00, all codes start with 387, 39015, 39640–00, 40003, 40100–00, 40103–00, 41883, 43801–00, 43807–00, 43816–02, 43837, 43843, 43852–00, 43864, 43867, 43870–00, 43873, 43876, 43900–00, 43915–00, 43930–00, 43945–00, 43963–00, 43978, 90180, 90224–00); Any intravenous fluids (96199). | Validated.^1^ |
| Perinatal death | Stillbirth or neonatal death.  Stillbirth: defined the death of an unborn baby who is over 400 grams of birthweight or 20 weeks of gestation and identified from the perinatal data.  Neonatal death: defined as the death of a liveborn baby within 28 days from date of birth. Neonatal deaths were obtained from mortality data. |  |  |  |
| Apgar score (<7) at 5 minutes | Obtained from the perinatal records.  Clinical indicators of a baby’s condition shortly after birth obtained from the perinatal record. A score of less than 7 indicates complications for the baby. |  |  |  |
| Small for gestational age (<10th centile) | Obtained from the perinatal records:  SGA was defined as birthweight of less than the tenth percentile specific for sex and gestational age according to Australian national birthweight percentiles in singleton pregnancies.^10^ |  |  |  |
| Neonatal abstinence syndrome | Diagnosis was recorded in women’s hospital admission for childbirth, baby’s birth admission or any infant hospital admission within 28 days of childbirth. | P96.1 |  |  |

Table 5: List of covariates, relevant databases, times and codes used for their identification.

| **Covariate** | **Data collection & ascertainment time** | **Relevant codes** |
| --- | --- | --- |
| **Maternal Characteristics** | | |
| Year of trial entry | Perinatal registry:  Gestational age and date of birth, recorded at delivery, were used to determine the year of trial entry. |  |
| Maternal age at trial entry  (Categorical: <25, 25-34, ≥35) | Perinatal registry:  Maternal date of birth recorded at the time of childbirth.  Supplemented with NSW Registry of Births, Deaths and Marriages, hospital records and emergency department records. |  |
| Country of birth  (Dichotomous: Australia-born, overseas-born) | Perinatal registry:  Recorded at the time of childbirth and supplemented with hospital records. |  |
| Partner status  (Dichotomous: does not have a partner/has a partner) | Hospital records:  Recorded at the time of childbirth and supplemented with social security records. | Missing partner status was imputed as the most common category (“has a partner”); this applied to 20,534 eligible person-trials (0.1% of the total). |
| Social security beneficiary  (Dichotomous: No/Yes) | Social security records:  Records from the year prior to treatment assignment until treatment assignment. |  |
| Residential socio-economic disadvantage quintile  (Categorical: Quintile 1/2/3/4/5) | Perinatal registry:  Derived from Statistical Area Level 2 (SA2) variable (recorded at the time of childbirth) and supplemented with hospital records. | Missing socioeconomic status was imputed as the median category (quintile 3); this applied to 5,441 eligible person-trials (0.0% of the total). |
| Residential remoteness (Categorical: Major cities/inner regional/outer regional/remote and very remote) | Perinatal registry:  Derived from SA2 variable (recorded at the time of childbirth) and supplemented with hospital records. | Missing remoteness status was imputed as the most common category (“major cities”); this applied to 409,323 eligible person-trials (2.2% of the total). |
| **Obstetric and health-related characteristics (Recorded at delivery)** | | |
| Parity  (Categorical: 0, 1, 2, ≥3) | Perinatal registry:  Recorded at the time of childbirth. | Missing previous pregnancy status was imputed as the most common category (category 0: “no previous pregnancy”); this applied to 21,146 eligible person-trials (0.1% of the total). |
| Previous caesarean section  (Dichotomous: No/Yes) | Perinatal registry:  Recorded at the time of childbirth. | Missing information was imputed using the most common category (“no previous caesarean section”); this applied to 271,903 eligible person-trials (1.5% of the total). |
| Pregnancy interval  (Categorical: No previous birth, 0 - <6 months, 6 - 18months, 18 – 33 months, >33months) | Perinatal registry:  Calculated as conception date – prior date of childbirth – 7. |  |
| Gestational age at the time of trial entry  (Continuous) |  |  |
| Multifetal gestation  (Dichotomous: No/Yes) | Perinatal registry:  Recorded at the time of childbirth. |  |
| Composite indicator -previous adverse pregnancy outcome (Dichotomous: No/Yes) |  | See appendix Table 4. |
| Composite indicator -previous adverse infant outcome (Dichotomous: No/Yes) |  | See appendix Table 4. |
| **Maternal Morbidities** | | |
| Anaemia and coagulation  (Dichotomous: No/Yes) | Hospital records:  Records from the year prior treatment assignment until delivery. | D50-D53, D55-D68 |
|  | Medicine dispensing records:  Records from the year prior to treatment assignment until delivery. | Anticoagulants: B01AA03 - B01AB06, B01AE07, B01AF01, B01AF02, B01AX05  Antiplatelets: B01AC04 - B01AC07, B01AC12 - B01AC30, PBS items (05030R, 05035B, 05042J, 10111E, 10117L, 10129D, 10130E, 05751Q, 06456T, 11065J, 11082G, 11069N, 1109Q) |
| Cardiovascular disease  (Dichotomous: No/Yes) | Hospital records:  Records from the year prior to treatment assignment until delivery. | Records from the year prior to trial entry until delivery: I05-I09, I34-I39, I50, I20, I25, I27, I28, Q20-Q25, O99.4, U82.1, U82.2  Records from the year prior to trial entry until trial entry: I00-I02, I21-I24, I26, I30-I33, I40-I43, I44-I49, I51-I52, I60-I64, G45.8, G45.9, I65, I66, I67.2, I70, I73, I74, I77, U82.1, U82.2 |
|  | Medicine dispensing records:  Records from the year prior to treatment assignment until delivery. | Congestive heart failure: C01EB17, C09DX04, C03DA02 - C03DA99, C07AB07, C07AB12, C07AG02, [(C03CA01 - C03CC01) and (C09AA01- C09AX99 or C09CA01 - C09CX99)], PBS items (08732N, 08733P, 08734Q, 08735R) ^‡^  Ischaemic heart disease-hypertension: C07AA01 - C07AA06, C07AA08 - C07AB01, [C07AB02 if PBS item code is not (08732N, 08733P, 08734Q, 08735R) ^‡^], C07AB03, C07AG01, C08CA01 - C08DB01, C09BB02 - C09BB10, C09DB01 - C09DB04, C09DX01, C09DX03, C10BX03*  Ischaemic heart disease-angina: C01DA02-C01DA14, C01DX16, C08EX02 |
| Chronic liver failure  (Dichotomous: No/Yes) | Hospital records:  Records from the year prior to treatment assignment until delivery | B18, K70.0 - K70.3, K70.9, K71.3 - K71.5, K71.7, K73, K74, K76.0, K76.2 - K76.4, K76.8, K76.9, Z94.4 |
| Chronic renal disease  (Dichotomous: No/Yes) | Hospital records: | Records from the year prior to trial entry until delivery: N02-N08, N1-N12, N14-N16, N18-N19, N25-N28, Q60-Q63, N39.1, N39.2, T82.4, T86.1, Z49, Z94.0, Z99.2  Records from the year prior to trial entry until trial entry: N00, N01, N17 |
|  | Medicine dispensing records:  Records from the year prior to treatment assignment until delivery | A11CC04 (PBS authority code 5401), B03XA01 – B03XA08, V03AE02, V03AE03, V03AE05 |
| Drug and alcohol disorder  (Dichotomous: No/Yes) | Hospital records:  Records from the year prior to treatment assignment until delivery. | F10-F19, Z50.2, Z50.3, Z72.1, Z72.2 |
|  | Medicine dispensing records:  Records from the year prior to treatment assignment until delivery. | Alcohol dependency: N07BB01 - N07BB99  Smoking cessation pharmacotherapies: N07BA01-N07BA02 |
| Epilepsy  (Dichotomous: No/Yes) | Hospital records:  Records from the year prior to treatment assignment until treatment assignment. | G40, F80.3 |
|  | Medicine dispensing records:  Records from the year prior to treatment assignment until treatment assignment. | N03AA01-N03AD51, N03AF01-N03AX11, N03AX13-N03AX30 |
| Gastro-oesophageal reflux disease  (Dichotomous: No/Yes) | Hospital records:  Records from the year prior treatment assignment until treatment assignment. | K21.0, K21.9 |
|  | Medicine dispensing records:  Records from the year prior to treatment assignment until treatment assignment. | A02BA01 - A02BX05 |
| Infertility  (Dichotomous: No/Yes) | Hospital records:  Records from the year prior to treatment assignment until delivery. | N97 |
| Mental health  (Dichotomous: No/Yes) | Hospital records:  Records from the year prior to treatment assignment until treatment assignment. | F20-F25, F28-F34, F38- F41, F44, F48, O99.5, U79.2, U79.3 |
|  | Medicine dispensing records:  Records from the year prior treatment assignment until treatment assignment. | Anxiety: N05BA01 - N05BA12, N05BE01  Bipolar: N05AN01*  Depression: N06AA01-N06AG02, N06AX03 - N06AX11, N06AX13 - N06AX18, N06AX21 - N06AX26  Psychotic illness: N05AA01 - N05AB02, N05AB06 - N05AL07, N05AX07 - N05AX13 |
|  | Health service use:  Records from the year prior to treatment assignment until treatment assignment. | MBS item codes for mental health treatment plans prepared and reviewed by general practitioners, psychiatrist services, psychologist services, allied health provider services, paediatrician mental health services. |
| Pre-existing diabetes  (Dichotomous: No/Yes) | Hospital records:  Records from the year prior to treatment assignment until delivery. | E10, E11, E13, E14, O24.0-O24.3 |
|  | Medicine dispensing records:  Records from the year prior to treatment assignment until delivery. | A10A, A10BB – A10BK |
|  | Perinatal data:  Recorded at the time of childbirth. |  |
| Gestational diabetes  (Dichotomous: No/Yes)  Applied to trials beginning after gestational week 25. | Hospital records:  Records from conception until delivery. | O24.4, O24.9 |
|  | Perinatal data:  Recorded at the time of childbirth. |  |
| Pre-existing hypertension  (Dichotomous: No/Yes) | Hospital records:  Records from the year prior to treatment assignment until delivery. | I10-I15, O10, U82.3 |
|  | Medicine dispensing records:  Records from the year prior to treatment assignment until delivery. | C03AA01–C03BA11, C03DB01, C03DB99, C03EA01, C09BA02–C09BA09, C09DA01–C09DA08, C02AB01–C02AC05, C02DB02–C02DB99 (C03AA01–C03AX99 or C09CA01–C09CX99) |
|  | Perinatal data:  Recorded at the time of childbirth. |  |
| Gestational hypertension  (Dichotomous: No/Yes)  Applied to trials beginning after gestational week 25. | Hospital records:  Records from conception until delivery. | O13, O16 |
| Obesity  (Dichotomous: No/Yes) | Hospital records:  Records from the year prior to treatment assignment until delivery. | E66.0, E66.01, E66.09, E66.1, E66.2, E66.3, E66.8, E66.9, U78.1 |
| Respiratory disease  (asthma and chronic obstructive pulmonary disease (COPD))  (Dichotomous: No/Yes) | Hospital records: | Records from the year prior to trial entry until delivery.  J40-J44 (COPD)  Records from the year prior to trial entry until trial entry: J45, J46, U83.3 (Asthma) |
|  | Medicine dispensing records: | Records from the year prior to trial entry until delivery: R03AC18, R03AL03, R03AL04, R03AL05, R03AL06, R03BB04, R03BB05, R03BB06, R03BB07, PBS item codes 10018G, 10199T, 8432T, 8519J, 8750M (COPD)  Records from the year prior to trial entry until trial entry: R03AK, R03BA, R03DC (Asthma) |
| Smoking  (Dichotomous: No/Yes)  If smoking was recorded during the first half of pregnancy or in hospital records, it applied to trials up to gestational week 20. If smoking was recorded during the second half of pregnancy or in hospital records, it applied to trials after gestational week 20. | Perinatal data:  Recorded during the first half of pregnancy (conception up until gestational week 20) and during the second half of pregnancy (≥20 gestational weeks). |  |
|  | Hospital records:  Recorded during pregnancy until the time of delivery. | Z72.0 |
| Thyroid disease  (Dichotomous: No/Yes) | Hospital records:  Records from the year prior to treatment assignment until delivery. | E00-E07, E89.0 |
|  | Medicine dispensing records:  Records from the year prior to treatment assignment until delivery. | Hyperthyroidism: H03BA02 - H03BB01  Hypothyroidism: H03AA01 - H03AA02 |
| **Conditions associated with pain** | | |
| Abdominal and back pain  (Dichotomous: No/Yes) | Hospital records:  Records from the year prior to treatment assignment until treatment assignment. | M54, R10 (principal diagnosis code) |
| Chronic viral Hepatitis/HIV  (Dichotomous: No/Yes) | Hospital records:  Records from the year prior to treatment assignment until delivery. | B18.2, B20-B24, B94.2, Z22.5 |
|  | Medicine dispensing records:  Records from the year prior to treatment assignment until delivery. | J05AE01-J05AE12, J05AE14, J05AF01-J05AG05, J05AR01-J05AR99, J05AX07- J05AX09, J05AX12, J05AX14, J05AX15, J05AX65, |
| Musculoskeletal pain  (Dichotomous: No/Yes) | Hospital records:  Records from the year prior to treatment assignment until treatment assignment. | M07, M13, M15.0, M15.1, M15.2, M15.4, M15.8, M15.9, M40.0, M40.2, M40.3, M40.4, M40.5, M41, M42, M43, M45, M46.0, M46.1, M46.2, M47, M48.0, M48.1, M48.2, M48.5, M48.8, M48.9, G95.0, G95.1 (Joint and spinal disease)  M79.7 (Fibromyalgia) |
| Rheumatic disease  (Dichotomous: No/Yes) | Hospital records:  Records from the year prior to treatment assignment until delivery. | D89.1, M05, M06.1, M30.0, M30.1, M30.8, M31.3-M31.7, M32, M33.0-M33.9, M34, M35.0, M35.2, M35.3, M34, M94.1 |
|  | Medicine dispensing records:  Records from the year prior to treatment assignment until delivery. | A07EC, L01BA (item codes 01622J, 01623K, 2272N), L04AA, L04AX, M01C, P01BA, |
| Surgery  (Dichotomous: No/Yes) | Hospital records:  Records from the year prior to treatment assignment until treatment assignment. | Based on the code from Gong et al. applied to procedural block.^11^ |
| Trauma  (Dichotomous: No/Yes) | Hospital records:  Records from the year prior to treatment assignment until treatment assignment. | S00 – T14 in the principal diagnosis code |
| **Medicine use** | | |
| Metformin  (Dichotomous: No/Yes) | Medicine dispensing records:  Records from the year prior to treatment assignment until treatment assignment. | A10BA |
| Non-opioid analgesics  (Dichotomous: No/Yes) | Medicine dispensing records:  Records from the year prior to treatment assignment until treatment assignment. | M01AB01 – M01AH06 (non-steroidal anti-inflammatory drugs [NSAIDS], N02BE01 (paracetamol), N02BG (pregabalin), N02CX01, N02BG PBS items codes 04591P, 04592Q, 04593R, 04594T, 04595W (gabapentin). |
| Psychotropics | Medicine dispensing records:  Records from the year prior to treatment assignment until treatment assignment. | N03A, N05A, N05B, N05C, N06A, N02AX16, N05BA12, N05BA01, N05BA04, N05CD02, N05CD07. |
| Opioids  (Dichotomous: No/Yes) | Medicine dispensing records:  Records from the year prior to treatment assignment until treatment assignment. | See appendix Table 3. |
| Pregnancy Category D/X medicines (suspected teratogens, medicines with other adverse effects)  (Dichotomous: No/Yes) | Medicine dispensing records:  Records from the year prior to treatment assignment until treatment assignment. | Based on published ATC and item number codes.^12^ |
| Systemic corticosteroids | Medicine dispensing records:  Records from the year prior treatment assignment until treatment assignment. | H02AB01-H02AB10 |
| **Healthcare utilisation** | | |
| Number of hospital admissions  (Categorical: 0, 1, ≥2) | Hospital records:  Records from the year prior to treatment assignment until treatment assignment. |  |
| Number of emergency hospital contacts  (Categorical: 0, 1, ≥2) | Emergency presentation records:  Records from the year prior treatment assignment until treatment assignment. |  |
| Number of GP visits  (Categorical: 0, 1, ≥2) | MBS records:  Records from the year prior to treatment assignment until treatment assignment. | MBS category: item numbers  A1 GP: 3, 23, 36, 44 (in surgery visits), 4, 24, 37, 47 (Home/institution/hospital visits).  A5 Prolonged: 160.  A11 After hours: 585, 599.  A22 GP after-hours attendances to which no other item applies: 5000, 5003, 5010, 5020, 5023, 5028, 5040, 5043, 5049, 5060, 5063, 5067.  A30: Medical practitioner video conferencing consultation: 2100, 2122, 2126, 2137, 2143, 2147, 2195, 2199, 2125, 2138, 2179, 2220 |

Abbreviations: ATC, Anatomical Therapeutic Chemical Classification System; GP, General Practitioner; HIV, Human Immunodeficiency Virus; ICD-10-AM, International Classification of Diseases, 10th Revision, Australian Modification; MBS, Medicare Benefits Schedule.

Table 6: Number and percentage of opioid initiators by type of opioid initiated,

| **Opioid type** | **Number (Percent)** |
| --- | --- |
| Any opioid | 26,984 (100.0) |
| Codeine (including codeine in combination with paracetamol) | 23,402 (86.7) |
| Oxycodone (including oxycodone in combination with naloxone) | 3,111 (11.5) |
| Tramadol | 415 (1.5) |
| Tapentadol | 18 (0.07) |
| Fentanyl | 14 (0.05) |
| Buprenorphine | 10 (0.04) |
| Morphine | * (*) |
| Hydromorphone | ≤5 (*) |

*Data suppressed to prevent back calculation of cells with small count (≤5)

Table 7: Number of pregnant women initiating opioids according to the gestational week of pregnancy for any opioid and the three most commonly used opioids, codeine, oxycodone and tramadol.

| **Gestational week** | **No opioid initiation** | **Type of opioid initiated** | | | |
| --- | --- | --- | --- | --- | --- |
|  |  | **Any opioid** | **Codeine** | **Oxycodone** | **Tramadol** |
| **3** | 510126 | 909 | 726 | 128 | 49 |
| **4** | 509329 | 806 | 629 | 124 | 47 |
| **5** | 508635 | 694 | 542 | 120 | 28 |
| **6** | 508027 | 615 | 515 | 90 | 10 |
| **7** | 507466 | 558 | 472 | 64 | 19 |
| **8** | 506901 | 559 | 483 | 68 | 7 |
| **9** | 506333 | 569 | 491 | 67 | 10 |
| **10** | 505740 | 586 | 517 | 60 | 8 |
| **11** | 505190 | 558 | 496 | 59 | ≤5 |
| **12** | 504600 | 597 | 533 | 59 | ≤5 |
| **13** | 503937 | 665 | 582 | 76 | 6 |
| **14** | 503163 | 774 | 665 | 105 | ≤5 |
| **15** | 502419 | 743 | 654 | 75 | 9 |
| **16** | 501725 | 703 | 610 | 89 | ≤5 |
| **17** | 500973 | 742 | 638 | 91 | 10 |
| **18** | 500277 | 699 | 600 | 92 | 6 |
| **19** | 499588 | 694 | 587 | 103 | ≤5 |
| **20** | 499004 | 587 | 509 | 72 | ≤5 |
| **21** | 497956 | 607 | 527 | 77 | ≤5 |
| **22** | 496870 | 622 | 539 | 76 | 6 |
| **23** | 495859 | 596 | 518 | 71 | 6 |
| **24** | 494956 | 585 | 507 | 72 | ≤5 |
| **25** | 493985 | 622 | 548 | 69 | ≤5 |
| **26** | 492969 | 657 | 571 | 77 | 7 |
| **27** | 491961 | 603 | 530 | 66 | ≤5 |
| **28** | 490896 | 632 | 559 | 62 | 11 |
| **29** | 489726 | 622 | 538 | 71 | 11 |
| **30** | 488480 | 690 | 618 | 57 | 14 |
| **31** | 487089 | 678 | 593 | 70 | 14 |
| **32** | 485398 | 753 | 669 | 74 | 10 |
| **33** | 483302 | 784 | 674 | 92 | 16 |
| **34** | 480448 | 831 | 717 | 104 | 8 |
| **35** | 475931 | 939 | 819 | 104 | 13 |
| **36** | 468950 | 927 | 798 | 116 | 13 |
| **37** | 455151 | 1004 | 868 | 116 | 18 |
| **38** | 418633 | 1037 | 918 | 109 | 10 |
| **39** | 325184 | 852 | 798 | 48 | 6 |
| **40** | 175052 | 631 | 601 | 28 | ≤5 |
| **41** | 59952 | 253 | 240 | * | ≤5 |
| **42** | 3561 | 12 | 9 | ≤5 | ≤5 |
| **Total** | 18330000 | 26979 | 23403 | 3111 | 415 |

*Data suppressed to prevent back calculation of cells with small count (≤5)

Table 8: Characteristics and absolute standardised mean differences of opioid analgesic initiators and non-initiators before and after weighting in a sensitivity analysis restricting the cohort to women with a history of chronic conditions associated with pain.

| **Characteristic** | **Person-trials before Weighting**  (n = 446,398) | | |  | **After Weighting** |
| --- | --- | --- | --- | --- | --- |
|  | **Non-initiator person-trials** (n=444,713) | **Opioid initiator person-trials** (n=1,685) | **Absolute std diff** |  | **Absolute std diff** |
| ***Trial characteristics*** | | | | | |
| Year trial started^a^ |  |  |  |  |  |
| 2013-2015 | 164,287 (36.9%) | 623 (37.0%) | 0.00 |  | 0.02 |
| 2016-2017 | 160,853 (36.2%) | 613 (36.4%) | 0.00 |  | 0.02 |
| 2018-2019 | 119,573 (26.9%) | 449 (26.6%) | 0.01 |  | 0.01 |
| ***Maternal Characteristics*** |  |  |  |  |  |
| Maternal age at trial entry (years)^a^ |  |  |  |  |  |
| <25 | 59,417 (13.4%) | 371 (22.0%) | 0.23 |  | 0.06 |
| 25-34 | 265,086 (59.6%) | 963 (57.2%) | 0.05 |  | 0.04 |
| ≥35 | 120,210 (27.0%) | 351 (20.8%) | 0.15 |  | 0.01 |
| Country of Birth^a^ |  |  |  |  |  |
| Australian-born | 280,198 (63.0%) | 1,310 (77.7%) | 0.33 |  | 0.00 |
| Partner status^a^ |  |  |  |  |  |
| Has a partner | 368,676 (82.9%) | 1,240 (73.6%) | 0.23 |  | 0.03 |
| Concessional beneficiary | 131,781 (29.6%) | 789 (46.8%) | 0.36 |  | 0.02 |
| Quintiles of residential socio-economic disadvantage^a^ |  |  |  |  |  |
| Most disadvantaged | 117,217 (26.4%) | 545 (32.3%) |  |  |  |
| Quintiles 2/3/4/5 (Least disadvantaged) | 327,496 (73.6%) | 1,140 (67.7%) | 0.13 |  | 0.06 |
| Residential remoteness^a^ |  |  |  |  |  |
| Major cities | 358,032 (80.5%) | 1,213 (72.0%) | 0.20 |  | 0.03 |
| Inner regional/Outer regional/Remote/Very remote | 86,681 (19.5%) | 472 (28.0%) |  |  |  |
| ***Pregnancy and maternal-related characteristics*** | | | | | |
| Parity^a^ |  |  |  |  |  |
| No previous birth | 185,976 (41.8%) | 588 (34.9%) |  |  |  |
| Any previous birth | 258,737 (58.2%) | 1,097 (65.1%) | 0.14 |  | 0.02 |
| Previous caesarean section^a^ | 90,211 (20.3%) | 369 (21.9%) | 0.04 |  | 0.06 |
| Pregnancy interval |  |  |  |  |  |
| No previous birth | 211,068 (47.5%) | 684 (40.6%) | 0.14 |  | 0.01 |
| <6 - <33 months | 68,805 (15.5%) | 305 (18.1%) | 0.07 |  | 0.09 |
| ≥33 months | 164,840 (37.1%) | 696 (41.3%) | 0.09 |  | 0.06 |
| Multifetal gestation^a^ | 7,792 (1.8%) | 31 (1.8%) | 0.01 |  | 0.08 |
| Previous adverse maternal or neonatal outcome | 109,438 (24.6%) | 488 (29.0%) | 0.10 |  | 0.04 |
| ***Maternal morbidities*** | | | | | |
| Anaemia and coagulation^b^ | 58,743 (13.2%) | 251 (14.9%) | 0.05 |  | 0.03 |
| Cardiovascular disease^b^ | 32,169 (7.2%) | 119 (7.1%) | 0.01 |  | 0.02 |
| Chronic liver failure^b,c^ |  |  |  |  |  |
| Chronic renal disease^b,c^ |  |  |  |  |  |
| Drug and alcohol disorder^b^ | 12,270 (2.8%) | 78 (4.6%) | 0.10 |  | 0.05 |
| Epilepsy | 1,342 (0.3%) | 6 (0.4%) | 0.01 |  | 0.02 |
| Gastro-oesophageal reflux disease | 77,268 (17.4%) | 346 (20.5%) | 0.08 |  | 0.08 |
| Infertility^b^ | 22,628 (5.1%) | 60 (3.6%) | 0.08 |  | 0.02 |
| Gestational hypertension^a,b^ | 11,924 (2.7%) | 53 (3.1%) | 0.03 |  | 0.00 |
| Pre-existing hypertension^a,b^ | 20,300 (4.6%) | 75 (4.5%) | 0.01 |  | 0.03 |
| Gestational diabetes^a,b^ | 18,381 (4.1%) | 69 (4.1%) | 0.00 |  | 0.03 |
| Pre-existing diabetes^a,c,d^ | 120,925 (27.2%) | 338 (20.1%) | 0.17 |  | 0.00 |
| Mental Health | 116,885 (26.3%) | 632 (37.5%) | 0.24 |  | 0.06 |
| Obesity^b^ | 30,987 (7.0%) | 157 (9.3%) | 0.09 |  | 0.06 |
| Respiratory disease^b^ | 13,851 (3.1%) | 96 (5.7%) | 0.13 |  | 0.02 |
| Smoking^a,b^ | 50,268 (11.3%) | 394 (23.4%) | 0.32 |  | 0.00 |
| Thyroid disease^b^ | 52,171 (11.7%) | 114 (6.8%) | 0.17 |  | 0.07 |
| ***Conditions associated with pain*** | | | | | |
| Abdominal and back pain^c^ | 96,214 (21.6%) | 564 (33.5%) | 0.27 |  | 0.04 |
| Chronic viral Hepatitis/HIV^b,c^ | 82,509 (18.6%) | 248 (14.7%) | 0.10 |  | 0.05 |
| Rheumatic disease^b,c^ | 111,175 (25.0%) | 262 (15.5%) | 0.24 |  | 0.05 |
| Surgery | 117,351 (26.4%) | 452 (26.8%) | 0.01 |  | 0.04 |
| Trauma | 4,183 (0.9%) | 42 (2.5%) | 0.12 |  | 0.02 |
| Musculoskeletal pain^c^ | 2,410 (0.5%) | 25 (1.5%) | 0.09 |  | 0.06 |
| ***Medicine use*** | | | | | |
| Metformin | 63,996 (14.4%) | 199 (11.8%) | 0.08 |  | 0.02 |
| Non-opioid analgesics | 52,553 (11.8%) | 335 (19.9%) | 0.33 |  | 0.03 |
| Opioids^d^ | 58,661 (13.2%) | 396 (23.5%) | 0.27 |  | 0.05 |
| Psychotropics | 15,706 (3.5%) | 103 (6.1%) | 0.12 |  | 0.02 |
| Pregnancy category D/X medicines | 62,320 (14.0%) | 218 (12.9%) | 0.03 |  | 0.11 |
| Systemic corticosteroids | 56,802 (12.8%) | 227 (13.5%) | 0.02 |  | 0.06 |
| ***Healthcare utilisation*** | | | | | |
| Number of hospital admissions |  |  |  |  |  |
| 0 | 216,960 (48.8%) | 518 (30.7%) |  |  |  |
| ≥1 | 227,753 (51.2%) | 1,167 (69.3%) | 0.38 |  | 0.03 |
| Number of ED presentations |  |  |  |  |  |
| 0 | 238,376 (53.6%) | 518 (30.7%) |  |  |  |
| ≥1 | 206,337 (46.4%) | 1,167 (69.3%) | 0.48 |  | 0.01 |
| Number of GP visits |  |  |  |  |  |
| 0 | 99,267 (22.3%) | 384 (22.8%) |  |  |  |
| ≥1 | 345,446 (77.7%) | 1,301 (77.2%) | 0.01 |  | 0.07 |

Abbreviations: ED, Emergency department; GP, General practitioners; IPTW, Inverse probability of treatment weighting; Std diff, Standardized differences. Category D/X medicines: based on Australian categorisation system for medicines in pregnancy and includes. suspected teratogens, medicines with other adverse effects.
^a^Based on all or some records recorded at the time of childbirth.
^b^Based on records obtained from the year prior to trial entry until the time of delivery.
^c^Conditions used in the analysis restricting to individuals with chronic conditions associated with pain.
^d^Prior opioid use in the 365 days prior to trial entry until the 90 days prior to conception.

Table 9: Characteristics and absolute standardised mean differences of opioid analgesic initiators and non-initiators before and after weighting in an active‑comparator sensitivity analysis comparing initiation of other opioids (exposed) with initiation of codeine (reference).

| **Characteristic** | **Person trials before weighting**  (n = 26,984) | | |  | **After Weighting** |
| --- | --- | --- | --- | --- | --- |
|  | **Non-initiator person-trials** (n=23,403) | **Opioid initiator person-trials** (n=3,581) | **Absolute std diff** |  | **Absolute std diff** |
| ***Trial characteristics*** | | | | | |
| Year trial started^a^ |  |  |  |  |  |
| 2013-2015 | 9,279 (39.6%) | 1,218 (34.0%) | 0.12 |  | 0.00 |
| 2016-2017 | 7,919 (33.8%) | 1,299 (36.3%) | 0.05 |  | 0.04 |
| 2018-2019 | 6,205 (26.5%) | 1,064 (29.7%) | 0.07 |  | 0.04 |
| ***Maternal Characteristics*** |  |  |  |  |  |
| Maternal age at trial entry (years)^a^ |  |  |  |  |  |
| <25 | 4,703 (20.1%) | 724 (20.2%) | 0.00 |  | 0.00 |
| 25-34 | 14,160 (60.5%) | 2,106 (58.8%) | 0.03 |  | 0.00 |
| ≥35 | 4,540 (19.4%) | 751 (21.0%) | 0.04 |  | 0.00 |
| Country of Birth^a^ |  |  |  |  |  |
| Australian-born | 17,672 (75.5%) | 2,873 (80.2%) | 0.11 |  | 0.05 |
| Partner status^a^ |  |  |  |  |  |
| Has a partner | 18,286 (78.1%) | 2,762 (77.1%) | 0.02 |  | 0.01 |
| Concessional beneficiary | 8,404 (35.9%) | 1,292 (36.1%) | 0.00 |  | 0.01 |
| Quintiles of residential socio-economic disadvantage^a^ |  |  |  |  |  |
| Most disadvantaged | 6,990 (29.9%) | 976 (27.3%) |  |  |  |
| Quintiles 2/3/4/5 (Least disadvantaged) | 16,413 (70.1%) | 2,605 (72.7%) | 0.06 |  | 0.01 |
| Residential remoteness^a^ |  |  |  |  |  |
| Major cities | 17,244 (73.7%) | 2,316 (64.7%) | 0.20 |  | 0.07 |
| Inner regional/Outer regional/Remote/Very remote | 6,159 (26.3%) | 1,265 (35.3%) |  |  |  |
| ***Pregnancy and maternal-related characteristics*** | | | | | |
| Parity^a^ |  |  |  |  |  |
| No previous birth | 8,507 (36.4%) | 1,319 (36.8%) |  |  |  |
| Any previous birth | 14,896 (63.6%) | 2,262 (63.2%) | 0.01 |  | 0.01 |
| Previous caesarean section^a^ | 4,156 (17.8%) | 668 (18.7%) | 0.02 |  | 0.01 |
| Pregnancy interval |  |  |  |  |  |
| No previous birth | 10,362 (44.3%) | 1,600 (44.7%) | 0.01 |  | 0.02 |
| <6 - <33 months | 3,853 (16.5%) | 578 (16.1%) | 0.01 |  | 0.00 |
| ≥33 months | 9,188 (39.3%) | 1,403 (39.2%) | 0.00 |  | 0.02 |
| Multifetal gestation^a^ | 417 (1.8%) | 68 (1.9%) | 0.01 |  | 0.04 |
| Previous adverse maternal outcome | 4,297 (18.4%) | 661 (18.5%) | 0.00 |  | 0.01 |
| Previous adverse neonatal outcome | 2,701 (11.5%) | 412 (11.5%) | 0.00 |  | 0.02 |
| ***Maternal morbidities*** | | | | | |
| Anaemia and coagulation^b^ | 2,123 (9.1%) | 525 (14.7%) | 0.17 |  | 0.01 |
| Cardiovascular disease^b^ | 1,066 (4.6%) | 201 (5.6%) | 0.05 |  | 0.00 |
| Chronic liver failure^b,c^ | 215 (0.9%) | 48 (1.3%) | 0.04 |  | 0.01 |
| Chronic renal disease^b,c^ | 230 (1.0%) | 71 (2.0%) | 0.08 |  | 0.04 |
| Drug and alcohol disorder^b^ | 597 (2.6%) | 115 (3.2%) | 0.04 |  | 0.02 |
| Epilepsy | 52 (0.2%) | 13 (0.4%) | 0.03 |  | 0.02 |
| Gastro-oesophageal reflux disease | 2,743 (11.7%) | 508 (14.2%) | 0.07 |  | 0.01 |
| Infertility^b^ | 640 (2.7%) | 140 (3.9%) | 0.07 |  | 0.03 |
| Gestational hypertension^a,b^ | 661 (2.8%) | 92 (2.6%) | 0.02 |  | 0.01 |
| Pre-existing hypertension^a,b^ | 340 (1.5%) | 78 (2.2%) | 0.05 |  | 0.00 |
| Gestational diabetes^a,b^ | 1,305 (5.6%) | 166 (4.6%) | 0.04 |  | 0.03 |
| Pre-existing diabetes^a,c,d^ | 281 (1.2%) | 57 (1.6%) | 0.03 |  | 0.01 |
| Mental Health | 6,386 (27.3%) | 1,192 (33.3%) | 0.13 |  | 0.02 |
| Obesity^b^ | 1,188 (5.1%) | 216 (6.0%) | 0.04 |  | 0.01 |
| Respiratory disease^b^ | 463 (2.0%) | 121 (3.4%) | 0.09 |  | 0.00 |
| Smoking^a,b^ | 3,568 (15.2%) | 632 (17.6%) | 0.06 |  | 0.01 |
| Thyroid disease^b^ | 1,387 (5.9%) | 209 (5.8%) | 0.00 |  | 0.02 |
| ***Conditions associated with pain*** | | | | | |
| Abdominal and back pain^c^ | 430 (1.8%) | 133 (3.7%) | 0.11 |  | 0.02 |
| Chronic viral Hepatitis/HIV^b,c^ | 2,562 (10.9%) | 446 (12.5%) | 0.05 |  | 0.02 |
| Rheumatic disease^b,c^ | 197 (0.8%) | 65 (1.8%) | 0.09 |  | 0.01 |
| Surgery | 2,767 (11.8%) | 796 (22.2%) | 0.28 |  | 0.04 |
| Trauma | 245 (1.0%) | 131 (3.7%) | 0.17 |  | 0.01 |
| Musculoskeletal pain^c^ | 16 (0.1%) | 9 (0.3%) | 0.05 |  | 0.00 |
| ***Medicine use*** | | | | | |
| Metformin | 1,033 (4.4%) | 144 (4.0%) | 0.02 |  | 0.01 |
| Non-opioid analgesics | 2,562 (10.9%) | 446 (12.5%) | 0.05 |  | 0.02 |
| Opioids^d^ | 2,863 (12.2%) | 546 (15.2%) | 0.09 |  | 0.01 |
| Psychotropics | 816 (3.5%) | 163 (4.6%) | 0.05 |  | 0.01 |
| Pregnancy category D/X medicines | 1,796 (7.7%) | 341 (9.5%) | 0.07 |  | 0.04 |
| Systemic corticosteroids | 1,619 (6.9%) | 308 (8.6%) | 0.06 |  | 0.01 |
| ***Healthcare utilisation*** | | | | | |
| Number of hospital admissions |  |  |  |  |  |
| 0 | 15,761 (67.3%) | 1,800 (50.3%) |  |  |  |
| ≥1 | 7,642 (32.7%) | 1,781 (49.7%) | 0.35 |  | 0.02 |
| Number of ED presentations |  |  |  |  |  |
| 0 | 13,187 (56.3%) | 1,539 (43.0%) |  |  |  |
| ≥1 | 10,216 (43.7%) | 2,042 (57.0%) | 0.27 |  | 0.02 |
| Number of GP visits |  |  |  |  |  |
| 0 | 6,992 (29.9%) | 1,100 (30.7%) |  |  |  |
| ≥1 | 16,411 (70.1%) | 2,481 (69.3%) | 0.02 |  | 0.02 |

Abbreviations: ED, Emergency Department; GP, General practitioners; IPTW, Inverse probability of treatment weighting; Std diff, Standardized differences. Category D/X medicines: based on Australian categorisation system for medicines in pregnancy and includes. suspected teratogens, medicines with other adverse effects.
^a^Based on all or some records recorded at the time of childbirth.
^b^Based on records obtained from the year prior to trial entry until the time of delivery.
^c^Conditions used in the analysis restricting to individuals with chronic conditions associated with pain.
^d^Prior opioid use in the 365 days prior to trial entry until the 90 days prior to conception.

Table 10: Characteristics and absolute standardised mean differences of opioid analgesic initiators and non-initiators before and after weighting, in a sensitivity analysis restricted to pregnancies conceived on or after February 2018.

| **Characteristic** | **Person trials before weighting**  (n = 3,267,177) | | |  | **After Weighting** |
| --- | --- | --- | --- | --- | --- |
|  | **Non-initiator person-trials** (n=3,262,422) | **Opioid initiator person-trials** (n=4,755) | **Absolute std diff** |  | **Absolute std diff** |
| ***Trial characteristics*** | | | | | |
| Year trial started^a^ |  |  |  |  |  |
| 2018 | 1,596,873 (48.9%) | 2,182 (45.9%) |  |  |  |
| 2019 | 1,665,549 (51.1%) | 2,573 (54.1%) | 0.06 |  | 0.05 |
| ***Maternal Characteristics*** |  |  |  |  |  |
| Maternal age at trial entry (years)^a^ |  |  |  |  |  |
| <25 | 443,353 (13.6%) | 833 (17.5%) | 0.11 |  | 0.02 |
| 25-34 | 2,059,149 (63.1%) | 2,940 (61.8%) | 0.03 |  | 0.02 |
| ≥35 | 759,920 (23.3%) | 982 (20.7%) | 0.06 |  | 0.00 |
| Country of Birth^a^ |  |  |  |  |  |
| Australian-born | 1,975,845 (60.6%) | 3,678 (77.4%) | 0.37 |  | 0.02 |
| Partner status^a^ |  |  |  |  |  |
| Has a partner | 2,830,426 (86.8%) | 3,778 (79.5%) | 0.20 |  | 0.02 |
| Concessional beneficiary | 622,438 (19.1%) | 1,477 (31.1%) | 0.28 |  | 0.01 |
| Quintiles of residential socio-economic disadvantage^a^ |  |  |  |  |  |
| Most disadvantaged | 771,090 (23.6%) | 1,366 (28.7%) | 0.12 |  | 0.02 |
| 2 | 633,964 (19.4%) | 1,086 (22.8%) | 0.08 |  | 0.03 |
| 3 | 725,855 (22.2%) | 1,015 (21.3%) | 0.02 |  | 0.02 |
| 4 | 459,782 (14.1%) | 566 (11.9%) | 0.07 |  | 0.01 |
| Least disadvantaged | 671,731 (20.6%) | 722 (15.2%) | 0.14 |  | 0.01 |
| Residential remoteness^a^ |  |  |  |  |  |
| Major cities | 2,591,780 (79.4%) | 3,376 (71.0%) | 0.20 |  | 0.02 |
| Inner regional | 505,249 (15.5%) | 1,013 (21.3%) | 0.15 |  | 0.02 |
| Outer regional | 150,317 (4.6%) | 339 (7.1%) | 0.11 |  | 0.00 |
| Remote/Very remote | 15,076 (0.5%) | 27 (0.6%) | 0.01 |  | 0.01 |
| ***Pregnancy and maternal-related characteristics*** | | | | | |
| Parity^a^ |  |  |  |  |  |
| No previous birth | 1,414,479 (43.4%) | 1,713 (36.0%) | 0.15 |  | 0.02 |
| 1 | 1,144,227 (35.1%) | 1,647 (34.6%) | 0.01 |  | 0.02 |
| 2 | 459,800 (14.1%) | 806 (17.0%) | 0.08 |  | 0.00 |
| ≥3 | 243,916 (7.5%) | 589 (12.4%) | 0.16 |  | 0.01 |
| Previous caesarean section^a^ | 516,799 (15.8%) | 919 (19.3%) | 0.09 |  | 0.00 |
| Pregnancy interval |  |  |  |  |  |
| No previous birth | 1,644,763 (50.4%) | 1,969 (41.4%) | 0.18 |  | 0.02 |
| <6 months | 61,531 (1.9%) | 105 (2.2%) | 0.02 |  | 0.03 |
| 6 - <18 months | 477,768 (14.6%) | 762 (16.0%) | 0.04 |  | 0.02 |
| 18 - <33 months | 523,625 (16.1%) | 788 (16.6%) | 0.01 |  | 0.01 |
| ≥33 months | 554,735 (17.0%) | 1,131 (23.8%) | 0.17 |  | 0.03 |
| Multifetal gestation^a^ | 41,530 (1.3%) | 77 (1.6%) | 0.04 |  | 0.03 |
| Previous adverse maternal outcome | 440,522 (13.5%) | 899 (18.9%) | 0.15 |  | 0.00 |
| Previous adverse neonatal outcome | 297,259 (9.1%) | 541 (11.4%) | 0.07 |  | 0.00 |
| ***Maternal morbidities*** | | | | | |
| Anaemia and coagulation^b^ | 158,043 (4.8%) | 532 (11.2%) | 0.24 |  | 0.05 |
| Cardiovascular disease^b^ | 83,569 (2.6%) | 250 (5.3%) | 0.14 |  | 0.01 |
| Chronic liver failure^b,c^ | 14,059 (0.4%) | 42 (0.9%) | 0.06 |  | 0.02 |
| Chronic renal disease^b,c^ | 8,347 (0.3%) | 54 (1.1%) | 0.11 |  | 0.08 |
| Drug and alcohol disorder^b^ | 26,251 (0.8%) | 116 (2.4%) | 0.13 |  | 0.01 |
| Epilepsy | 3,867 (0.1%) | 11 (0.2%) | 0.03 |  | 0.01 |
| Gastro-oesophageal reflux disease | 195,821 (6.0%) | 586 (12.3%) | 0.22 |  | 0.04 |
| Infertility^b^ | 89,639 (2.7%) | 166 (3.5%) | 0.04 |  | 0.04 |
| Gestational hypertension^a,b^ | 48,201 (1.5%) | 113 (2.4%) | 0.06 |  | 0.01 |
| Pre-existing hypertension^a,b^ | 33,490 (1.0%) | 94 (2.0%) | 0.08 |  | 0.01 |
| Gestational diabetes^a,b^ | 175,844 (5.4%) | 245 (5.2%) | 0.06 |  | 0.01 |
| Pre-existing diabetes^a,c,d^ | 23,137 (0.7%) | 64 (1.3%) | 0.06 |  | 0.01 |
| Mental Health | 527,513 (16.2%) | 1,531 (32.2%) | 0.38 |  | 0.04 |
| Obesity^b^ | 106,776 (3.3%) | 473 (9.9%) | 0.27 |  | 0.06 |
| Respiratory disease^b^ | 35,302 (1.1%) | 157 (3.3%) | 0.15 |  | 0.02 |
| Smoking^a,b^ | 231,861 (7.1%) | 728 (15.3%) | 0.26 |  | 0.00 |
| Thyroid disease^b^ | 218,272 (6.7%) | 292 (6.1%) | 0.02 |  | 0.01 |
| ***Conditions associated with pain*** | | | | | |
| Abdominal and back pain^c^ | 17,386 (0.5%) | 100 (2.1%) | 0.08 |  | 0.02 |
| Chronic viral Hepatitis/HIV^b,c^ | 14,159 (0.4%) | 42 (0.9%) | 0.06 |  | 0.02 |
| Rheumatic disease^b,c^ | 21,591 (0.7%) | 55 (1.2%) | 0.05 |  | 0.00 |
| Surgery | 236,564 (7.3%) | 675 (14.2%) | 0.23 |  | 0.01 |
| Trauma | 9,733 (0.3%) | 51 (1.1%) | 0.09 |  | 0.01 |
| ***Medicine use*** | | | | | |
| Metformin | 108,325 (3.3%) | 252 (5.3%) | 0.03 |  | 0.01 |
| Non-opioid analgesics | 152,076 (4.7%) | 624 (13.1%) | 0.30 |  | 0.03 |
| Opioids^d^ | 148,701 (4.6%) | 603 (12.7%) | 0.29 |  | 0.02 |
| Psychotropics | 43,541 (1.3%) | 184 (3.9%) | 0.16 |  | 0.01 |
| Pregnancy category D/X medicines | 250,430 (7.7%) | 479 (10.1%) | 0.08 |  | 0.02 |
| Systemic corticosteroids | 117,704 (3.6%) | 388 (8.2%) | 0.19 |  | 0.04 |
| ***Healthcare utilisation*** | | | | | |
| Number of hospital admissions |  |  |  |  |  |
| 0 | 2,553,369 (78.3%) | 3,099 (65.2%) | 0.29 |  | 0.05 |
| 1 | 475,636 (14.6%) | 1,024 (21.5%) | 0.18 |  | 0.05 |
| ≥2 | 233,417 (7.2%) | 632 (13.3%) | 0.01 |  | 0.00 |
| Number of ED presentations |  |  |  |  |  |
| 0 | 2,381,744 (73.0%) | 2,502 (52.6%) | 0.43 |  | 0.04 |
| 1 | 549,579 (16.8%) | 1,110 (23.3%) | 0.16 |  | 0.04 |
| ≥2 | 331,099 (10.1%) | 1,143 (24.0%) | 0.38 |  | 0.01 |
| Number of GP visits |  |  |  |  |  |
| 0 | 973,487 (29.8%) | 1,284 (27.0%) | 0.06 |  | 0.07 |
| 1 | 792,018 (24.3%) | 1,192 (25.1%) | 0.03 |  | 0.02 |
| ≥2 | 1,496,917 (45.9%) | 2,279 (47.9%) | 0.04 |  | 0.04 |

Abbreviations: ED, Emergency Department; GP, General practitioners; IPTW, Inverse probability of treatment weighting; St diff, Standardized differences. Category D/X medicines: based on Australian categorisation system for medicines in pregnancy and includes. suspected teratogens, medicines with other adverse effects.
^a^Based on all or some records recorded at the time of childbirth.
^b^Based on records obtained from the year prior to trial entry until the time of delivery.
^c^Conditions used in the analysis restricting to individuals with chronic conditions associated with pain.
^d^Prior opioid use in the 365 days prior to trial entry until the 90 days prior to conception.

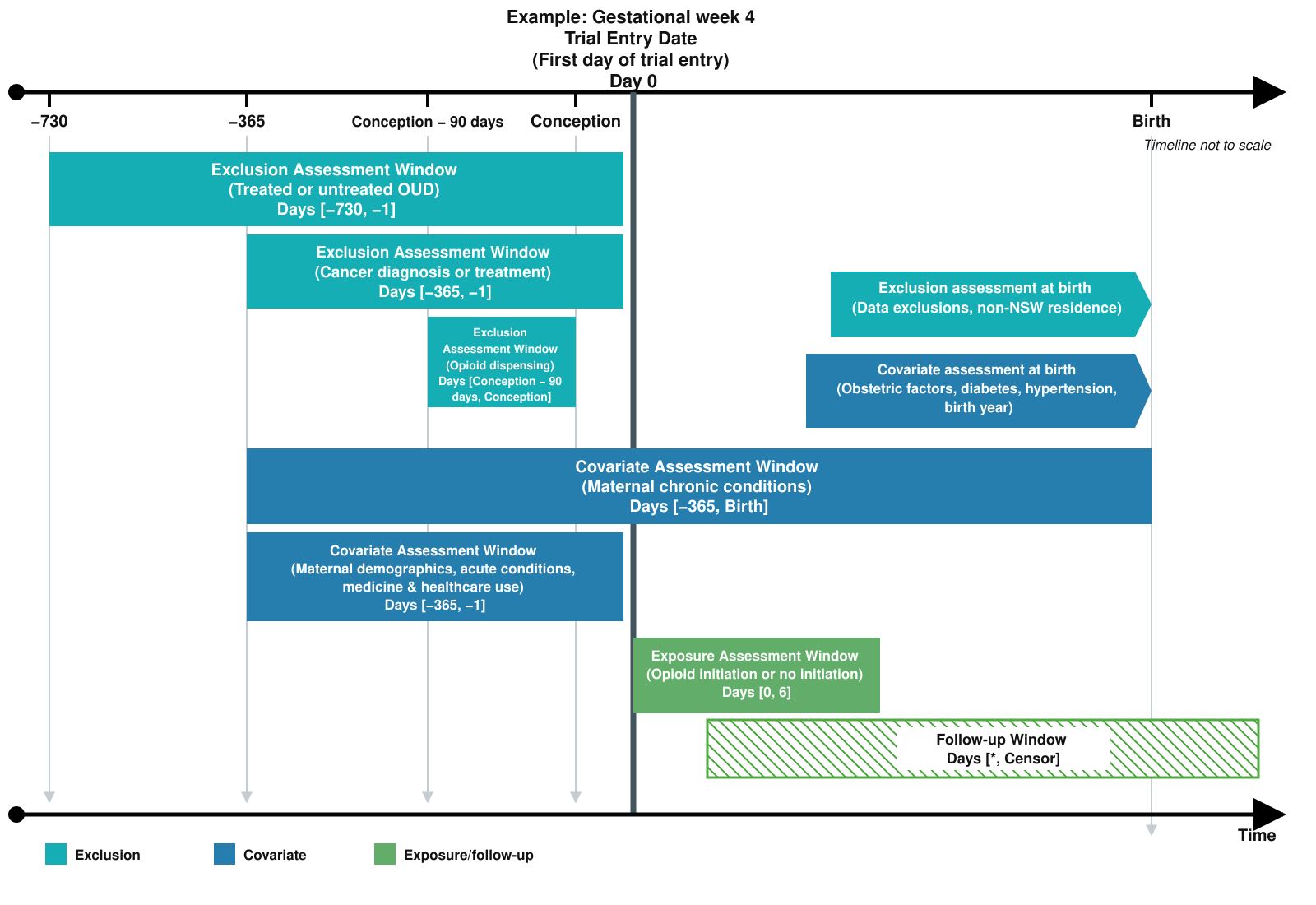

Figure 1: Study design diagram. Example trial occurring in gestational week 4.

* Within each gestational week, follow-up begun at the date of dispensing for those assigned to opioid treatment and midweek for those assigned to no opioid treatment and ends at the administrative end of follow-up.
a. For each gestational week at trial entry, stabilised inverse probability of treatment weighting based on propensity scores was used to balance covariates between the treatment and control arms.
b. Censored at outcome of interest or administrative end of study (discharge after birth for maternal outcomes and 28 days after birth for infant outcomes, or 31^st^ December 2019).

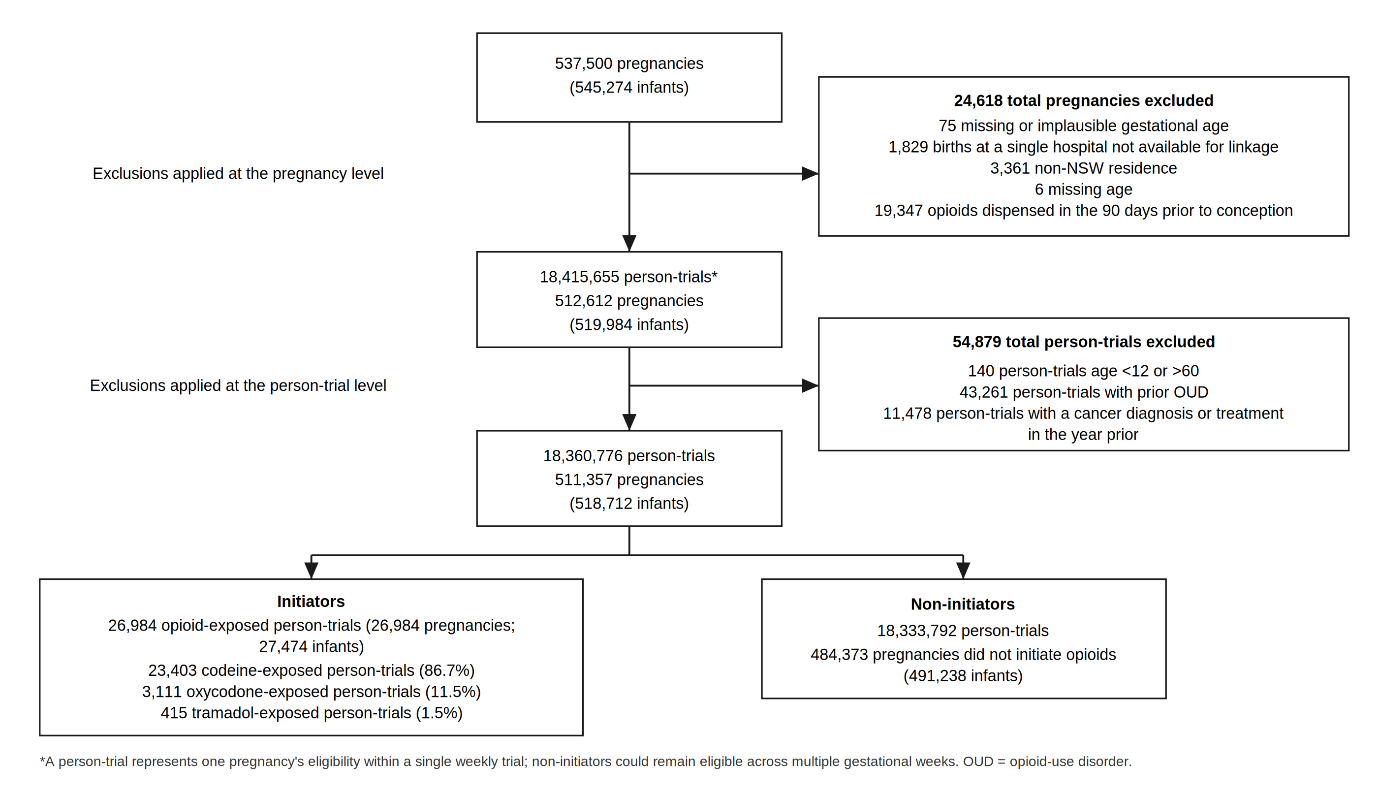

Figure 2: Flowchart of pregnancies, infants, and person-trials included in the analysis. Exclusions were applied at the pregnancy level and the person-trial level. Because non-initiators could remain eligible across multiple weekly trials, the number of person-trials exceeds the number of pregnancies. The final analytic sample comprised 18,360,776 person-trials (511,357 pregnancies; 518,712 infants): 26,984 initiator and 18,333,792 non-initiator person-trials. A person-trial represents one pregnancy's eligibility within a single weekly trial. OUD = opioid-use disorder; NSW = New South Wales.

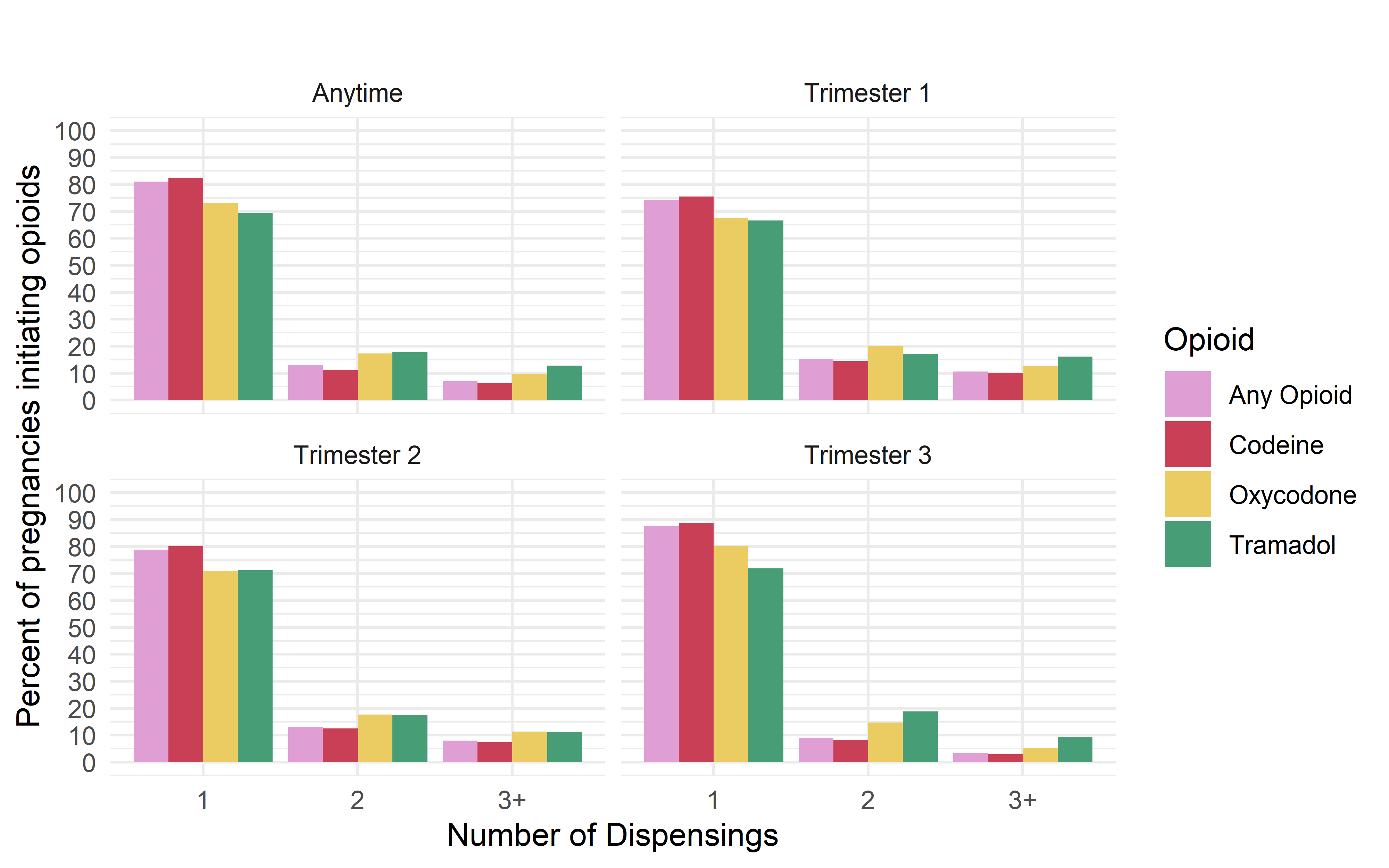

Figure 3: The percentage of opioid initiating pregnancies that received 1, 2, or 3 or more opioid dispensing during the gestational period, categorised by trimester of initiation and type of opioid initially dispensed.

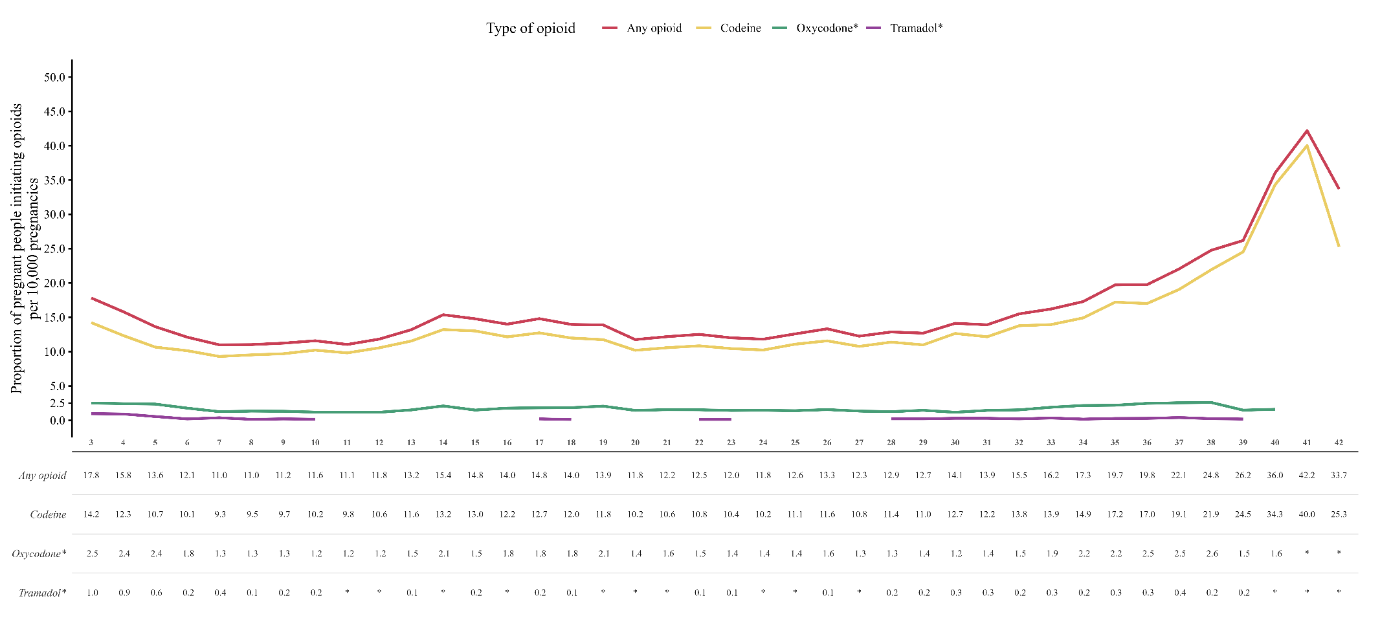

Figure 4: Analysis examining the proportion of pregnant people initiating opioids per 10,000 pregnancies according to the gestational week of initiation and type of opioid initiated.

* For privacy reasons, pregnancies initiating oxycodone or tramadol during gestational weeks with small cell counts are not shown in this figure.

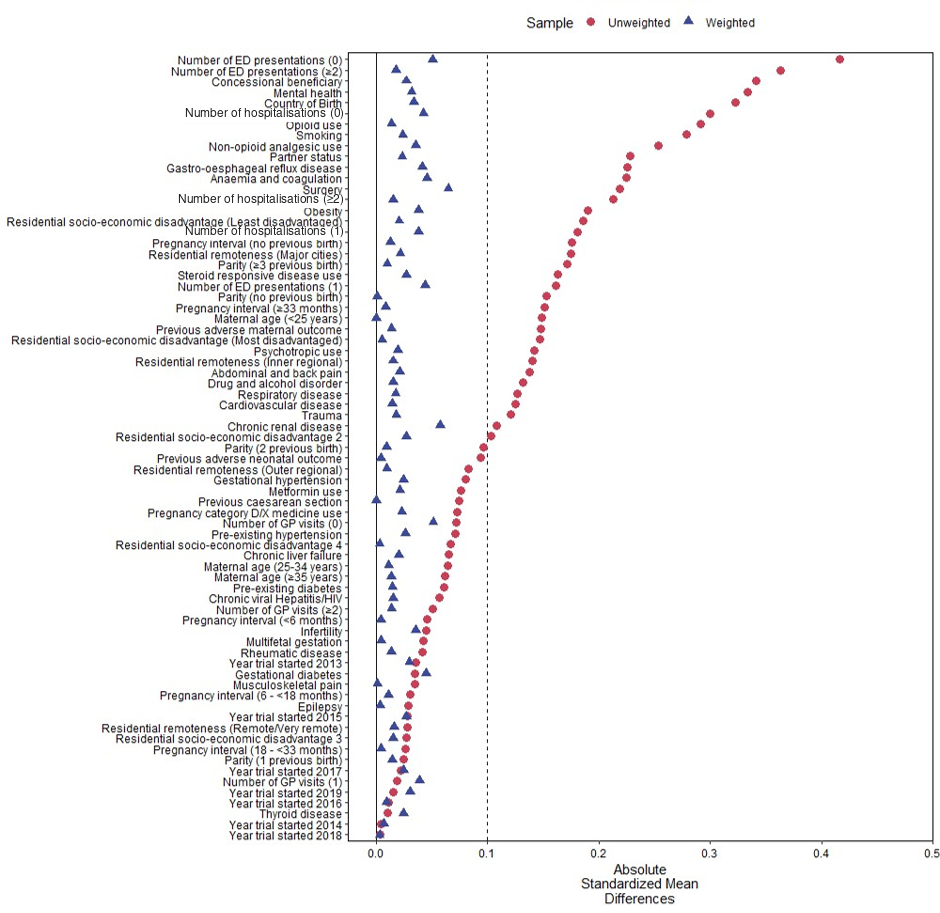

Figure 5: Absolute standardised mean differences in baseline characteristics between opioid initiators and non-initiators, before and after applying stabilised inverse probability of treatment weighting. Values closer to zero indicate better covariate balance. The dotted line represents the conventional thresholds of ±0.1 for acceptable balance.

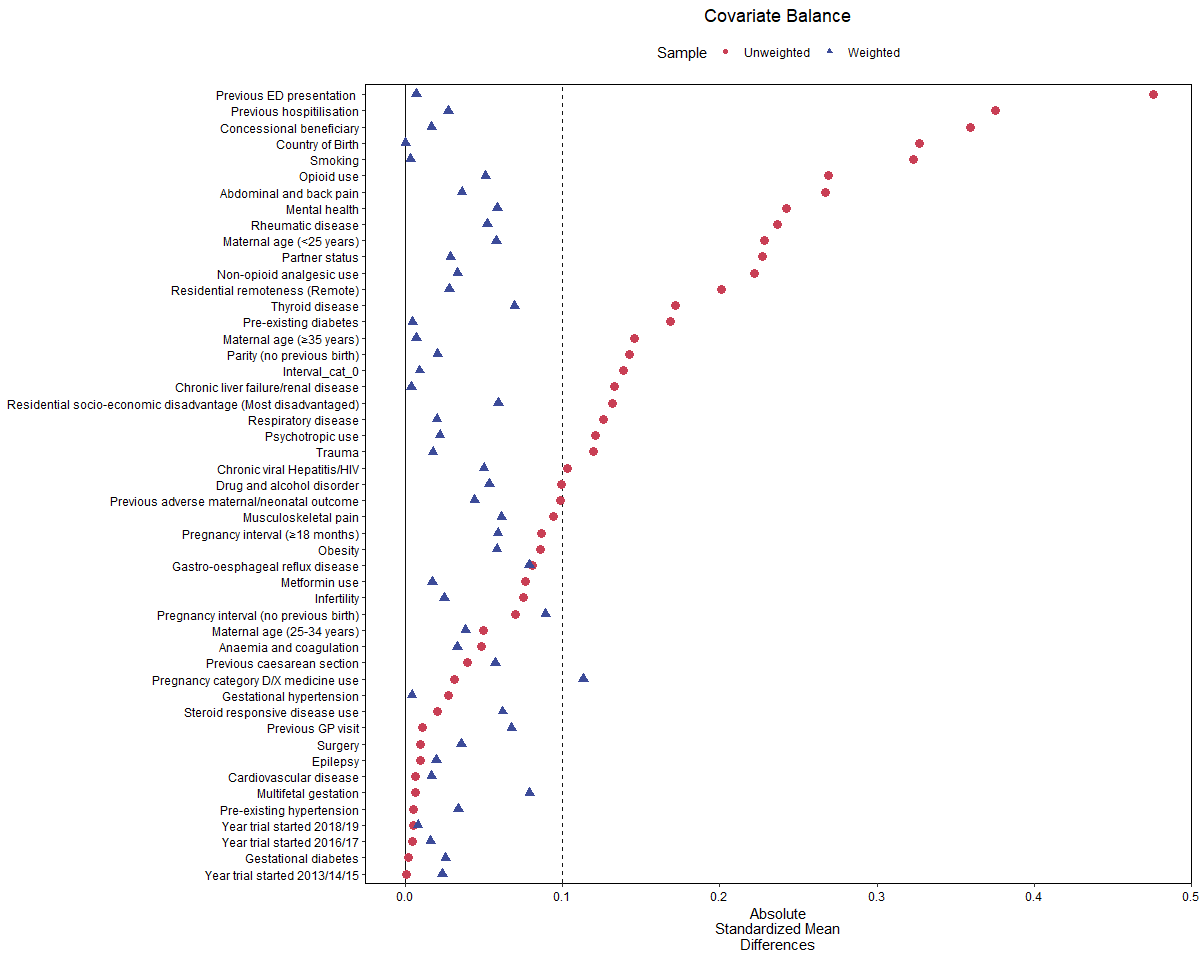

Figure 6: Absolute standardised mean differences in baseline characteristics between opioid initiators and non-initiators, before and after applying stabilised inverse probability of treatment weighting, in a sensitivity analysis restricting the cohort to women with a history of chronic conditions associated with pain. Values closer to zero indicate better covariate balance. The dotted line represents the conventional thresholds of ±0.1 for acceptable balance.

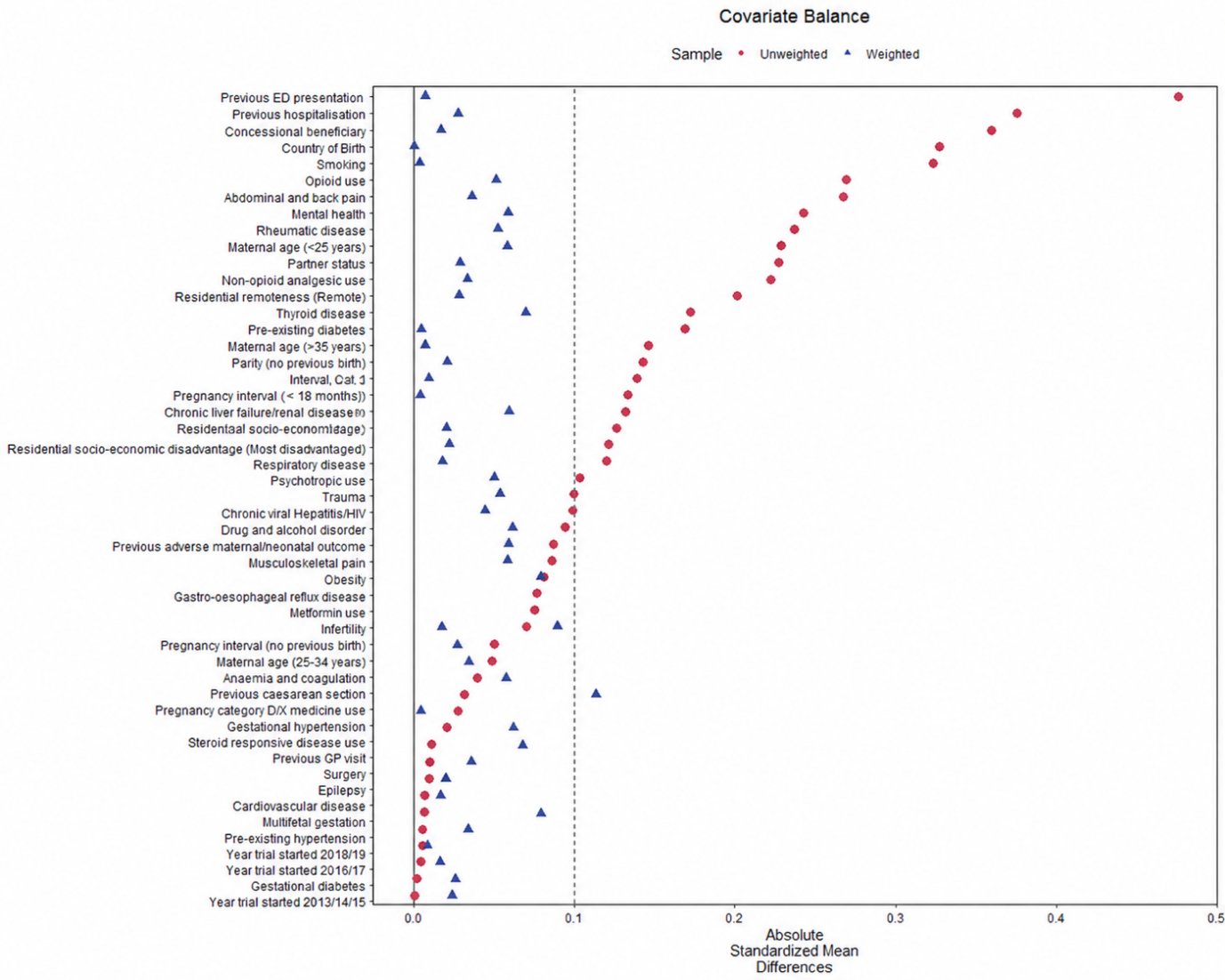

Figure 7: Absolute standardised mean differences in baseline characteristics between opioid initiators and non‑initiators before and after applying stabilized inverse probability of treatment weighting, in an active‑comparator sensitivity analysis comparing initiation of other opioids (exposed) with initiation of codeine (reference). Values closer to zero indicate better covariate balance. The dotted line represents the conventional thresholds of ±0.1 for acceptable balance.

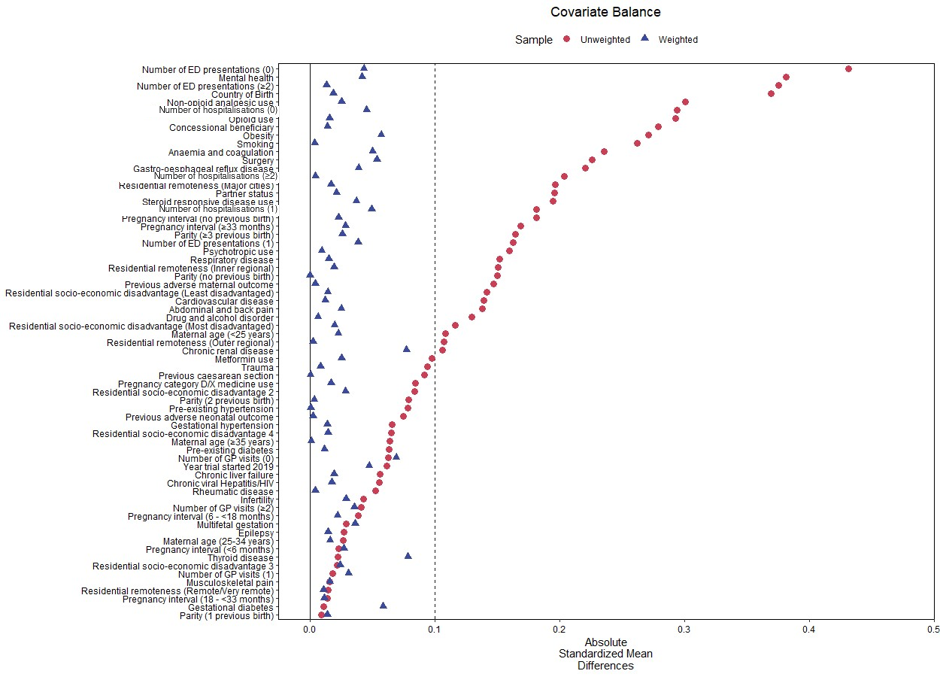

Figure 8: Absolute standardised mean differences in baseline characteristics between opioid initiators and non-initiators, before and after applying stabilised inverse probability of treatment weighting, in a sensitivity analysis restricted to pregnancies conceived on or after February 2018. Values closer to zero indicate better covariate balance. The dotted line represents the conventional thresholds of ±0.1 for acceptable balance.

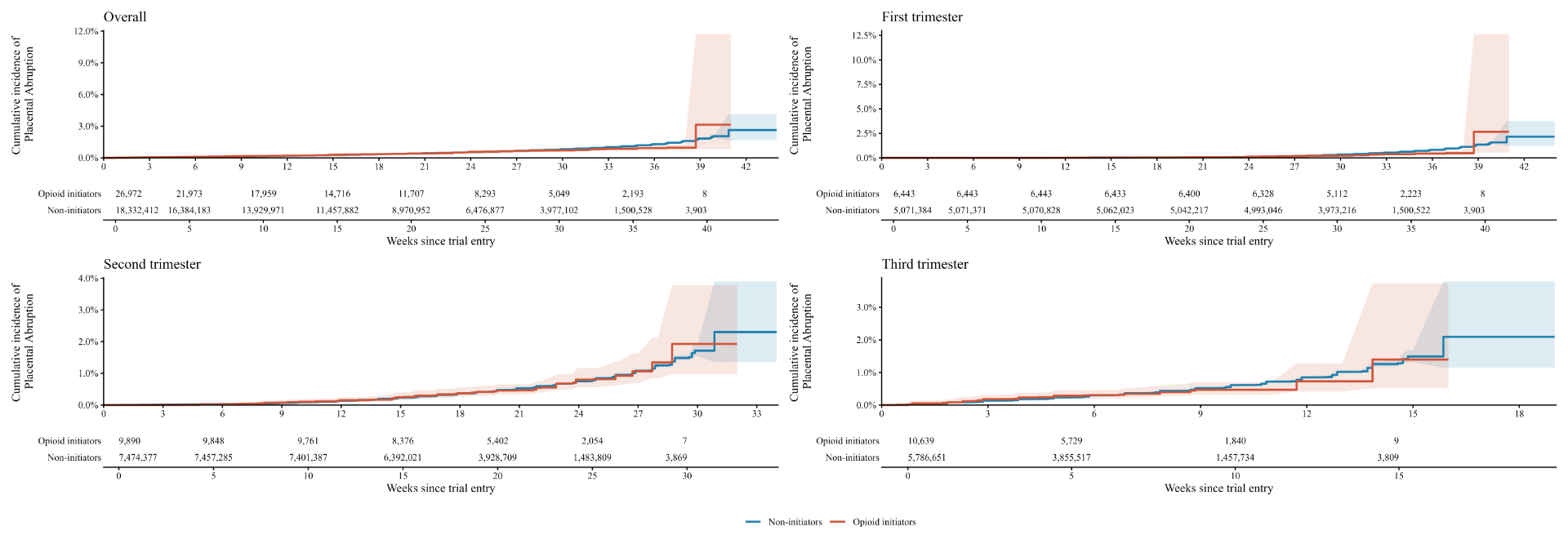

Figure 9: Adjusted risk of placental abruption by number of weeks since trial entry among opioid analgesic initiators compared with non-initiators, shown for overall: all gestational weeks combined and by trimester of treatment initiation: first, second and third trimester.

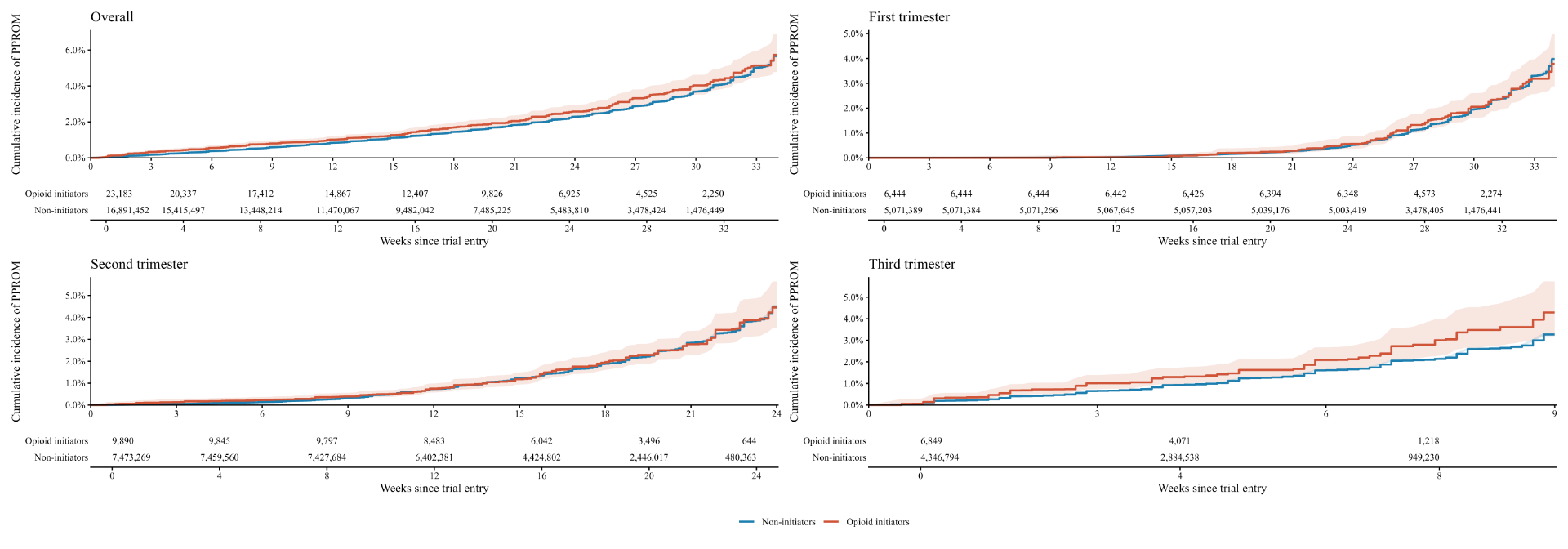

Figure 10: Adjusted risk of preterm premature rupture of membranes (PPROM) by number of weeks since trial entry among opioid analgesic initiators compared with non-initiators, shown for overall: all gestational weeks combined and by trimester of treatment initiation: first, second and third trimester.

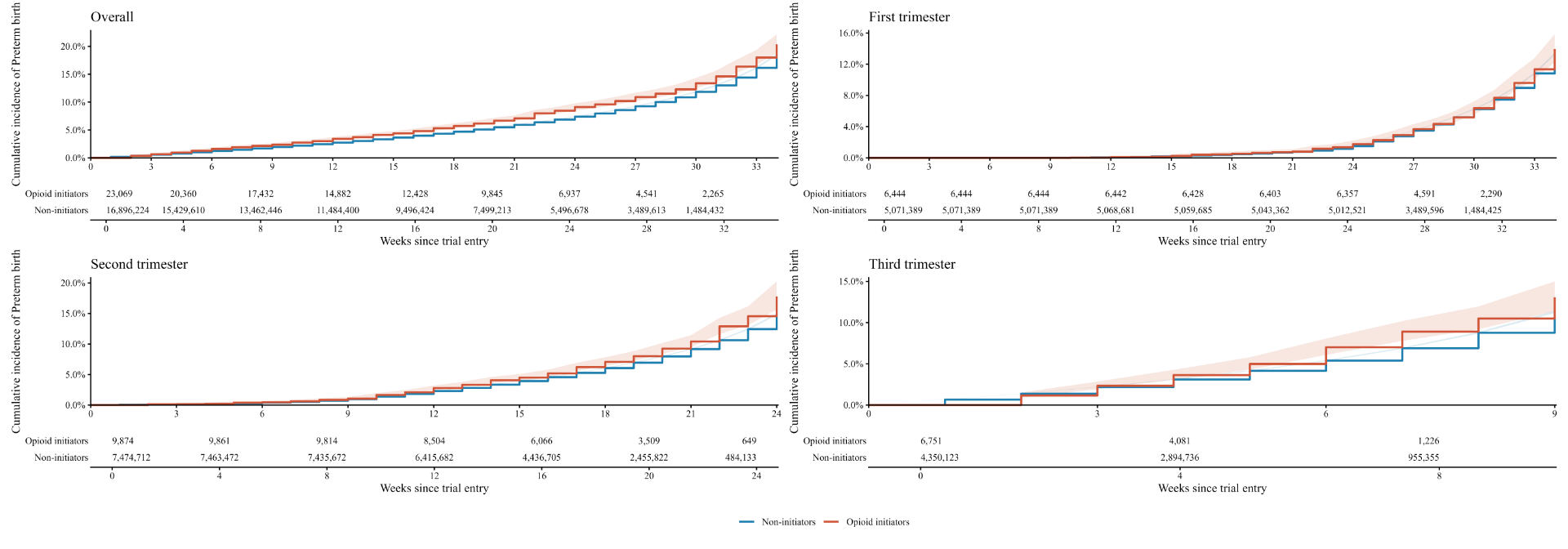

Figure 11: Adjusted risk of preterm birth by number of weeks since trial entry among opioid analgesic initiators compared with non-initiators, shown for overall: all gestational weeks combined and by trimester of treatment initiation: first, second and third trimester.

(a)

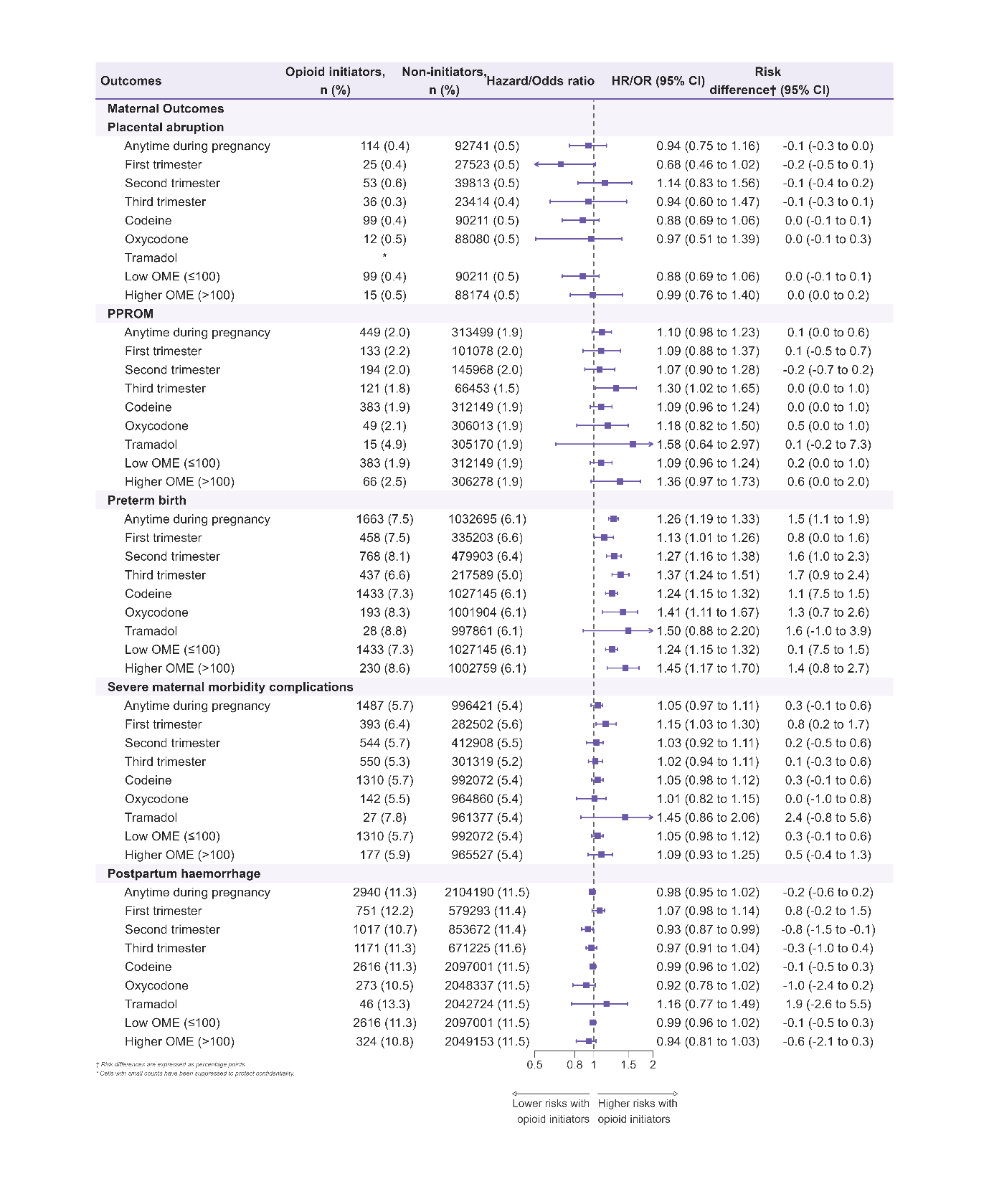

(b)

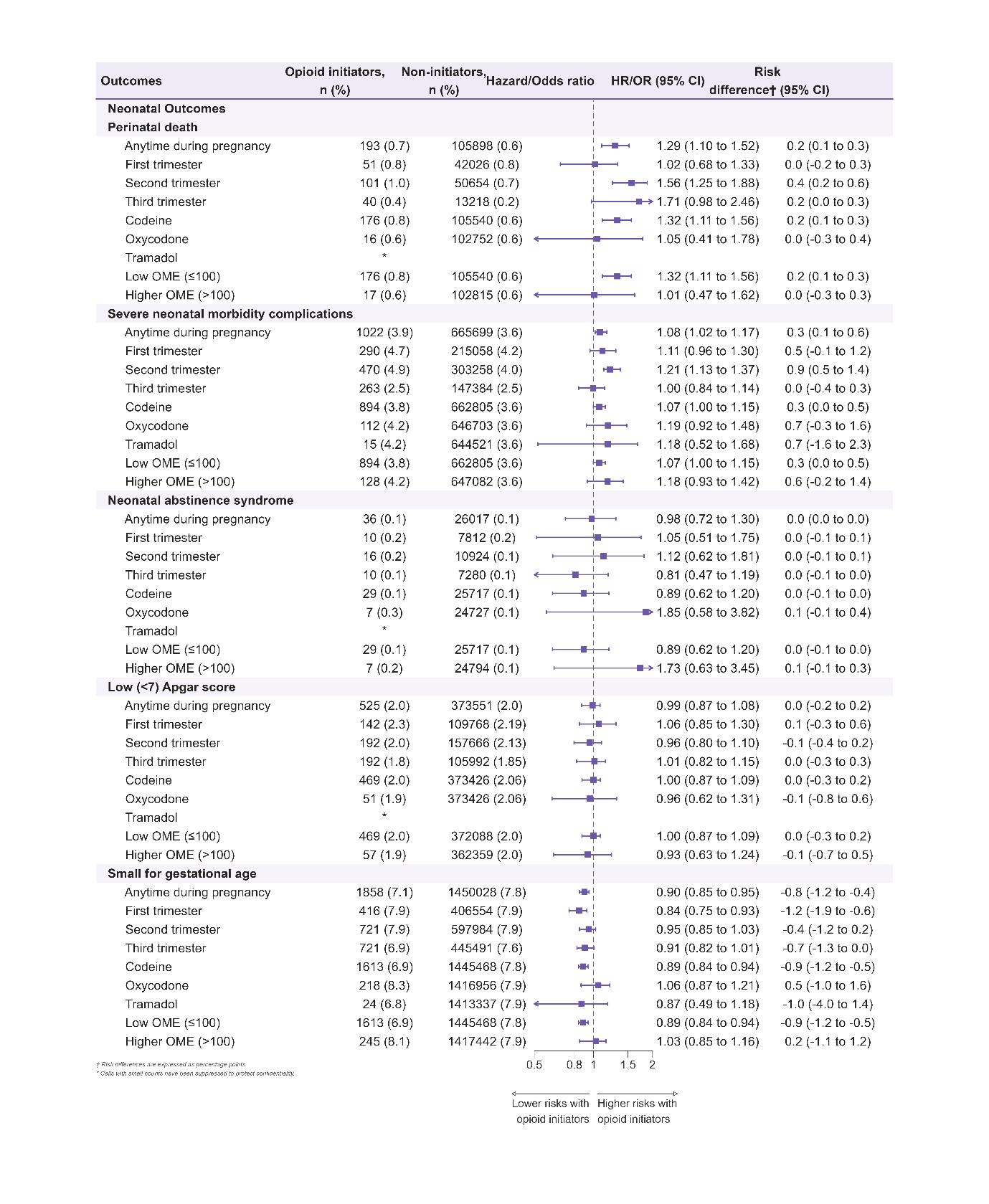

Figure 12: Subgroup analyses of adjusted odds ratios and risk differences (expressed as percentages) for adverse (a) maternal and (b) neonatal outcomes among women who initiated opioids during pregnancy compared with non‑initiators. These analyses correspond to Figure 3 and provide results in greater detail. Analyses are stratified by trimester of initiation, opioid type, and opioid dose (OME). The number and proportion of each outcome within each treatment group are shown as n (%).

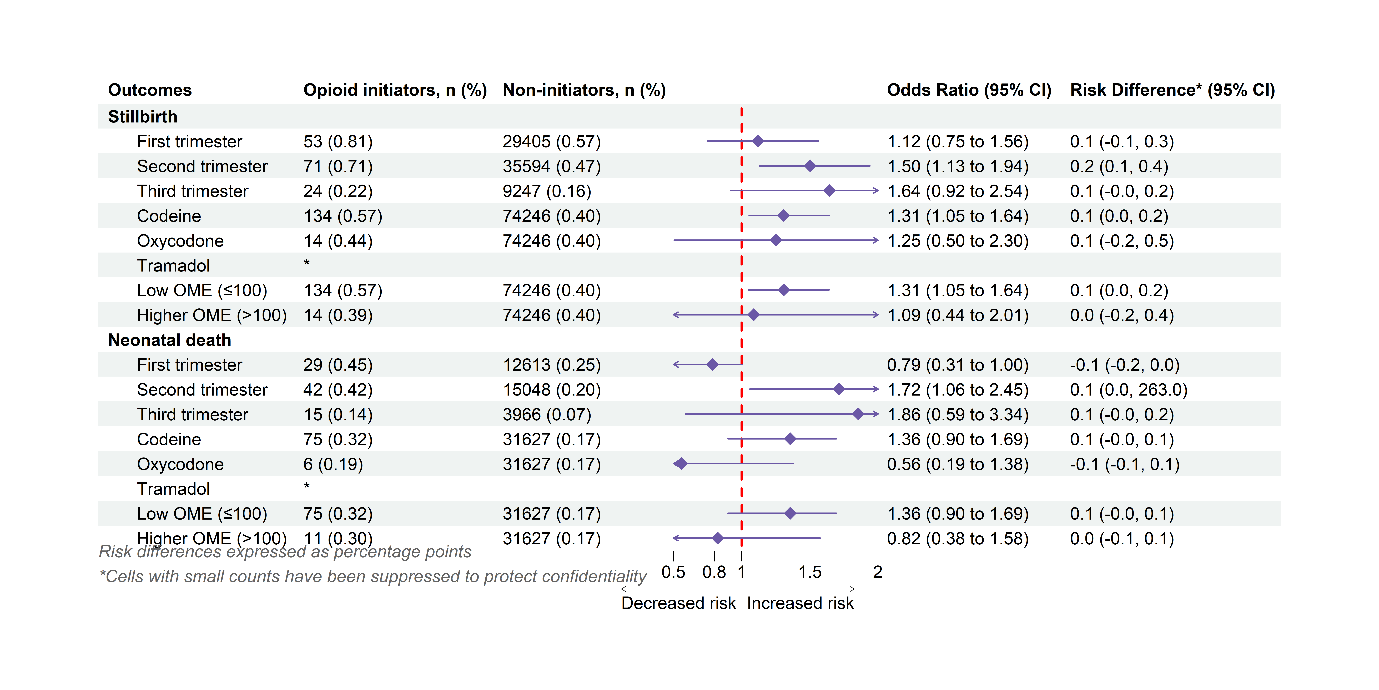

Figure 13: Subgroup analyses of adjusted odds ratios and risk differences (expressed as percentages) for stillbirth and neonatal death, components of the perinatal death outcome, comparing women who initiated opioids during pregnancy with non-initiators. Estimates are stratified by trimester of initiation, opioid type, and opioid dose (OME). The number and proportion of events in each group are presented as n (%).

(a)

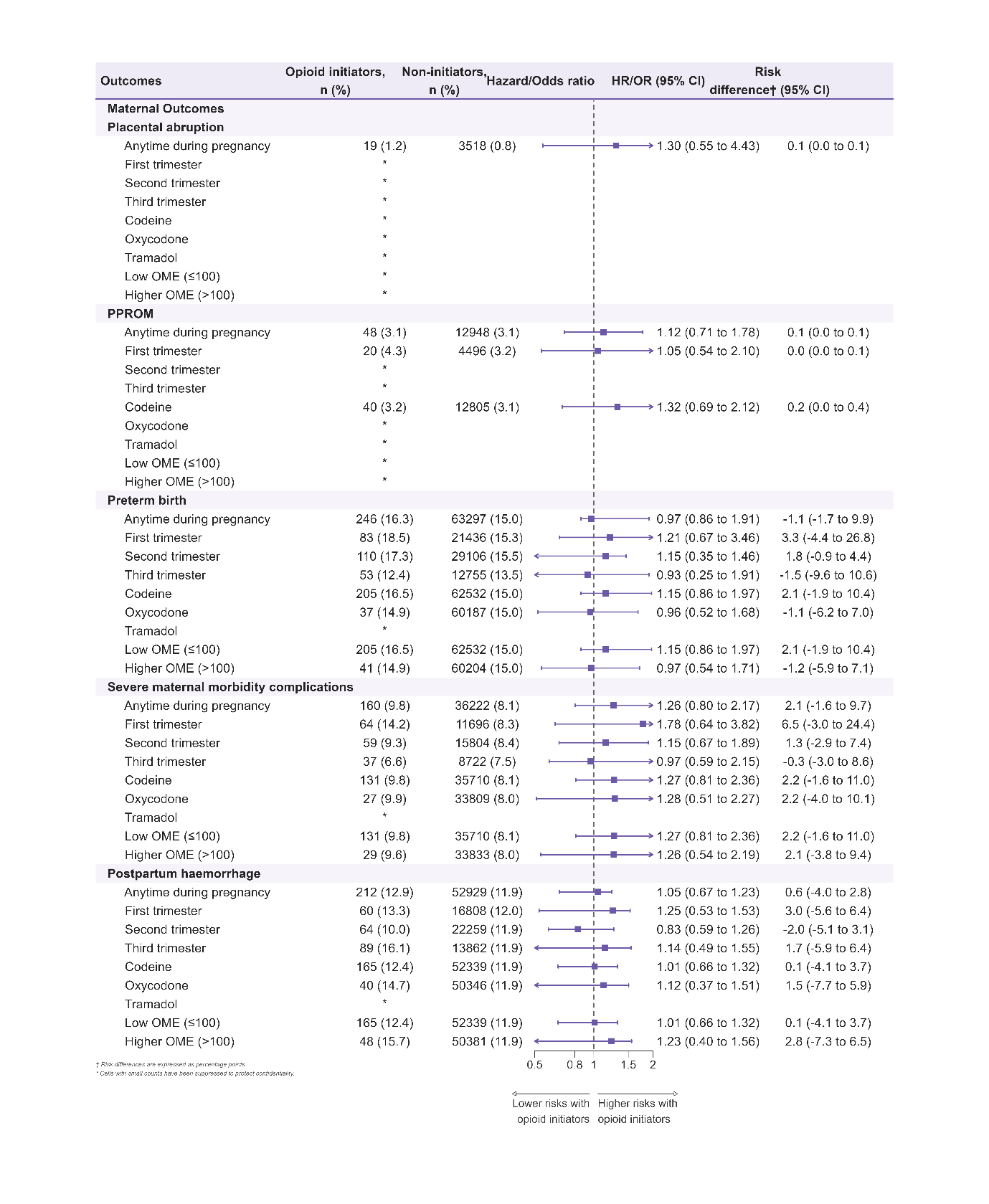

(b)

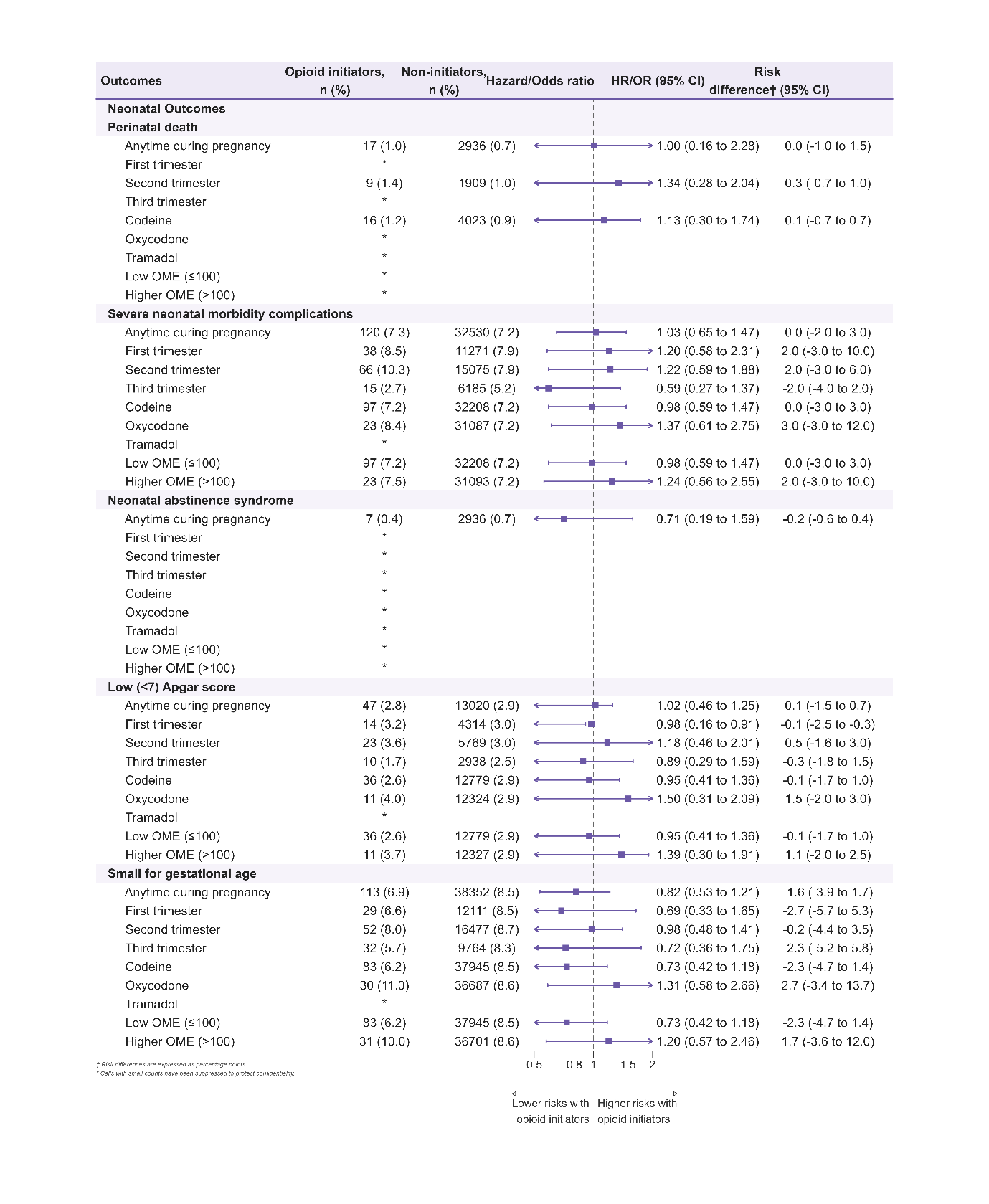
**Figure 14:** Sensitivity analyses of adjusted hazard or odds ratios and risk differences (expressed as percentage points) of adverse (a) maternal and (b) neonatal outcomes restricted to women with chronic pain-related conditions.

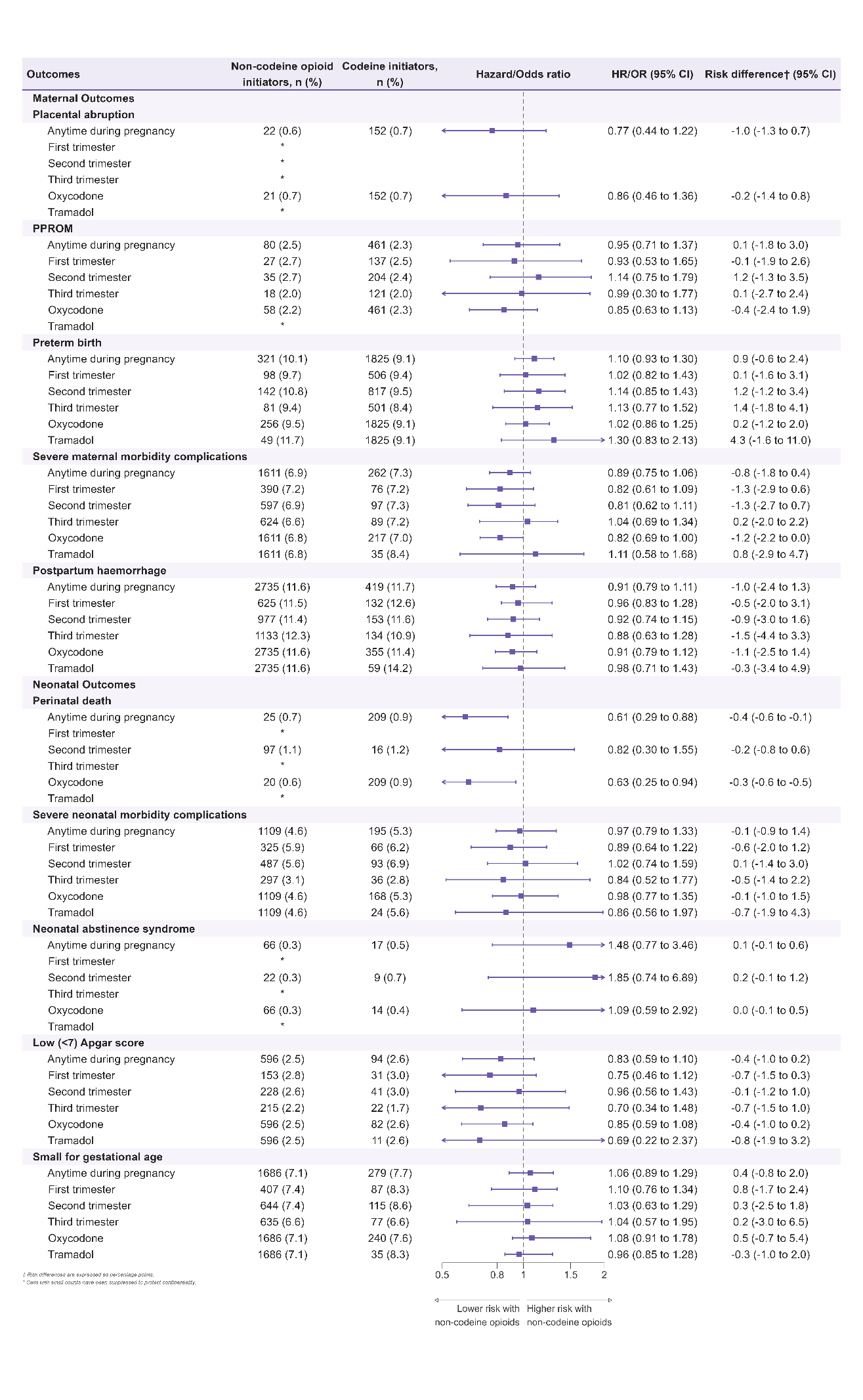

Figure 15: Sensitivity analyses of adjusted hazard or odds ratios and risk differences (expressed as percentages) of adverse maternal and neonatal outcomes following non-codeine opioid initiation versus codeine initiation (reference) during pregnancy.

* Risk estimates represent hazard ratios; remaining risk estimates represent odds ratios.

(a)

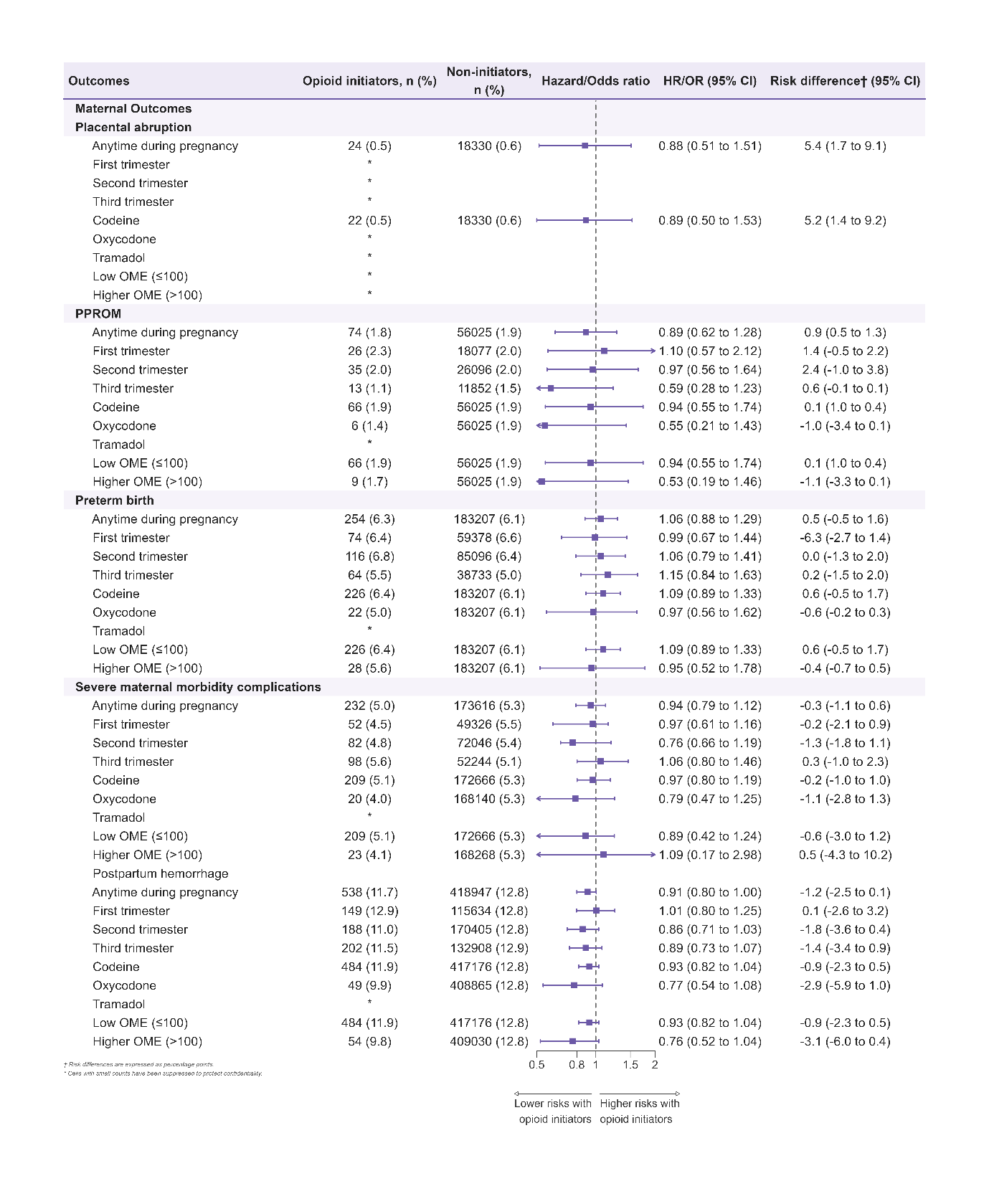

(b)

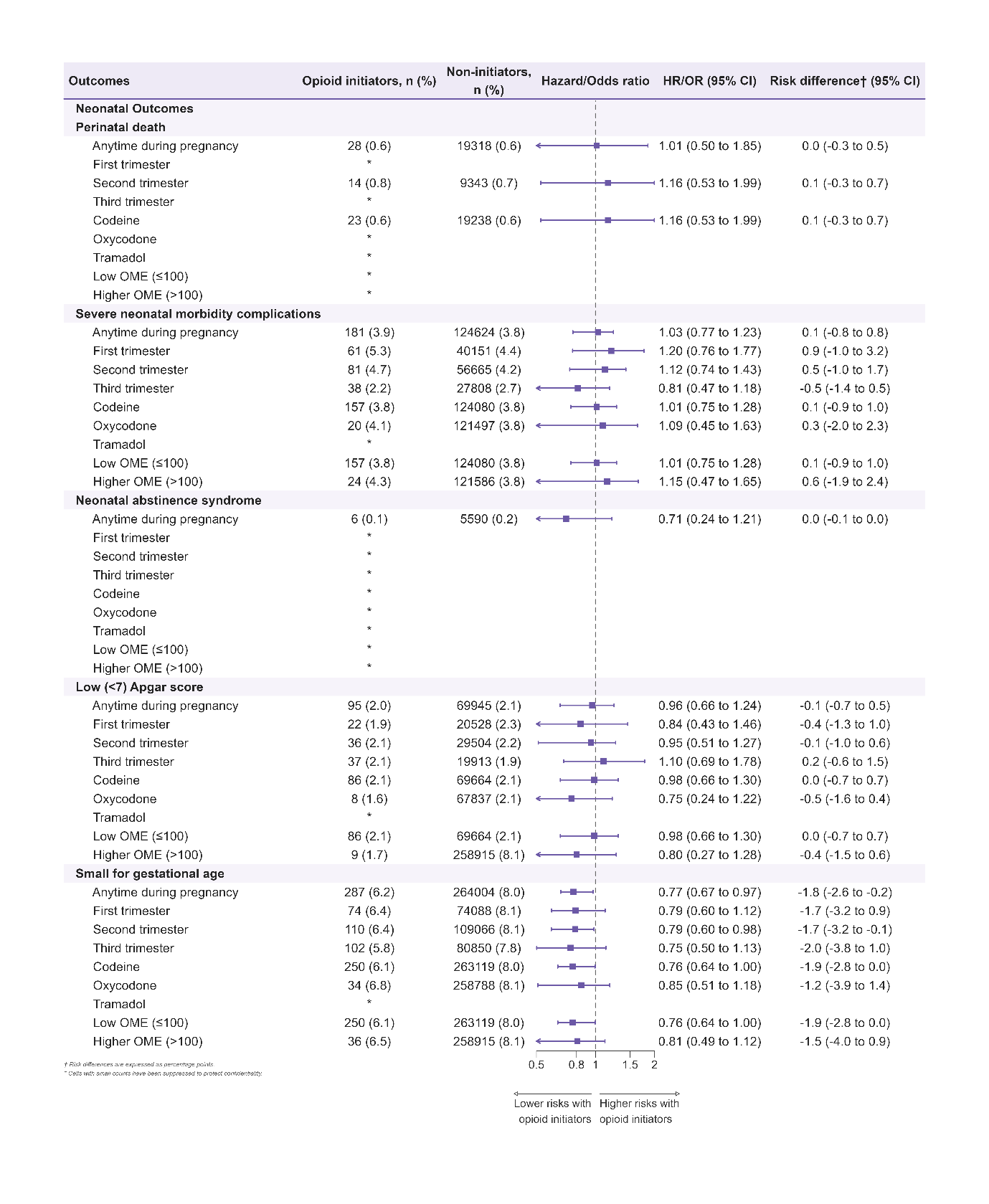

Figure 16: Sensitivity analyses of adjusted hazard or odds ratios and risk differences (expressed as percentage points) of adverse (a) maternal and (b) neonatal outcomes among pregnancies restricted to pregnancies conceived on or after February 2018, among women who initiated opioids during pregnancy compared with non‑initiators. Analyses are stratified by trimester of initiation, opioid type, and opioid dose (OME). The number and proportion of each outcome within each treatment group are shown as n (%).

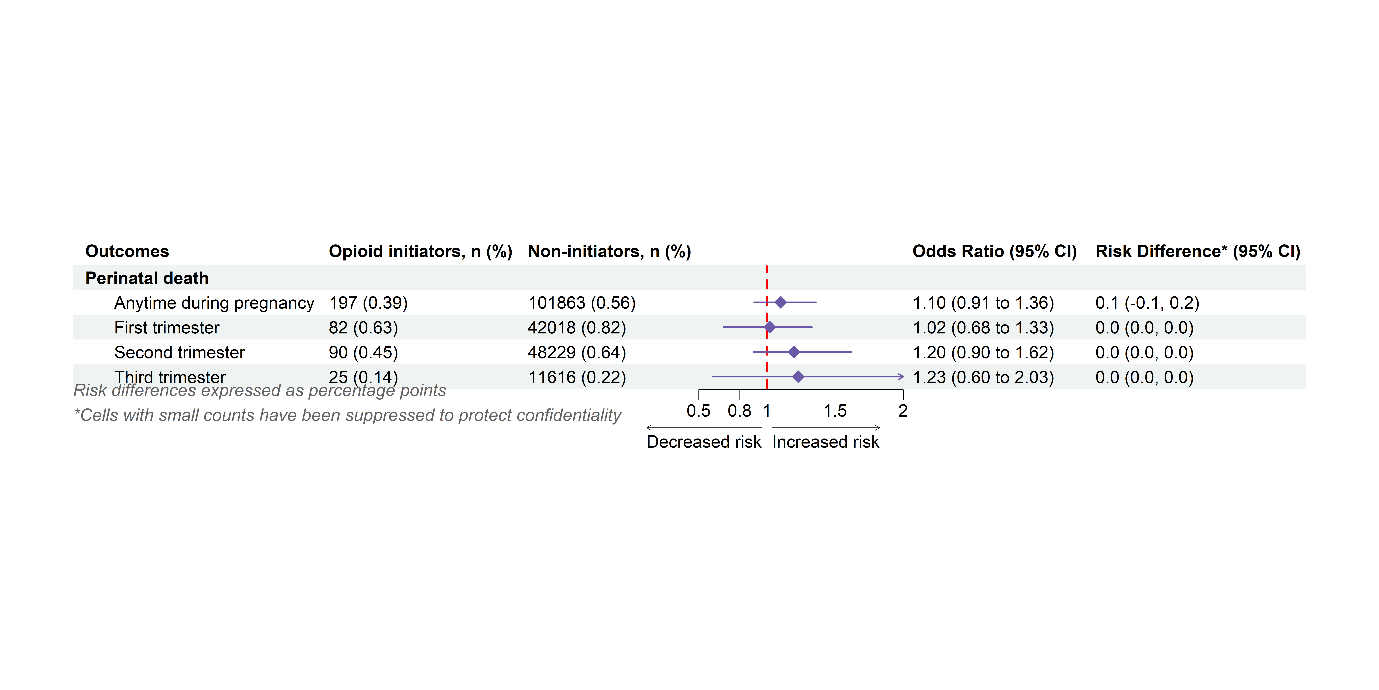

Figure 17: Post-hoc analysis shifting the date of childbirth forward by 7 days to assess potential confounding in the perinatal death outcome among women who initiated opioids during pregnancy compared with non-initiators. Analyses are stratified by trimester of initiation, and the number and proportion of each outcome within each exposure group are presented as n (%).

**a)**

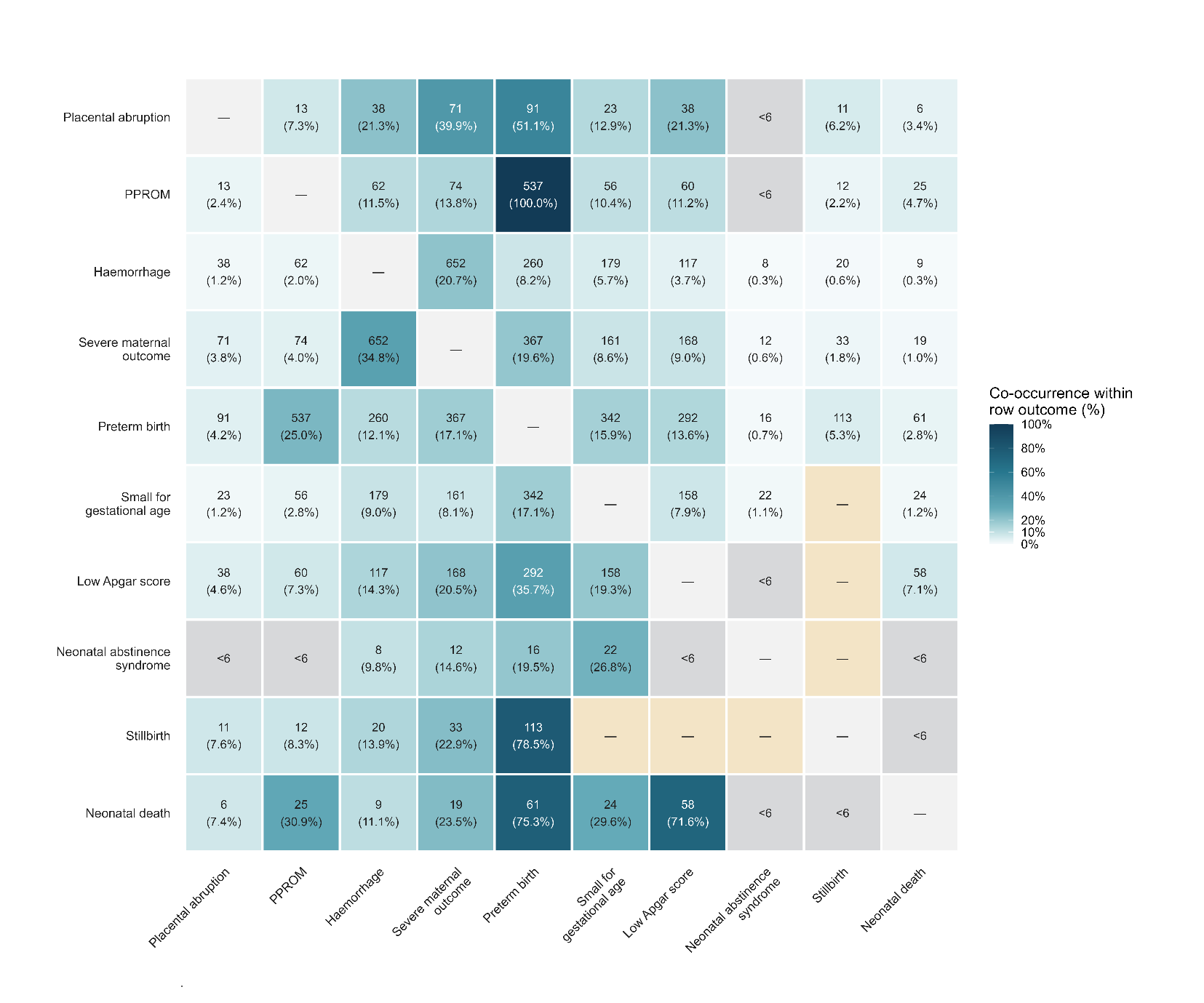

**b)**

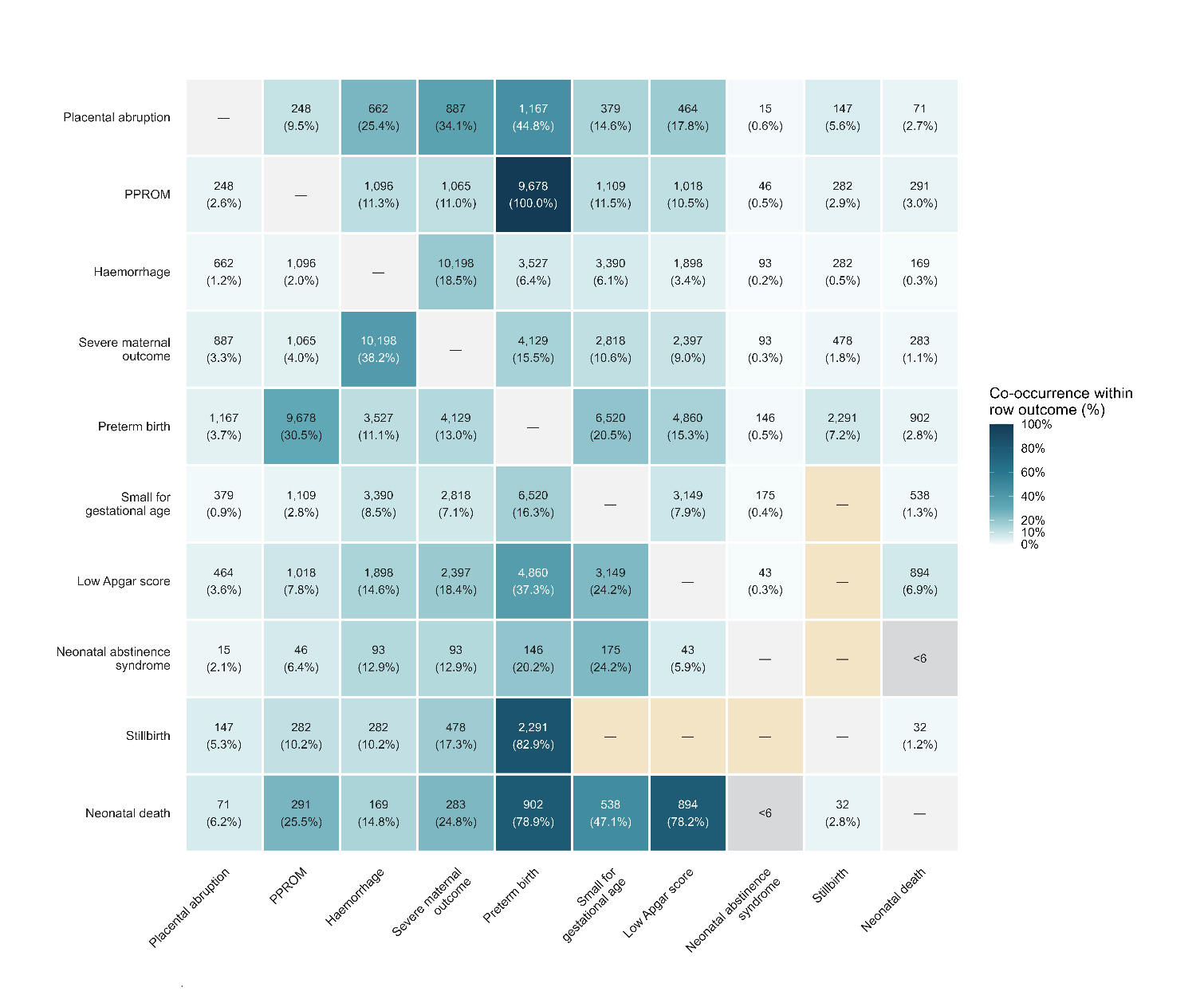
 **Figure 18:** Co-occurrence of maternal and neonatal outcomes among (a) opioid initiators and (b) non-initiators. Each cell shows the number and percentage of pregnancies with the row outcome in which the column outcome also occurred. Counts fewer than six were suppressed. Yellow cells indicate outcome combinations that were not assessed or were not applicable, while grey diagonal cells represent comparisons of an outcome with itself.

3.	National Centre for Classification in Health. The International Statistical Classification of Diseases and Related Health Problems, 10th Revision, Australian Modification (ICD-10-AM). Australia: National Centre for Classification in Health, Faculty of Health Sciences, University of Sydney; 1998.; 1998.

4.	National Centre for Classification in Health. Australian Classification of Health Interventions (ACHI). 8th ed. New South Wales, Australia: National Casemix and Classification Centre, Australian Health Services Research Institute; 2012.
